# Exposing Limitations of Clinical Laboratory Tests in COVID-19 and the Promise of Immunological Biomarkers

**DOI:** 10.1101/2022.01.29.22270016

**Authors:** Adrián Sánchez-Montalvá, Daniel Álvarez-Sierra, Mónica Martínez-Gallo, Janire Perurena-Prieto, Iria Arrese-Muñoz, Juan Carlos Ruiz Rodríguez, Juan Espinosa-Pereiro, Pau Bosch-Nicolau, Xavier Martínez-Gómez, Andrés Anton, Ferran Martínez-Valle, Mar Riveiro-Barciela, Albert Blanco-Grau, Francisco Rodríguez-Frias, Pol Castellano-Escuder, Elisabet Poyatos-Canton, Jordi Bas-Minguet, Eva Martínez-Cáceres, Alex Sánchez-Pla, Coral Zurera-Egea, Aina Teniente-Serra, Manuel Hernández González, Ricardo Pujol-Borrell, the Hospital Vall d’Hebron COVID-19 Immune Profile Group

**Affiliations:** Infectious Disease Department, Hospital Universitari Vall Hebron, Barcelona, Spain; International Health Programme Institut Català de la Salut, Vall Hebron Research Institute (VHIR), Campus Vall Hebron, Barcelona, Spain.; Department of Medicine, Universitat Autònoma Barcelona, Campus Vall Hebron, Barcelona, Spain; Translational Immunology Research Group, Vall Hebron Research Institute (VHIR), Campus Vall Hebron, Barcelona, Spain.; Immunology Department, Hospital Universitari Vall Hebron, Barcelona, Spain; Department of Cell Biology, Physiology, and Immunology, Universitat Autònoma Barcelona, Campus Vall Hebron, Barcelona, Spain.; Intensive Medicine Department, Hospital Universitari Vall Hebron, Barcelona, Spain.; Organ Dysfunction and Resuscitation Research Group, Vall Hebron Research Institute (VHIR), Campus Vall Hebron, Barcelona, Spain.; Epidemiology and Public Health Department, Hospital Universitari Vall Hebron, Campus Vall Hebron, Barcelona, Spain; Epidemiology and Public Health Research, Group, Vall Hebron Research Institute (VHIR), Campus Vall Hebron, Barcelona, Spain.; Department of Pediatrics, Obstetrics and Gynecology, Epidemiology and Pub Health, Universitat Autònoma Barcelona, Barcelona, Spain.; Microbiology Department, Hospital Universitari Vall Hebron, Barcelona, Spain.; Microbiology Research Group, Vall Hebron Research Institute (VHIR), Campus Vall Hebron, Barcelona, Spain; Department of Genetics and Microbiology, Autonomous University of Barcelona, Campus Vall Hebron, Barcelona, Spain.; Internal Medicine Department, Hospital Universitari Vall Hebron, Barcelona, Spain; Systemic Disease Research Group, Valle Hebron Research Institute (VHIR), Campus Vall Hebron, Barcelona, Spain.; Liver Disease Research Group, Valle Hebron Research Institute (VHIR), Campus Vall Hebron, Barcelona, Spain.; CIBERehd - Instituto de Salud Carlos III, Campus Vall Hebron, Barcelona, Spain; Clinical Laboratory Department, Hospital Universitari Vall Hebron, Barcelona, Spain; Bioinformatics and Statistics Group, University of Barcelona, Barcelona, Spain; Immunology Division, Bellvitge University Hospital, Hospitalet de Llobregat, Barcelona, Spain; Statistics and Bioinformatics Unit, Vall Hebron Research Institute (VHIR), Campus Vall Hebron, Barcelona Spain; Immunology Group, Health Sciences Research Institute Germans Trias Pujol (IGTP), Badalona, Barcelona, Spain; Immunology Department, Hospital Universitari Germans Trias Pujol, Badalona (Barcelona), Spain

## Abstract

**Background:** Almost two years since the onset of the COVID-19 pandemic no predictive algorithm has been generally adopted, nor new tests identified to improve the prediction and management of SARS-CoV-2 infection.

**Methods:** Retrospective observational analysis of the predictive performance of clinical parameters and laboratory tests in hospitalised patients with COVID-19. Outcomes were 28-day survival and maximal severity in a cohort of 1,579 patients and two validation cohorts of 598 and 434 patients. A pilot study conducted in a patient subgroup measured 17 cytokines and 27 lymphocyte phenotypes to explore additional predictive laboratory tests.

**Findings:** 1) Despite a strong association of 22 clinical and laboratory variables with the outcomes, their joint prediction power was limited due to redundancy. 2) Eight variables: age, comorbidity index, oxygen saturation to fraction of inspired oxygen ratio, neutrophil-lymphocyte ratio, C-reactive protein, aspartate aminotransferase/alanine aminotransferase ratio, fibrinogen, and glomerular filtration rate captured most of the statistical predictive power. 3) The interpretation of clinical and laboratory variables was improved by grouping them in categories. 4) Age and organ damage-related tests were the best predictors of survival, and inflammatory-related tests were the best predictors of severity. 5) The pilot study identified several immunological tests (including chemokine ligand 10, chemokine ligand 2, and interleukin 1 receptor antagonist), that performed better than currently used tests.

**Conclusions:** Currently used tests for clinical management of COVID-19 patients are of limited predictive value due to redundancy, as all measure aspects of two major processes: inflammation, and organ damage. There are no independent predictors based on the quality of the nascent adaptive immune response. Understanding the limitations of current tests would improve their interpretation and simplify clinical management protocols. A systematic search for better biomarkers is urgent and feasible.

This study was funded by Instituto de Salud Carlos III, Madrid, Spain, grants COV20/00416, Cov20/00654 and COV20/00388 to R.P–B, ATS and JBM respectively and co–financed by the European Regional Development Fund (ERDF). DÁ–S is recipient of a doctoral fellowship from the Vall d’Hebron Research Institute, Barcelona, Spain. ASM was supported by a postdoctoral grant “Juan Rodés” (JR18/00022) from Instituto de Salud Carlos III through the Ministry of Economy and Competitiveness, Spain

## INTRODUCTION

Two years after the onset of the coronavirus disease (COVID-19) pandemic, the clinical, laboratory, and imaging features of patients with severe acute respiratory syndrome coronavirus 2 (SARS-CoV-2) infection have been widely described[1–5]. The wide clinical spectrum of COVID-19 became obvious during the first wave, and although the effect of inoculum size should be considered [6, 7], variation has been mainly attributed to host factors, as variants of concern only appeared later [8][9]. The analysis of the first wave has obvious advantages for the identification of host factors and their biomarkers. Among host factors that affect the severity of illness, age, sex, genetic background, immunological status and prior immunity to coronaviruses[10] have been evaluated. Gene mutations of the interferon (IFN) pathway play a clear role in a small proportion of cases[11], and polymorphisms of several genes associated with immune response have been identified in genome-wide association studies[11, 12]; however, to date, the genotypes that convey a risk of severe COVID-19 have not been defined in a way that is practically applicable for prediction in clinical practice.

Reports originating from the analysis of electronic health records have confirmed the predictive value of clinical laboratory tests usually associated with poor outcomes in other infections (i.e., blood cell counts, acute-phase reactants [APRs], and coagulation factors)[14–22] but none of the proposed predictive algorithms combining demographic, clinical, and laboratory data have been widely adopted. In small case series, the state of the immune system in COVID-19 patients has been analysed using the latest tools[23–29] leading to the detection of deep perturbations in the immune system. However, inferences of the effect of these perturbations in the efficiency of the immune response and their clinical consequences are not simple and, to date, no new predictive tests have been validated or added to the clinical laboratory toolbox for COVID-19 management. We report a retrospective analysis of data from a cohort of 1,579 consecutive patients treated at the Vall d’Hebron University Hospital (HUVH) during the first wave of COVID-19 in Barcelona. We validated the main conclusions by comparison with cohorts from two other academic hospitals that belong to the same healthcare provider (the Catalan Institute of Health [ICS]) in Catalonia, Spain. We conducted the study assuming that the predictive power of clinical laboratory tests had not been fully exploited, with the main objective of improving their interpretation. A secondary objective was to explore the predictive value of a selection of robust immunological tests that might identify an early dysregulated immune response associated with severe COVID-19.

## PATIENTS AND METHODS

The database of the HUVH cohort was obtained by merging data sets from the Infectious Disease, Epidemiology and Clinical Laboratory departments. Consecutive patients aged ≥18 years with a SARS-CoV-2 positive polymerase chain reaction (PCR) from any respiratory sample, hospitalised in HUVH between 10 March and 29 April 2020 were included in the study (see Tables 1 and 1S). This COVID-19 HUVH cohort consisted of 1,579 patients (Fig 1). All patient medical records included the number of symptoms, days from symptom onset (DFSO), initial assessment of vital signs, comorbidities, length of hospital stay (LOS), intensive care unit (ICU) admission, oxygen supplementation and supportive ventilation requirements, outcome during the hospitalisation and results from clinical laboratory tests. Data were censored on the date of discharge, death, or 28 days after admission, whichever occurred first. The outcome of all patients discharged before the 28th day was ascertained through a review of the primary care electronic health record notes. Comorbidities were classified as 1) cardiovascular disease and/or hypertension, 2) chronic lung disease, 3) diabetes, 4) neurological disease, 5) chronic kidney disease, 6) active non-terminal malignancy, 7) obesity, and 8) chronic liver disease. Each comorbidity was assigned value of 1, and a global comorbidity index (1 to 8) was generated. The clinical severity category was assigned as the maximal score attained during hospitalisation, using a simplified version of the World Health Organization (WHO) 10-point COVID-19 disease clinical progression score[30] as follows: 1) Mild, no activity limitations or not requiring hospitalisation; 2) Moderate, hospitalised, not requiring high-flow oxygen therapy or ventilation support; 3) Severe, hospitalised requiring high-flow oxygen therapy or ventilation support; and 4) Deceased, those who died before day 28 of hospitalisation. These categories correspond to the WHO scores 1–3, 4–5, 6–9, and 10, respectively. For some analyses, the mild and moderate categories were combined into a non-severe category, and the severe and deceased categories were combined into a severe category.

**Fig 1.**
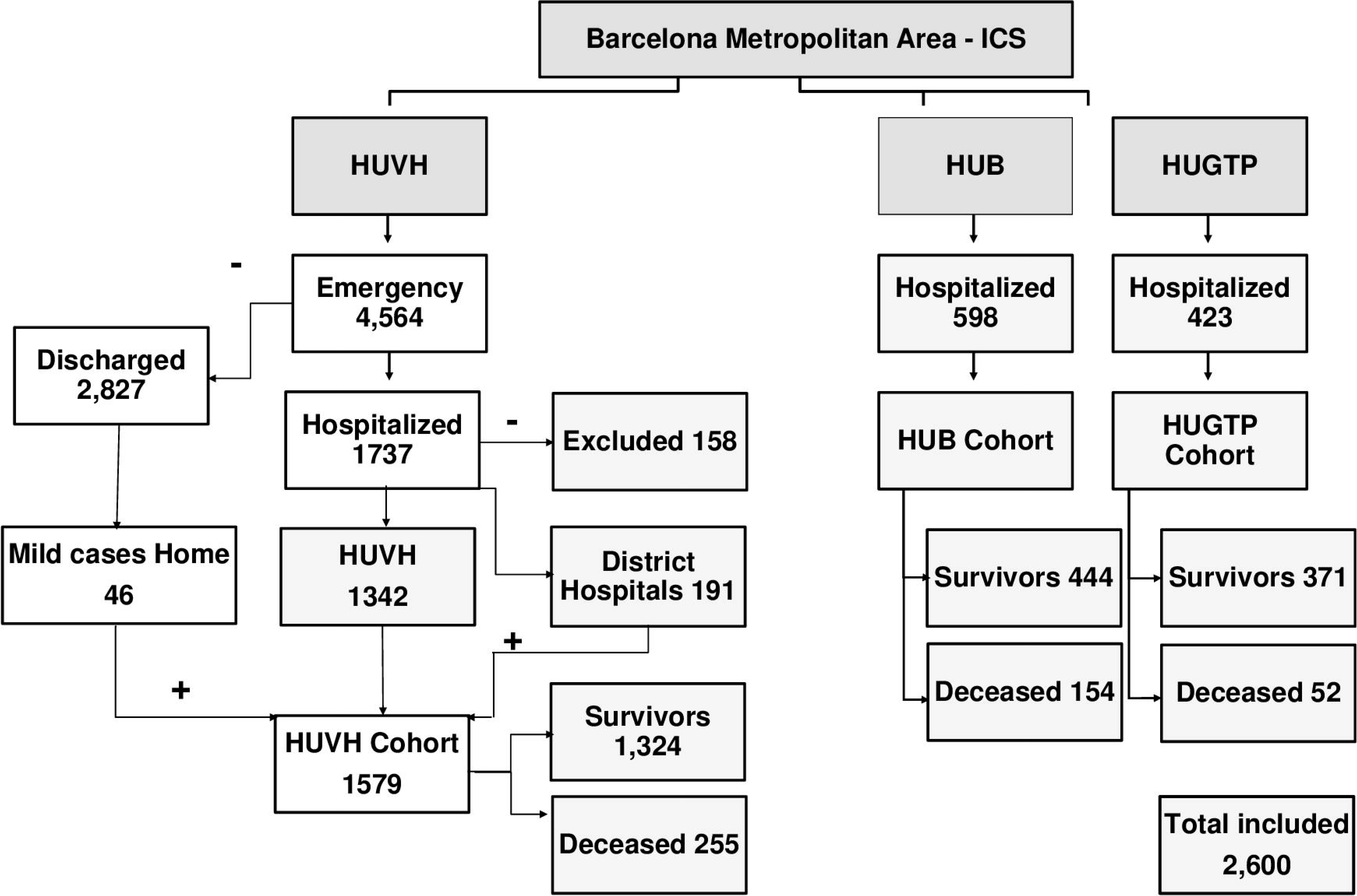
Selection of patients for the cohorts from Vall d’Hebron University Hospital (HUVH), Bellvitge University Hospital (HUB), and Germans Trias i Pujol University Hospital (HUGTP). All patients were confirmed by polymerase chain reaction (PCR) to have coronavirus disease (COVID-19). The details of the excluded patients are provided in Table 1S. The data from HUVH corresponds to patients who were admitted to the emergency division between 10 March and 29 April 2020; to HUGTP between 17 March and 12 May 2020; and to HUB between 16 March and 23 September 2020. The number of deceased patients corresponds to the 28-day follow-up period. The HUB and HUGTP cohorts include only hospitalised patients.

**Table 1.** Summary of the clinical and demographic features of HUVH cohort.

|  |  | Survivors vs deceased |  |  |  | Non-severe vs severe |  |  |
| --- | --- | --- | --- | --- | --- | --- | --- | --- |
|  |  | Patients<br>n (%) | Survivors<br>n (%) | Deceased<br>n (%) | *p*-value | Non-severe<br>n (%) | Severe<br>n (%) | *p*-value |
| **All** |  | 1579 (100) | 1324 (83.9) | 255 (16.1) | NA | 1040 (63.8) | 539 (34.1) | NA |
| **Female** |  | 699 (44.3) | 592 (44.7) | 107 (42.0) | 0.449 | 491 (47.2) | 208 (38.6) | **0.001** |
| **Male** |  | 880 (55.7) | 732 (55.3) | 148 (58.0) |  | 549 (52.8) | 331 (61.4) |  |
| **ICU** |  | 236 (14.9) | 203 (15.3) | 33 (13.9) | 0.4 | 1 (0.1) | 235 (43.6) | **1.612E-126** |
|  |  | Median (IQR) | Median (IQR) | Median (IQR) | *p*-value | Median (IQR) | Median (IQR) | *p*-value |
| **Age, years** |  | 62 (50–75) | 57 (48–70) | 82 (74–87) | 7.26E-81 | 58 (47–71) | 70 (54–82) | **2.4872E-29** |
| **DFSO** |  | 7 (4–9) | 7 (5–10) | 5 (2–7) | 9.40E-19 | 7 (5–10) | 6 (3–8) | **2.9561E-14** |
| **LOS** |  | 7 (2–20) | 7 (2–24) | 7 (4–11) | 0.4226 | 5 (2–10) | 14 (6–36) | **1.5748E-44** |
| **Disease duration, days** |  | 15.0 (10–28) | 15 (10–34) | 12 (9–18) | 4.45E-11 | 13 (9–21) | 22 (11–43) | **1.4952E-17** |
| Clinical Presentation |  | n (%) | n (%) | n (%) | *p*-value | n (%) | n (%) | *p*-value |
| **General** | Fever | 1314 (83.2) | 1132 (85.4) | 199 (75.3) | 0.040 | 886 (85.2) | 438 (81.3) | 0.051 |
| **Respiratory** | Upper respiratory symptoms (only) | 94 (5.9) | 85 (6.5) | 9 (3.4) | 0.063 | 69 (6.5) | 25 (4.6) | 0.118 |
|  | Lower respiratory symptoms | 1351 (85.6) | 1148 (86.7) | 203 (79.6) | 0.020 | 892 (85.8) | 459 (85.2) | 0.763 |
|  | Pneumonia | 1524 (96.5) | 1271 (96.0) | 253 (99.2) | 0.008 | 989 (95.1) | 535 (99.3) | **3.62E-06** |
| **Digestive** | All | 491 (31.1) | 440 (33.2) | 51 (20.0) | 1.182E-05 | 353 (33.9) | 138 (25.6) | **7.11E-04** |
| Comorbidities |  | n (%) | n (%) | n (%) | *p*-value | n (%) | n (%) | *p*-value |
|  | Cardiovascular & hypertension | 712 (45.1) | 508 (38.4) | 204 (80.0) | 8.534E-37 | 394 (37.9) | 318 (59.0) | **1.463E-15** |
|  | Chronic lung disease | 278 (17.6) | 198 (15.0) | 80 (31.4) | 5.134E-09 | 145 (13.9) | 133 (24.7) | **2.143E-07** |
|  | Diabetes | 293 (18.5) | 223 (16.8) | 70 (28.3) | 1.90E-05 | 170 (16.3) | 123 (22.8) | **0.002** |
|  | Neurological disease | 227 (14.4) | 156 (11.8) | 71 (27.8) | 1.29E-09 | 121 (11.6) | 106 (19.7) | **2.89E-05** |
|  | Chronic kidney disease | 134 (8.5) | 79 (6) | 55 (21.6) | 1.03E-13 | 55 (5.3) | 79 (14.7) | **9.101E-10** |
|  | Active non-terminal malignancy | 113 (7.2) | 62 (4.7) | 51 (20.0) | 1.10E-15 | 41 (3.9) | 72 (13.4) | **4.197E-11** |
|  | Obesity | 261 (16.5) | 227 (17.3) | 34 (12.8) | 0.85 | 145 (13.9) | 116 (21.5) | **2.00E-04** |
|  | Chronic liver disease | 61 (3.9) | 48 (3.7) | 13 (4.9) | 0.139 | 37 (3.6) | 24 (4.5) | 0.409 |
| Comorbidity index* |  | Median (IQR) | Median (IQR) | Median (IQR) | *p*-value | Median (IQR) | Median (IQR) | *p*-value |
|  |  | 1 (0-2) | 2 (1-3) | 2 (1-3) | 2.2961E-38 | 1 (0-2) | 2 (1-3) | **1.0118E-26** |

\* The breakdown of patients by comorbidity index is in Table 1S.
Exact *p*-values from the Mann–Whitney U test, Significance should be considered for $p \leq 0.001$ after Bonferroni. F, female; M, male; DFSD, days from symptom onset; LOS, length

The validation cohorts from the Bellvitge University Hospital (HUB) and the Germans Trias i Pujol University Hospital (HUGTP) included 598 and 423 patients, respectively, and, together with the HUVH cohort, at total of 2,600 patients were included in the analysis.

### Outcomes

Final outcomes for comparison included survival vs. death, and maximum clinical severity. For the validation cohorts the only available outcome was survival for 28 days (survivors) and death (deceased).

### Clinical laboratory tests

Detection of SARS-CoV-2 was first performed by an in-house PCR assay with primers and probes from 2019-nCoV CDC PCR panel, using the One-Step RT-PCR kit (Qiagen, Germany). When commercial assays became available, a real-time multiplex RT-PCR assay (Allplex^TM^ 2019-nCoV Assay, Seegene, South Korea) was used.

The clinical laboratories were equipped with Beckman Coulter (Brea, CA, USA) and Roche Diagnostics (Indianapolis, IN, USA) automatic analysers that were integrated with two TECAN (Zug, Switzerland) continuous lines and two automatic cold storage and retrieval units that ensure sample integrity. Interleukin 6 (IL-6) levels were measured in a Elycsis® Cobas analyser (Roche). Samples for assessing the predictive performance of clinical laboratory tests were taken on admission to the hospital; glomerular filtration rate (GFR) was calculated by applying the algorithm of Levey et al.[31]; additional laboratory test data for the 28-day follow-up period were available from 9,475 samples corresponding to 1,079 of the 1,579 patients in the HUVH cohort.

### Immunological tests

The levels of cytokines chemokine ligand 2 (CCL2), chemokine ligand 10 (CXCL10), granulocyte-macrophage colony-stimulating factor (GM-CSF), interferon (IFN)-alpha, IFN-gamma, interleukin (IL)-10, IL-12 p70, IL-13, IL-15, IL-17A, IL-1RA, IL-2, IL-4, IL-6, IL-7, and tumour-necrosis factor (TNF), and granzyme B were measured in sera using the ELLA microfluidic platform (Biotechne®, Minneapolis, MN, USA); for sCD163 levels an enzyme-linked immunosorbent assay was used (CD163 human kit, Thermo Fisher Societies, Waltham, MA, USA).

The Human Immune Phenotyping Consortium protocol [32, 33] was adapted for the study of COVID-19 patients. The antibodies used are shown in Table 2S. Blood was collected in EDTA vacutainer tubes (BD-Plymouth, UK) and processed within 4 hours. A total of 10^5^ events were acquired from each sample in most cases, using a NAVIOS EX flow cytometer (Beckman Coulter). Data were analysed with Kaluza Beckman Software v.2.1. Absolute values were generated by loading counts from the haematological analyser (XN-2000; Sysmex, Japan) parallel sample analysis.

### Statistical analysis

Categorical variables were summarised as frequencies and proportions and continuous variables as means, standard deviations, and 95% confidence intervals (CIs) or medians and interquartile ranges (IQRs), depending on their distribution. Pairwise comparisons used the Mann–Whitney U-test and Kruskal-Wallis test, adjusted for the false-discovery rate (FDR) using the Benjamini and Hochberg, or Bonferroni method (where indicated). C-reactive protein (CRP), IL-6, ferritin, and D-dimer values were logarithmically transformed. A threshold of 30% of laboratory missing data was used as the exclusion criteria for data analyses. As an exception, because the initial oxygen saturation to fraction of inspired oxygen ratio (SpO_2_/FiO_2_) value while breathing room air was available for a subset of 827 patients, but this is crucial variable, the data from these patients were either analysed separately, or when this parameter, was included in a general analysis, this is indicated in the text.

Bivariable logistic regression was used to calculate the age-adjusted odd ratios (OR) and effect size (Z score) of each variable. Multivariable logistic regression was used to calculate the predictive power of different combinations of variables. Correlation among variables was analysed using the non-parametric Spearman test. For analysis of follow-up data of the HUVH cohort, locally weighted smoothing (LOESS) was applied to clinical laboratory variables to visualise the relationship between the mean and CI of each variable, time and 28-day outcome, as described by Abers et al.[1]. To assess the performance of each clinical laboratory test, the receiver-operating characteristic (ROC) curve and the corresponding area under the curve (AUC) values were calculated. In addition, random forest simulation and principal component analysis (PCA) were performed to further compare the influence of the laboratory and clinical variables on the outcomes in each hospital dataset.

Statistical tests were 2-sided and used a significance threshold of at least p <0.05. R, version 4.1.0 (The R Foundation for Statistical Computing, Vienna, Austria) and Prism 9® (GraphPad, San Diego, CA, USA) packages were used for all analyses. Statistical analysis was conducted by the Statistics and Bioinformatics Unit (UEB), Vall d’Hebron Hospital Research Institute, and by co-authors PC-E and RP-B under the supervision of UEB.

### Ethical review

This project was approved by the institutional ethics board of the three institutions (HUVH, HUGTP, and HUB) which waived the requirement for individual informed consent (protocol R(AG)242/2020). In the HUVH cohort, residual sera samples were transferred to the Vall d’Hebron University Hospital Biobank (PT17/0015/0047) part of the Carlos III Institute of Health network of biobanks (Number C.0006012).

## RESULTS

### Overall clinical features of HUVH cohort

The HUVH cohort included 1,579 PCR-confirmed COVID-19 patients with a median age of was 62 years (IQR: 50–75 years), of whom 255 (16.1%) died during the first 28 days after hospitalisation (Fig 2A). Eight hundred eighty (55.7%) patients were male; this proportion was higher among the deceased patients (58.0%) and also higher than that of males in the Barcelona metropolitan area at the time of the pandemic first peak (47.5%, p <0.001) [34, 35]. A total of 236 (14.9%) patients were admitted to the ICU with a 28-day case fatality rate of 13.9%[35].

**Fig 2.**
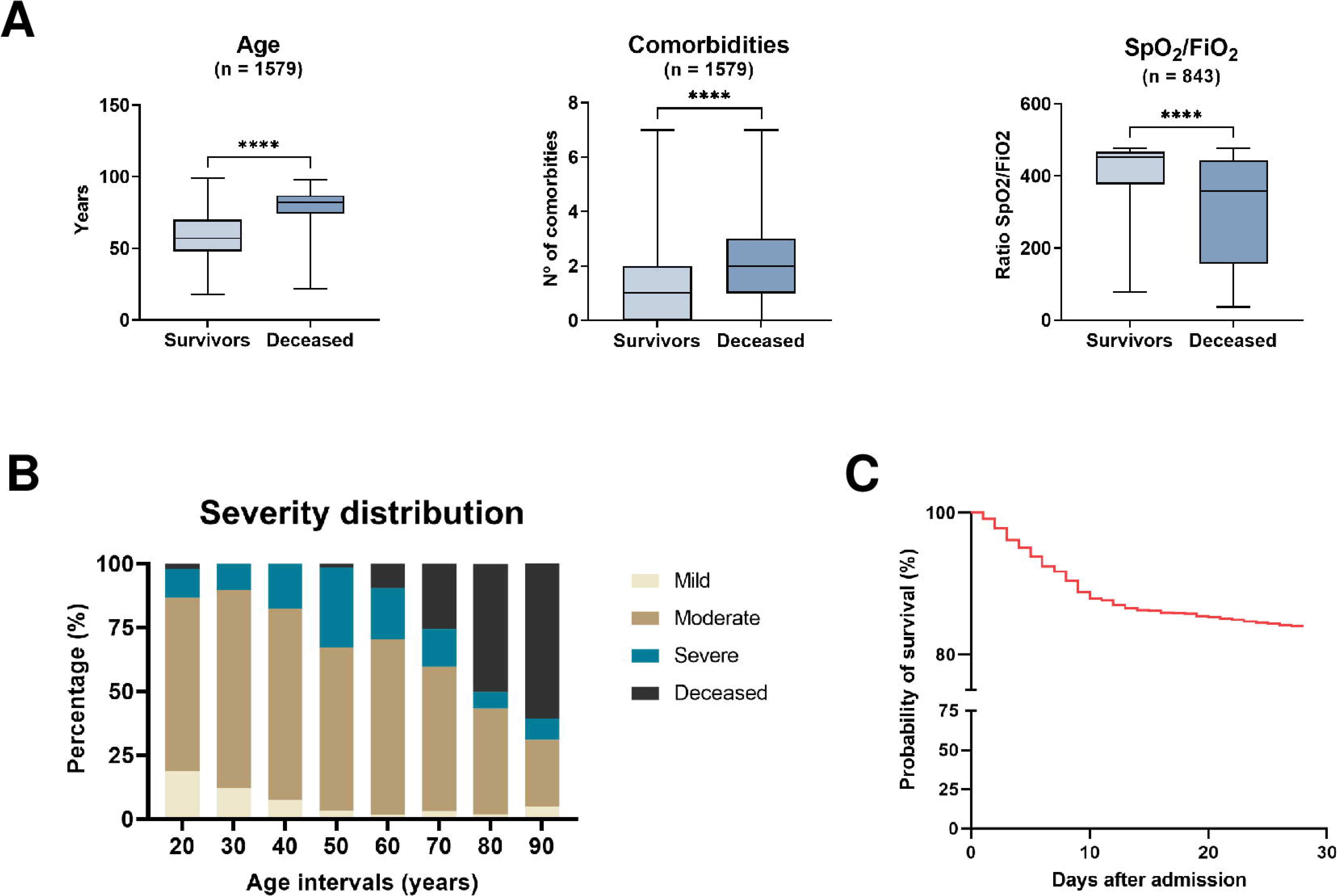
The structure and outcomes of the Vall d’Hebron University Hospital (HUVH) cohort. (A) Left panel, age distribution of the survivors and that of the deceased is markedly different (median [IQR]: 62 years [50–75 years] vs. 82 years [74–87 years], p<0.001) as are comorbidities (central panel) and SpO_2_/FiO_2_ (right panel). (B) Distribution of the patients in the HUVH cohort among the four severity categories, based on the World Health Organization criteria (described in the Material and Methods section). The number of patients in the mild category is small (n=71) as only patients with bilateral pneumonia or severe associated pathologies were hospitalised during this period of the pandemic. (C) Survival after admission: this graph highlights mortality during the initial 10 days, with a high number of patients older than 80 years dying in the initial 3–4 days (see text “Overall Clinical Features of HUVH cohort”).

The presenting symptoms are shown in Table 1. Of note, digestive symptoms were more frequent in survivors (31.1% vs. 20.0%, p <0.001). Cardiovascular and/or hypertension, chronic lung disease, diabetes, neurological disease, chronic kidney disease, and active non-terminal malignancy were associated to decreased 28-day survival, but not chronic liver disease nor obesity. The comorbidity index was significantly higher in deceased patients and patients with severe disease than in survivors and patients with non-severe disease. Each comorbidity added 10% mortality risk up to an index of 4 (Table 3S).

The distribution of disease severity was as follows, 71 (4.5%), 969 (61.4%), 284 (17.9%), and 255 (16.1%) in the mild, moderate, severe, and deceased categories, respectively. The age of patients increased with increasing disease severity category, except between the moderate and severe disease groups (Fig 2B and Table 4S). The LOS increased with disease severity for the three initial disease severity categories but was shorter among the deceased because 24.9% of the deceased patients died during the initial 4 days of hospitalisation (Fig 2C). The median disease duration was 18 days (IQR: 10–18 days) and was progressively longer with increasing disease severity (Table 5S). Age had a strong effect on mortality: for patients in the age groups 50–59, 60–69, 70–79, 80– 89 and >90 years, with 28-day case fatality rates of 1.82%, 10.9%, 26.4%, 49.7% and 60.6% respectively.

In the dichotomous disease severity grouping, there were 1,040 and 539 patients in the non-severe and severe categories, respectively. Deceased patients accounted for 43.7% of the severe category. The disease severity was significantly associated with age, DFSO, LOS, disease duration, and comorbidities other than chronic liver disease. Disease severity was greater in males than in females, but after adjusting for multiple comparisons the statistical significance was moderate compared with the other statistically significant associations (exact p =0.001, after Bonferroni’s correction p=0.03) (Table 1).

### Predictive power of current clinical laboratory tests

The exploratory statistical analysis of the HUVH cohort revealed that, despite the strong association of 22 of the 30 variables with 28-day outcomes (Fig 3 and “S and Tables 2 and 5S), the predictive power of the combined variables was limited and appeared to rely disproportionally on age, a non-laboratory variable (Table 3). Further analyses described in the supplementary section “Sequence of statistical biomarker analyses” were undertaken to determine the reason of this limitation, as summarised below

**Fig 3.**
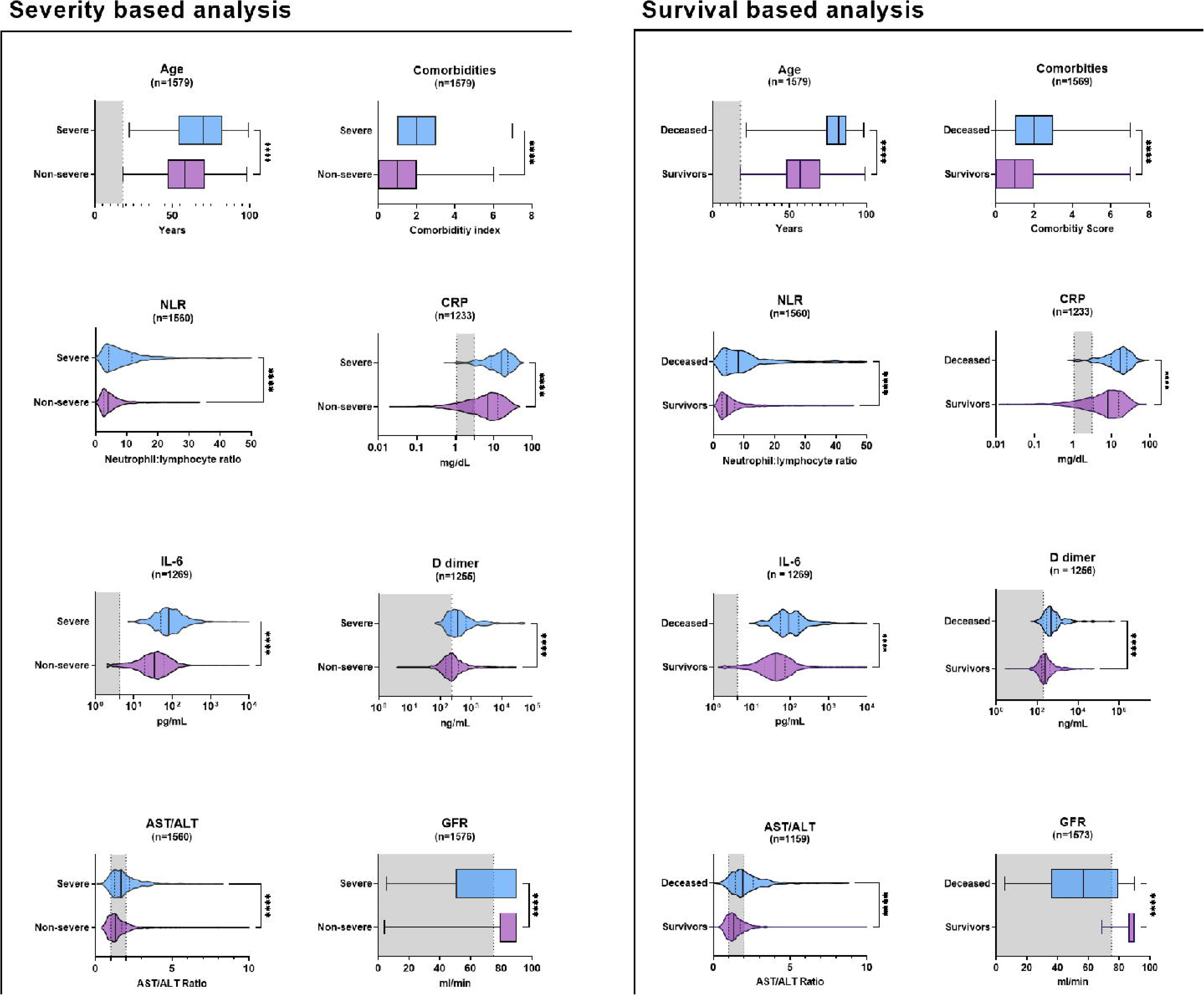
Pairwise comparisons of a selection of clinical laboratory-derived variables at admission and 28-days survival for the survival/decease and non-severe, severe outcomes in the Vall d’Hebron University Hospital cohort. N, number of cases plotted; NLR, neutrophil-to-lymphocyte ratio; CRP, C-reactive protein; AST, aspartate aminotransferase; ALT, alanine aminotransferase; GFR, glomerular filtration rate. * p <0.05; ** p <0.01; *** p <0.001; **** p <0.0001. When non-significant, the numerical p-values are given. The exact p-values for all tests are given in Table 2. Of note, the distribution of age and GFR values are different in the severity and survival analysis. The grey area indicates the normal range of the test.

**Table 2.**
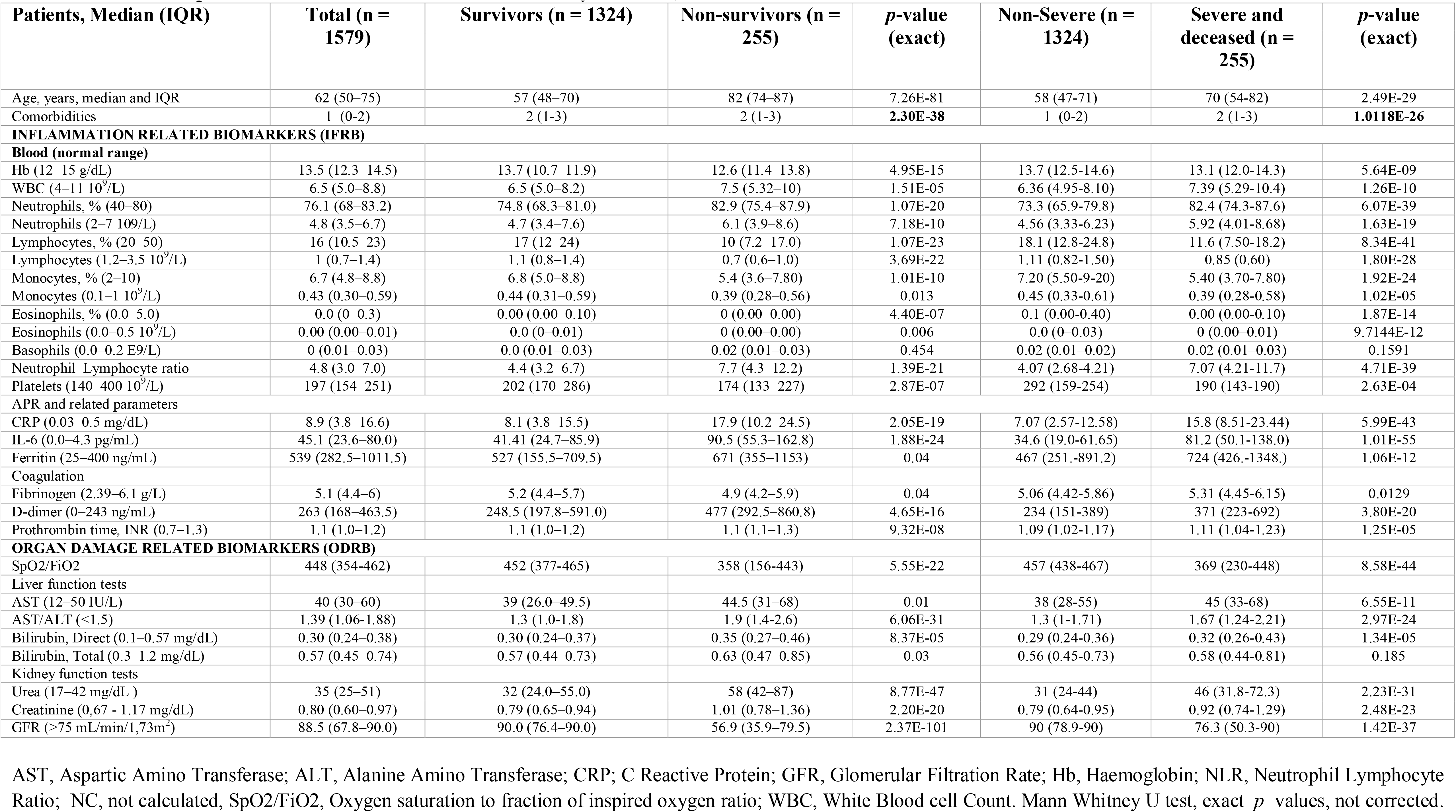
Pairwise comparison of biomarkers for decease and severity outcomes, HUVH cohort.

| Patients, Median (IQR) | Total (n = 1579) | Survivors (n = 1324) | Non-survivors (n = 255) | *p*-value (exact) | Non-Severe (n = 1324) | Severe and deceased (n = 255) | *p*-value (exact) |
| --- | --- | --- | --- | --- | --- | --- | --- |
| Age, years, median and IQR | 62 (50–75) | 57 (48–70) | 82 (74–87) | 7.26E-81 | 58 (47-71) | 70 (54-82) | 2.49E-29 |
| Comorbidities | 1 (0-2) | 2 (1-3) | 2 (1-3) | **2.30E-38** | 1 (0-2) | 2 (1-3) | **1.0118E-26** |
| **INFLAMMATION RELATED BIOMARKERS (IFRB)** |  |  |  |  |  |  |  |
| **Blood (normal range)** |  |  |  |  |  |  |  |
| Hb (12–15 g/dL) | 13.5 (12.3–14.5) | 13.7 (10.7–11.9) | 12.6 (11.4–13.8) | 4.95E-15 | 13.7 (12.5-14.6) | 13.1 (12.0-14.3) | 5.64E-09 |
| WBC (4–11 10<sup>9</sup>/L) | 6.5 (5.0–8.8) | 6.5 (5.0–8.2) | 7.5 (5.32–10) | 1.51E-05 | 6.36 (4.95-8.10) | 7.39 (5.29-10.4) | 1.26E-10 |
| Neutrophils, % (40–80) | 76.1 (68–83.2) | 74.8 (68.3–81.0) | 82.9 (75.4–87.9) | 1.07E-20 | 73.3 (65.9-79.8) | 82.4 (74.3-87.6) | 6.07E-39 |
| Neutrophils (2–7 10<sup>9</sup>/L) | 4.8 (3.5–6.7) | 4.7 (3.4–7.6) | 6.1 (3.9–8.6) | 7.18E-10 | 4.56 (3.33-6.23) | 5.92 (4.01-8.68) | 1.63E-19 |
| Lymphocytes, % (20–50) | 16 (10.5–23) | 17 (12–24) | 10 (7.2–17.0) | 1.07E-23 | 18.1 (12.8-24.8) | 11.6 (7.50-18.2) | 8.34E-41 |
| Lymphocytes (1.2–3.5 10<sup>9</sup>/L) | 1 (0.7–1.4) | 1.1 (0.8–1.4) | 0.7 (0.6–1.0) | 3.69E-22 | 1.11 (0.82-1.50) | 0.85 (0.60) | 1.80E-28 |
| Monocytes, % (2–10) | 6.7 (4.8–8.8) | 6.8 (5.0–8.8) | 5.4 (3.6–7.80) | 1.01E-10 | 7.20 (5.50-9.20) | 5.40 (3.70-7.80) | 1.92E-24 |
| Monocytes (0.1–1 10<sup>9</sup>/L) | 0.43 (0.30–0.59) | 0.44 (0.31–0.59) | 0.39 (0.28–0.56) | 0.013 | 0.45 (0.33-0.61) | 0.39 (0.28-0.58) | 1.02E-05 |
| Eosinophils, % (0.0–5.0) | 0.0 (0–0.3) | 0.00 (0.00–0.10) | 0 (0.00–0.00) | 4.40E-07 | 0.1 (0.00-0.40) | 0.00 (0.00-0.10) | 1.87E-14 |
| Eosinophils (0.0–0.5 10<sup>9</sup>/L) | 0.00 (0.00–0.01) | 0.0 (0–0.01) | 0 (0.00–0.00) | 0.006 | 0.0 (0–0.03) | 0 (0.00–0.01) | 9.7144E-12 |
| Basophils (0.0–0.2 E9/L) | 0 (0.01–0.03) | 0.0 (0.01–0.03) | 0.02 (0.01–0.03) | 0.454 | 0.02 (0.01–0.02) | 0.02 (0.01–0.03) | 0.1591 |
| Neutrophil–Lymphocyte ratio | 4.8 (3.0–7.0) | 4.4 (3.2–6.7) | 7.7 (4.3–12.2) | 1.39E-21 | 4.07 (2.68-4.21) | 7.07 (4.21-11.7) | 4.71E-39 |
| Platelets (140–400 10<sup>9</sup>/L) | 197 (154–251) | 202 (170–286) | 174 (133–227) | 2.87E-07 | 292 (159-254) | 190 (143-190) | 2.63E-04 |
| **APR and related parameters** |  |  |  |  |  |  |  |
| CRP (0.03–0.5 mg/dL) | 8.9 (3.8–16.6) | 8.1 (3.8–15.5) | 17.9 (10.2–24.5) | 2.05E-19 | 7.07 (2.57-12.58) | 15.8 (8.51-23.44) | 5.99E-43 |
| IL-6 (0.0–4.3 pg/mL) | 45.1 (23.6–80.0) | 41.41 (24.7–85.9) | 90.5 (55.3–162.8) | 1.88E-24 | 34.6 (19.0-61.65) | 81.2 (50.1-138.0) | 1.01E-55 |
| Ferritin (25–400 ng/mL) | 539 (282.5–1011.5) | 527 (155.5–709.5) | 671 (355–1153) | 0.04 | 467 (251.-891.2) | 724 (426.-1348.) | 1.06E-12 |
| **Coagulation** |  |  |  |  |  |  |  |
| Fibrinogen (2.39–6.1 g/L) | 5.1 (4.4–6) | 5.2 (4.4–5.7) | 4.9 (4.2–5.9) | 0.04 | 5.06 (4.42-5.86) | 5.31 (4.45-6.15) | 0.0129 |
| D-dimer (0–243 ng/mL) | 263 (168–463.5) | 248.5 (197.8–591.0) | 477 (292.5–860.8) | 4.65E-16 | 234 (151-389) | 371 (223-692) | 3.80E-20 |
| Prothrombin time, INR (0.7–1.3) | 1.1 (1.0–1.2) | 1.1 (1.0–1.2) | 1.1 (1.1–1.3) | 9.32E-08 | 1.09 (1.02-1.17) | 1.11 (1.04-1.23) | 1.25E-05 |
| **ORGAN DAMAGE RELATED BIOMARKERS (ODRB)** |  |  |  |  |  |  |  |
| SpO2/FiO2 | 448 (354-462) | 452 (377-465) | 358 (156-443) | 5.55E-22 | 457 (438-467) | 369 (230-448) | 8.58E-44 |
| **Liver function tests** |  |  |  |  |  |  |  |
| AST (12–50 IU/L) | 40 (30–60) | 39 (26.0–49.5) | 44.5 (31–68) | 0.01 | 38 (28-55) | 45 (33-68) | 6.55E-11 |
| AST/ALT (<1.5) | 1.39 (1.06-1.88) | 1.3 (1.0-1.8) | 1.9 (1.4-2.6) | 6.06E-31 | 1.3 (1-1.71) | 1.67 (1.24-2.21) | 2.97E-24 |
| Bilirubin, Direct (0.1–0.57 mg/dL) | 0.30 (0.24–0.38) | 0.30 (0.24–0.37) | 0.35 (0.27–0.46) | 8.37E-05 | 0.29 (0.24-0.36) | 0.32 (0.26-0.43) | 1.34E-05 |
| Bilirubin, Total (0.3–1.2 mg/dL) | 0.57 (0.45–0.74) | 0.57 (0.44–0.73) | 0.63 (0.47–0.85) | 0.03 | 0.56 (0.45-0.73) | 0.58 (0.44-0.81) | 0.185 |
| **Kidney function tests** |  |  |  |  |  |  |  |
| Urea (17–42 mg/dL) | 35 (25–51) | 32 (24.0–55.0) | 58 (42–87) | 8.77E-47 | 31 (24-44) | 46 (31.8-72.3) | 2.23E-31 |
| Creatinine (0.67 - 1.17 mg/dL) | 0.80 (0.60–0.97) | 0.79 (0.65–0.94) | 1.01 (0.78–1.36) | 2.20E-20 | 0.79 (0.64-0.95) | 0.92 (0.74-1.29) | 2.48E-23 |
| GFR (>75 mL/min/1.73m<sup>2</sup>) | 88.5 (67.8–90.0) | 90.0 (76.4–90.0) | 56.9 (35.9–79.5) | 2.37E-101 | 90 (78.9-90) | 76.3 (50.3-90) | 1.42E-37 |

AST, Aspartic Amino Transferase; ALT, Alanine Amino Transferase; CRP; C Reactive Protein; GFR, Glomerular Filtration Rate; Hb, Haemoglobin; NLR, Neutrophil Lymphocyte Ratio; NC, not calculated, SpO2/FiO2, Oxygen saturation to fraction of inspired oxygen ratio; WBC, White Blood cell Count. Mann Whitney U test, exact *p* values, not corrected.

**Table 3.** Classification tables from multiple logistic regression including different sets of variables for survival vs decease or severe vs non-severe as outcome.

| **Predicted outcome, 19 clinical and laboratory variables\* for survival versus decease** |  |  |  |  |
| --- | --- | --- | --- | --- |
| Classification table | Predicted survival | Predicted decease | Total | % Correctly classified |
| Observed Survival | 357 | 8 | 365 | 97.81 |
| Observed Decease | 21 | 25 | 46 | **54.35** |
| Total | 378 | 33 | 411 | 92.94 |
| **Predicted outcome, 19 clinical and laboratory variables\* severe versus non-severe** |  |  |  |  |
| Classification table | Predicted non-severe | Predicted severe | Total | % Correctly classified |
| Observed non-severe | 244 | 26 | 270 | 90.37 |
| Observed severe | 64 | 77 | 141 | **54.61** |
| Total | 308 | 103 | 411 | 78.10 |
| **Predicted outcome, 16 non-clinical variables\*\* for survival versus decease** |  |  |  |  |
| Classification table | Predicted survival | Predicted decease | Total | % Correctly classified |
| Observed survival | 356 | 9 | 365 | 97.53 |
| Observed decease | 30 | 16 | 46 | **34.78** |
| Total | 386 | 25 | 411 | 90.51 |
| **Predicted outcome, 16 non-clinical variables\*\* for severe versus non-severe** |  |  |  |  |
| Classification table | Predicted non-severe | Predicted severe | Total | % Correctly classified |
| Observed non-severe | 245 | 25 | 270 | 90.74 |
| Observed severe | 59 | 82 | 141 | **58.16** |
| Total | 304 | 107 | 411 | 79.56 |
| **Predicted outcome, 15 non-clinical variables for survival versus decease (no SpO2/FiO2)\*\*\*** |  |  |  |  |
| Classification table | Predicted survival | Predicted decease | Total | % Correctly classified |
| Observed survival | 897 | 9 | 906 | 99.01 |
| Observed decease | 68 | 20 | 88 | **22.73** |
| Total | 965 | 29 | 994 | 92.25 |
| **Predicted outcome, 15 non-clinical variables for severe versus non-severe (no SpO2/FiO2)\*\*\*** |  |  |  |  |
| Classification table | Predicted non-severe | Predicted severe | Total | % Correctly classified |
| Observed non-severe | 691 | 49 | 740 | 93.38 |
| Observed severe | 162 | 92 | 254 | **36.22** |
| Total | 853 | 141 | 994 | 78.77 |

\*The 19 variables included were age, gender, comorbidity index, SpO2/FiO2, Hb, Neutrophil %, Lymph %, monocyte %, eosinophils %, NLR, Platelets, CRP, IL-6, D-dímer, ferritin, fibrinogen, prothrombin time INR, AST/ALT ratio and GFR. \*\* The 16 variables included were the same minus for age, sex, and comorbidities. \*\*\* The 15 variables were the same minus for age, sex, comorbidities and, SpO2/FiO2

Classification tables using different sets of variables show that, the despite good ROC curves and overall high proportion of correctly classified cases, their power in predicting poor outcomes, either decease or severe disease, is under the 60%. Prediction it is very dependent on age and, among biomarkers, on SpO2/FiO2 as seen comparing the different tables using 17 and 16 non-clinical variables. It should be noticed that laboratory variables, even without SpO2/FiO2, are better for predicting severity than decease. When SpO2/FiO2 is excluded (16 variables) the % of correctly classified drops even if the number of observations was increase to the double. Analyses with the reduced set of eight variables i.e., age, comorbidities, SpO2/FiO2, NLR, CRP, AST/ALT, fibrinogen and GFR, gave similar results confirming the redundancy of the variables. For more details see tables in xlsx format, “Multiple logistic regression by decease” and “Multiple logistic regression” in supplementary data.

### Interpretation of biomarkers

The white blood cell differential counts showed marked imbalance due to an approximately 250% reduction in the lymphocyte count and a 20–30% increase in the neutrophil count. At the individual level, the reduction of lymphocytes was deeper than the increase in neutrophils.

The APRs had a broad range of variation e.g., >10,000 and 50-fold for IL-6 and CRP, respectively, and in most patients the values were out of the normal range, while the aspartate aminotransferase/alanine aminotransferase (AST/ALT) ratio and kidney function test results were only moderately altered and often remained within the normal range.

Multivariable logistic regression (Table 3), bivariate age-adjusted logistic regression (Table 4), multiple correlation analyses (Fig 4), and examining their respective shifts from the normal range (Table 6S), suggested that these variables could be classified into three broad categories, clinico-demographic (CD), including age, sex and the comorbidity index; inflammation related biomarkers (IFRB) including blood cell counts, levels of APRs, and coagulation factors; and organ damage-related biomarkers (ODRB), including liver and kidney function tests and SpO_2_/FiO_2_. These analyses revealed that the neutrophil-lymphocyte ratio (NLR) and the AST/ALT ratio captured most of the predictive value of lymphocyte and neutrophils variations and of liver function test variations, respectively, and that SpO_2_/FiO_2_ conveyed much of the predictive power of the ODRBs (see supplementary text “Sequence of statistical biomarker analysis”).

**Fig 4.**
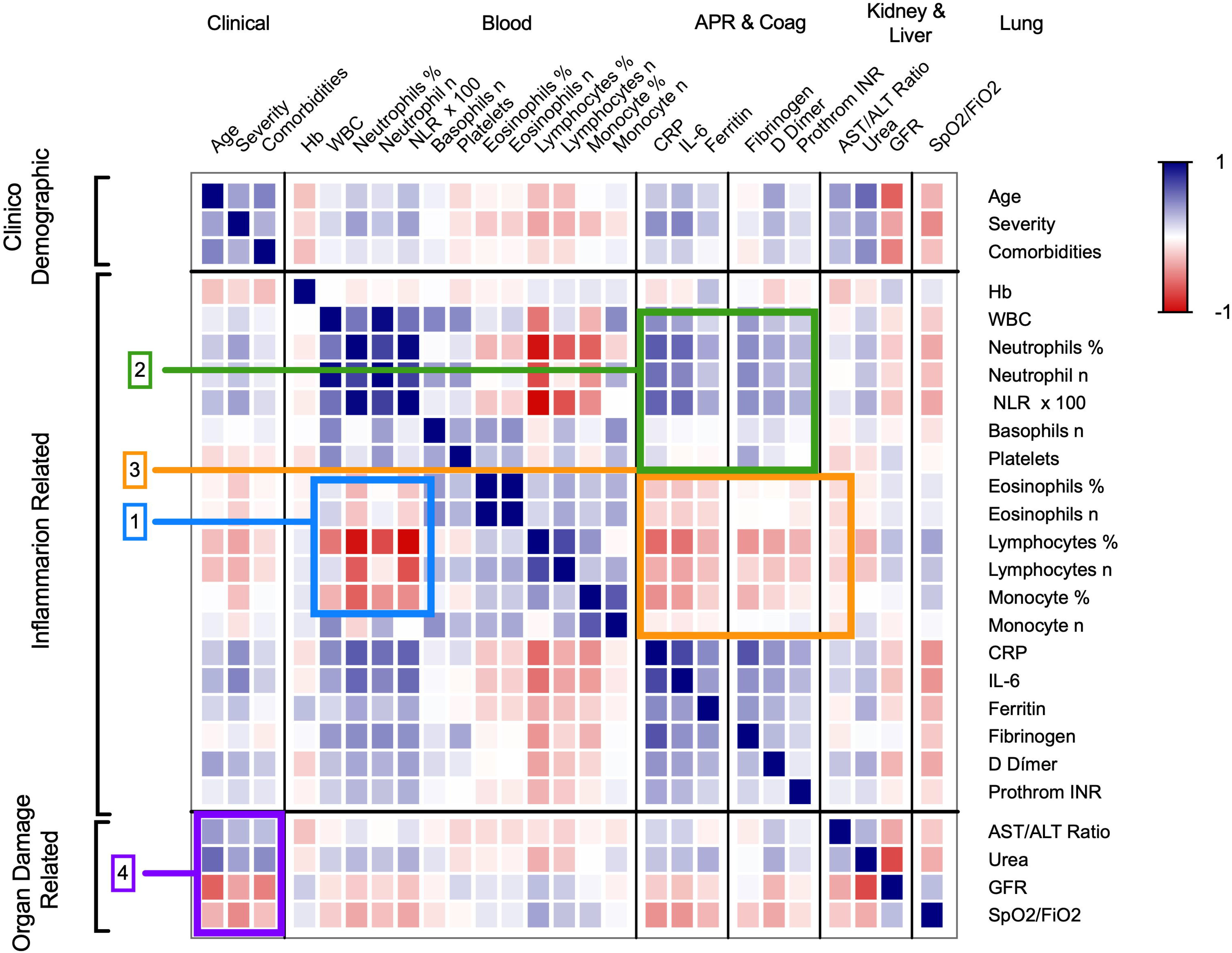
Overall correlograms of selected demographics and clinical laboratory variables organised in categories. [1] The blue rectangle highlights the negative correlation between neutrophils and the cluster of lymphocytes, monocytes, and eosinophils. [2] The green rectangle highlights the blood cell variables that correlate positively with the acute-phase reactants (APRs) and coagulation factors. [3] The orange rectangle highlights the negative correlation between lymphocytes, monocytes, and eosinophils with APRs and coagulation factors. [4] The magenta rectangle highlights the correlations of age, disease severity and comorbidities with liver, kidney and lung function variables. The cells following the diagonal highlights the seven families of variables: demographics/clinical, myeloid cells, lymphocytes/mononuclear cells, APRs, coagulation, liver function test, and kidney function test, which show the expected strong correlations among themselves. The thick lines between columns separate the main categories. APR, acute-phase reactants; SpO_2_/FiO_2_, oxygen saturation/fraction of inspired oxygen; NLR, neutrophil-to-lymphocyte ratio; CRP, C-reactive protein; AST, aspartate aminotransferase; ALT, alanine aminotransferase; GFR, glomerular filtration rate.

**Table 4.** Age adjusted logistic regression for 28-days survival/decease and non-severe/severe outcomes.

| 28d decease outcome age adjusted |  |  |  |  |  | 28 maximal severity outcome age adjusted |  |  |  |  |
| --- | --- | --- | --- | --- | --- | --- | --- | --- | --- | --- |
| Variables | Odds Ratio | CI | Z value | Pr(>\|z\|) | Z value Rank | Odds Ratio | CI | Z value | Pr(>\|z\|) | Z value Rank |
| IL-6 (pg/mL) | 2.59 | 2.06-3.31 | 7.84 | <0.001 | 1 | 12.24 | 8.24-18.56 | 12.10 | <0.001 | 1 |
| CRP (mg/dL) | 3.20 | 2.38-4.41 | 7.40 | <0.001 | 2 | 9.55 | 6.47-14.40 | 11.06 | <0.001 | 2 |
| SpO2/FiO2 ratio | 0.99 | 0.99-1.00 | 7.05 | <0.001 | 3 | 0.99 | 0.98-0.99 | 10.17 | 0.001 | 4 |
| Neutrophils (%) | 1.06 | 1.04-1.08 | 6.38 | <0.001 | 4 | 1.19 | 1.14-1.23 | 10.76 | <0.001 | 3 |
| NLR x 100 | 1.08 | 1.06-1.11 | 6.33 | <0.001 | 5 | 1.13 | 1.10--1.16 | 9.87 | 0.001 | 5 |
| Monocytes (%) | 0.86 | 0.81-0.90 | 6.20 | <0.001 | 6 | 0.86 | 0.83-0.89 | 7.89 | <0.001 | 8 |
| Neutrophils (10<sup>9</sup>/L) | 1.14 | 1.09-1.19 | 5.74 | <0.001 | 7 | 1.19 | 1.14-1.23 | 8.86 | <0.001 | 7 |
| GFR (mL/1.73 m<sup>2</sup>) | 0.98 | 0.97-0.99 | 5.44 | <0.001 | 8 | 0.98 | 0.97-0.98 | 6.94 | <0.001 | 11 |
| Lymphocytes (%) | 0.95 | 0.92-0.97 | 5.04 | <0.001 | 9 | 1.03 | 1.03-1.04 | 9.79 | <0.001 | 6 |
| WBC (10<sup>9</sup>/L) | 1.11 | 1.06-1.16 | 4.82 | <0.001 | 10 | 1.14 | 1.01-1.18 | 7.29 | <0.001 | 9 |
| AST/ALT ratio | 1.49 | 1.26-176 | 4.74 | <0.001 | 11 | 1.53 | 1.32-1.80 | 5.47 | <0.002 | 13 |
| D dimer (ng/mL) | 1.51 | 1.26-1.83 | 4.35 | <0.001 | 12 | 2.60 | 1.92-3.56 | 6.05 | <0.001 | 12 |
| Creatinine (mg/dL) | 1.32 | 1.14-1.53 | 3.68 | <0.001 | 13 | 1.39 | 1.20-1.65 | 4.02 | <0.001 | 16 |
| Lymphocytes (10<sup>9</sup>/L) | 0.51 | 0.35-0.73 | 3.61 | <0.001 | 14 | 0.41 | 0.32-0.52 | 6.95 | <0.001 | 10 |
| Ferritin (ng/mL) | 1.43 | 1.12-1.85 | 2.85 | 0.004 | 15 | 1.00 | 1.000-1.001 | 5.42 | <0.001 | 14 |
| Eosinophils (%) | 0.63 | 0.45-0.85 | 2.79 | 0.005 | 16 | 0.62 | 0.49-0.76 | 4.35 | <0.001 | 15 |
| Hb (g/dL) | 0.89 | 0.81-0.98 | 2.41 | 0.016 | 17 | 1.12 | 1.04-1.19 | 3.23 | 0.001 | 17 |
| Eosinophils (10<sup>9</sup>/L) | 0.02 | 0.00-0.62 | 2.07 | 0.039 | 18 | 0.04 | 0.003-0.360 | 2.67 | 0.007 | 18 |
| Monocytes (10<sup>9</sup>/L) | 0.72 | 0.45-1.07 | NC | 0.127 | NA | 0.74 | 0.51-1.04 | 1.66 | 0.097 | 20 |
| Platelets (10<sup>9</sup>/L) | 1.00 | 1.00-1.00 | NC | 0.203 | NA | 1.00 | 0.997-1.000 | 1.80 | 0.070 | 19 |

AST, Aspartic Amino Transferase; ALT, Alanine Amino Transferase; CRP; C Reactive Protein; GFR, Glomerular Filtration Rate; Hb, Haemoglobin; NLR, Neutrophil Lymphocyte Ratio; NA, not calculated, WBC, White Blood cell Count. The results are sorted by the value of Z.

Applying this classification to assess variable performance using ROC curve analysis (Table 5) showed that the CD and ODRB variables performed moderately better for predicting survival, while the IFRB variables were better for predicting disease severity and in distinguishing between moderate and severe disease in the four-group classification (Table 3 and Fig 5A). The strong influence of age was more evident in the analysis of survival curves (Fig 5S) using Youden index as cut-off, (Table 5). Furthermore, the hazard ratio (HR) for age under or above 60 years was 32, while the next highest HR was 9.3 for GFR. The predictors of disease severity in descending order were age, GFR, urea, IL-6, D-dimer, and comorbidities (Fig 5B). The predictive power of both the ODRB and IFRB variables was maintained in the logistic regression analysis after adjusting for age (Table 4) and in the ROC analysis of patients stratified by age interval (Table 7S), two approaches that reduced the influence of age. However, the random forest simulation further confirmed that age was the single best predictor of outcome, and that the combination of all variables was only partially additive (Table 8S).

**Fig 5.**
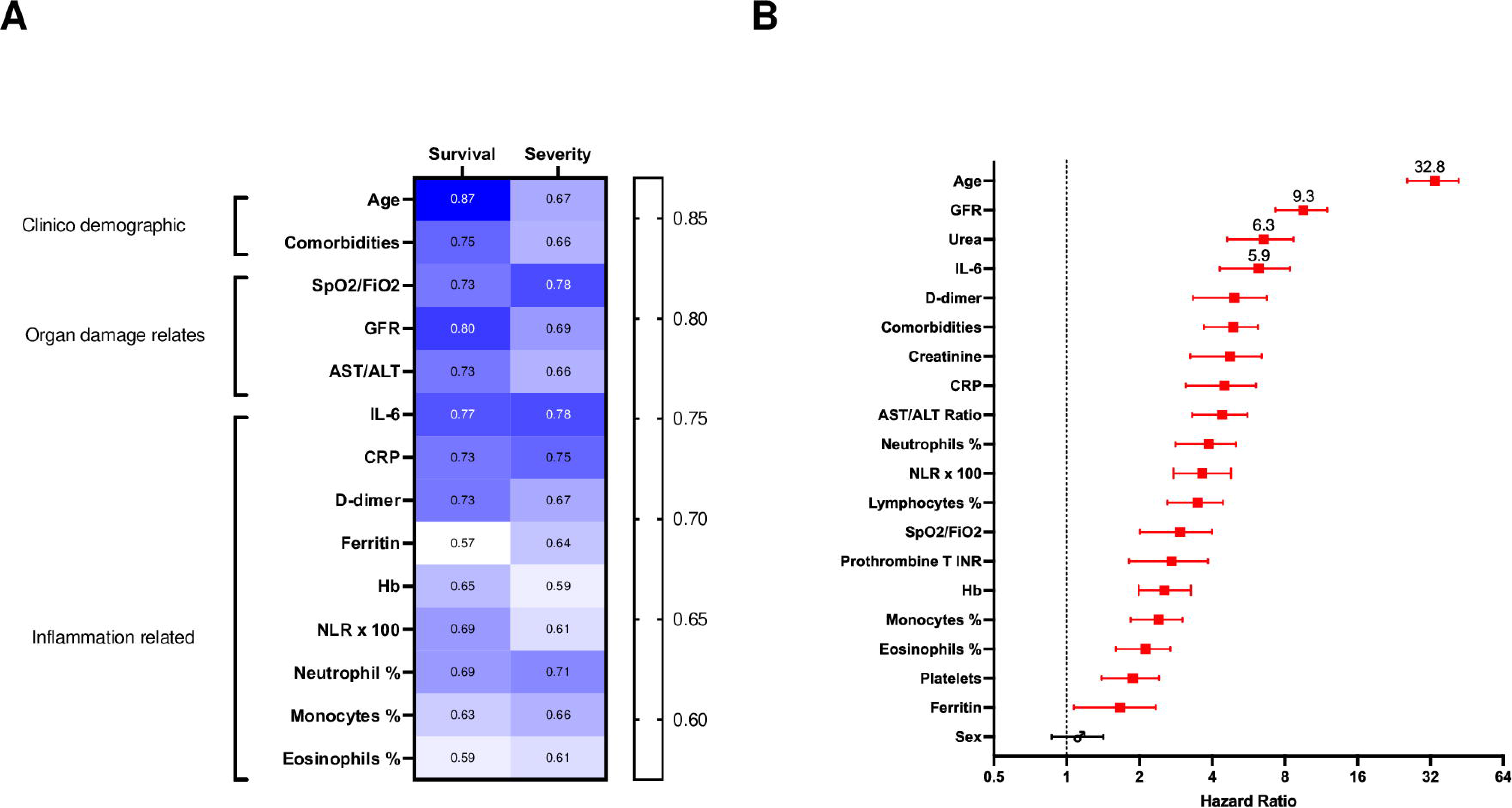
Relative weight of different variables in outcome prediction. (A) Heatmap summarising the values under the curve (AUC) of receiver operator curves (ROC) generated for to each of the main variables; the performance was assessed by survival/decease and for non-severity/severity outcomes. (B) Hazard ratios corresponding to survival curves for Youden index cut-off. Red, significant values. SpO2/FiO2, oxygen saturation/fraction of inspired oxygen; GFR, glomerular filtration rate, NLR, neutrophil-to-lymphocyte ratio; CRP, C-reactive protein; AST, aspartate aminotransferase; ALT, alanine aminotransferase; Hb, haemoglobin. The r- and p-values of the data represented in the heatmap are in xlsx format files in the supplementary material “Correlation of variables, r-values” and “Correlation of variables, p-values.

**Table 5.** ROC curve analysis as for clinical laboratory test performance comparison for survival/decease and non-severe/severe outcomes.

| **Decease Outcome** |  |  |  |  |
| --- | --- | --- | --- | --- |
| **Variable** | **AUC** | **CI** | ***p*** | **YOUDEN for**<br>**Hazard Ratios cut off** |
| Age | 0.87 | 0.85-0.89 | 0.001 | >60.50 |
| Comorbidities | 0.75 | 0.72 -0.78 | 5.95E-25 | >1.50 |
| GFR | 0.80 | 0.77-0.83 | 2.45E-53 | < 87.30 |
| IL-6 | 0.77 | 0.73-0.81 | 5.24E-26 | > 50.50 |
| AST/ALT | 0.73 | 0.69-0.77 | 6.08E-31 | > 1.64 |
| SpO2/FiO2 | 0.73 | 0.70-0.78 | 1.79E-43 | < 439.50 |
| CRP | 0.73 | 0.70-0.78 | 1.44E-20 | >11.13 |
| D-dimer | 0.73 | 0.69-0.77 | 7.42E-20 | > 353.00 |
| Creatinine | 0.71 | 0.67-0.74 | 4.35E-25 | > 1.12 |
| Lymphocyte % | 0.69 | 0.65-0.73 | 1.18E-21 | < 12.05 |
| NLR | 0.69 | 0.65-0.73 | 1.33E-21 | >6.85 |
| Neutrophil % | 0.69 | 0.65-0.73 | 1.61E-43 | > 82.15 |
| Hb | 0.65 | 0.62-0.69 | 5.67E-09 | < 13.45 |
| Monocytes | 0.63 | 0.59-0.67 | 5.05E-11 | < 6.65 |
| Prothrombin time (INR) | 0.61 | 0.56-0.65 | 1.37E-07 | > 1.21 |
| Platelets | 0.60 | 0.56-0.64 | 9.94E-07 | < 162.50 |
| Eosinophils % | 0.59 | 0.55-0.62 | 1.32E-5 | < 0.15 |
| Ferritin | 0.57 | 0.51-0.62 | 0.019 | > 668.00 |
| **Severity outcome** |  |  |  |  |
| IL6 | 0.78 | 0.75-0.80 | 8.65E-56 | NC |
| SpO2/FiO2 | 0.77 | 0.74-0.81 | 1.79E-43 | NC |
| CRP | 0.75 | 0.71-0.77 | 5.77E-43 | NC |
| NLR | 0.71 | 0.68-0.73 | 1.74E-41 | NC |
| GFR | 0.69 | 0.65-0.71 | 1.69E-33 | NC |
| Age | 0.67 | 0.64-0.70 | 2.55E-29 | NC |
| D-dimer | 0.67 | 0.63-0.69 | 2.63E-20 | NC |
| Monocytes % | 0.66 | 0.62-0.68 | 1.92E-24 | NC |
| AST/ALT | 0.66 | 0.62-0.68 | 2.97E-24 | NC |
| Comorbidity | 0.65 | 0.62-0.68 | 5.95E-25 | NC |
| Ferritin | 0.64 | 0.60-0.67 | 1.08E-12 | NC |
| Eosinophils % | 0.61 | 0.57-0.63 | 3.14E-12 | NC |
| Hb | 0.59 | 0.55-0.62 | 5.67E-09 | NC |
| Monocytes n | 0.57 | 0.53-0.59 | 1.02E-05 | NC |

AST, Aspartic Amino Transferase; ALT, Alanine Amino Transferase; CRP; C Reactive Protein; GFR, Glomerular Filtration Rate; Hb, Hemoglobin; NLR, Neutrophil Lymphocyte Ratio; NC, not calculated, because Youden index was applied only for survivals vs decease.

### Predictive power of variables during hospitalisation

The analysis of the 7,586-follow-up observations showed that association of biomarkers with survival varied during the 28 days of follow-up. The time course curves of the average clinical laboratory variables for survivors and deceased remained apart during the first few days of hospitalisation with maximum separation around day 5 (Fig 6). Interpretation of these values in patients with longer hospital stays was difficult due to the decreasing sample size and complications arising from medical interventions. The survival curves for ODRBs, GFRs and AST/ALT ratio maintained their separation for most of the follow-up period while IFRB tended to merge at the third week of follow-up.

**Fig 6.**
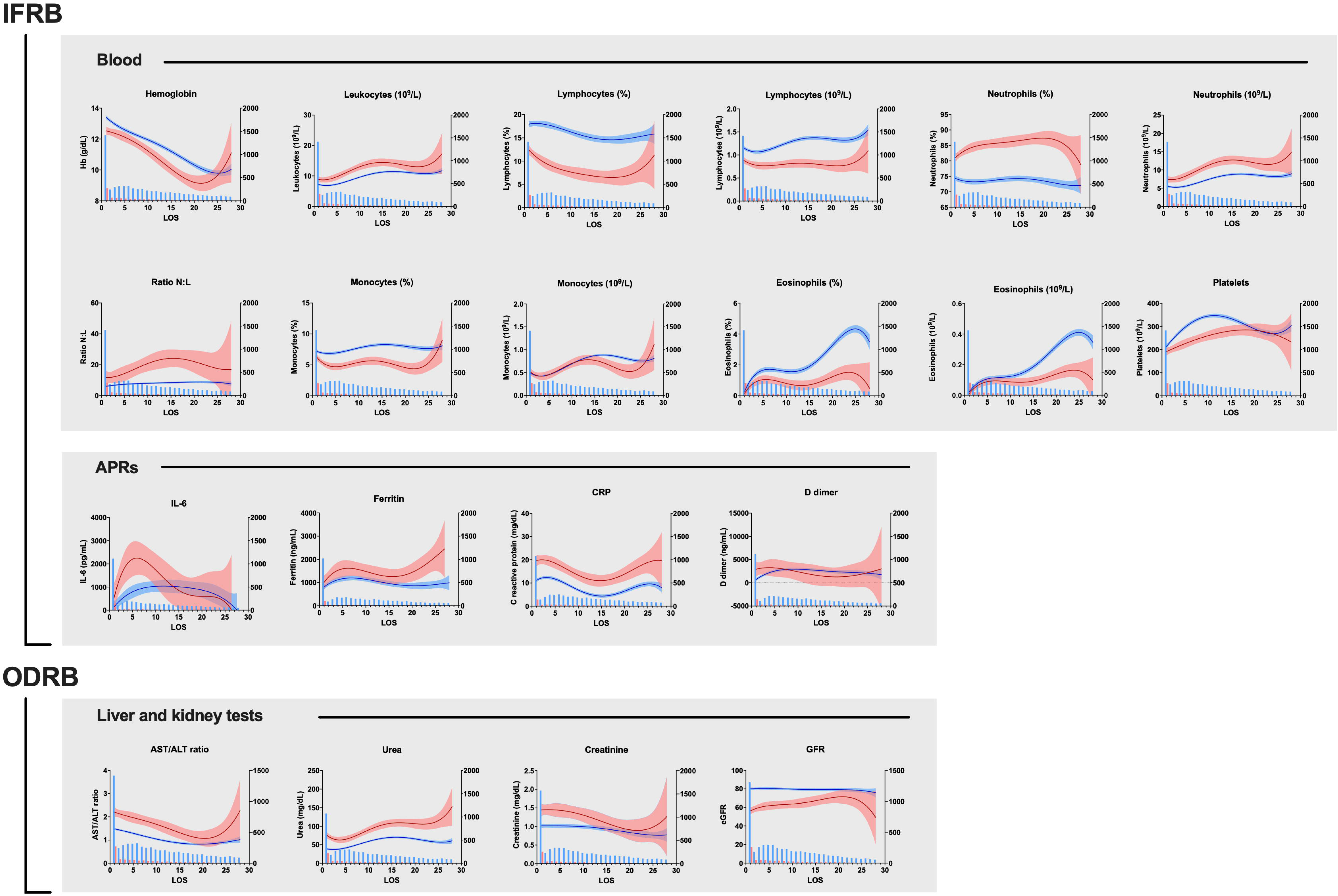
Vall d’Hebron University Hospital cohort, variations in the average clinical laboratory variables during the 28-day follow-up period. The blue and red lines represent the mean ± CI values of the corresponding parameter each day of follow-up for the survivors and deceased respectively. The blue bars indicate the number of values available for each day. Notice that samples were not obtained every day and therefore the averages result from plotting together all available values for each day of follow-up, as in Abers et al.[1] Data correspond to 7,586 samples, 6,589 from survivors and 997 from deceased out of 1,079 patients of the HUVH cohort. NLR, neutrophil-to-lymphocyte ratio; CRP, C-reactive protein; AST, aspartate aminotransferase; ALT, alanine aminotransferase; GFR, glomerular filtration rate; IFRB, inflammation-related biomarkers, ODBRs, organ damage-related biomarkers. APRs, acute-phase reactants.

### Selection of a core panel of clinical laboratory tests

At present in HUVH, as in many hospitals, approximately 30 clinical laboratory variables plus SpO2/FiO2 are routinely measured in COVID-19 patients as part of the work-up on admission. Correlation analysis and multivariable logistic regression showed that these variables had a high level of multicollinearity (Table 3), which was confirmed by random forest simulation and PCA (Fig 4 and 4S, Table 8S). Using repeated analysis and progressively excluding variables, a reduced set of eight variables: age, comorbidity index, SpO2/FiO2, haemoglobin, NLR, CRP, AST/ALT ratio, and GFR, were found to capture the prediction power of all variables (see supplementary material, “Sequence of statistical biomarkers analyses: complexity reduction” and tables “Repeated multivariable logistic regression deceased” and “Repeated multivariable logistic binary severity” Supplementary Excel tables). As age and comorbidities are non-time-varying, only six of the eight variables are really useful for clinical management. These results do not imply, however, that IL-6, ferritin, lactate dehydrogenase, triglycerides, procalcitonin, D-dimer, troponin and coagulation tests do not provide valuable information in clinical practice depending on the context.

### Results of the two validation cohort analyses

The comparison among the three cohorts confirmed the prognostic power of the main IFRB and ODRB variables (Table 6), even though the statistical ranking of their positions in logistical regression and ROC curves varied between cohorts (Fig 7 and 8) and in the pairwise analyses of association there was concordance in 21 of 23 variables, with discordance for fibrinogen and AST, see Figure 6S. In addition, biomarker performance as predictors of outcome was maintained in the three cohorts in the random forest simulations (Table 9S).

**Fig 7.**
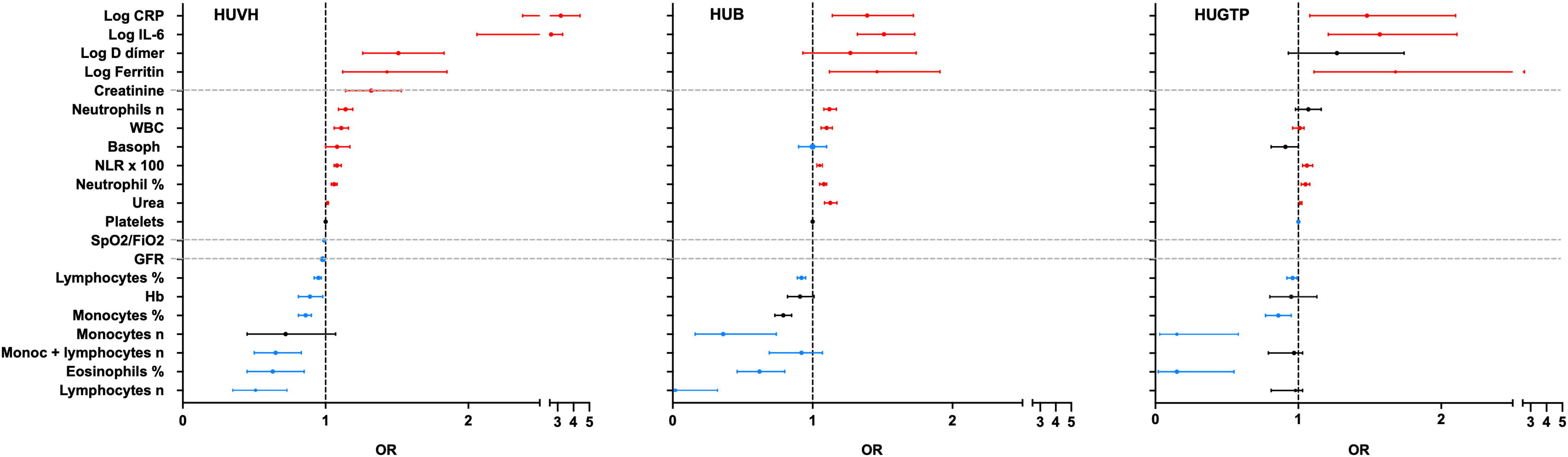
Multivariable logistic regression analysis, age-adjusted, for the main variables of the three hospital cohorts. The three forest plots show how, after correcting for age, acute phase reactants and other inflammatory biomarkers have a similar -but bot identical-ranking in the three cohorts. The horizontal whiskers represent the 95% confidence intervals; values in red indicate positive predictive and blue negative predictive effect on the 28-day survival/deceased as outcome. These graphs are for comparing the OR rankings among the different hospital cohorts, and not for comparing the weight of the variables within a cohort, as the ORs are derived from variables that use different units and ranges of variation. set; HUVH, Vall d’Hebron University Hospital; HUB, Hospital Universitari Bellvitge, HUGTP, Hospital Universitari Germans Trias Pujol. AST, aspartate aminotransferase; ALT, alanine aminotransferase; CRP, C-reactive protein; GFR, glomerular filtration rate; Hb, haemoglobi; IL, interleukin; LDH, lactate dehydrogenase and NLR, neutrophil-to-lymphocyte ratio.

**Fig 8.**
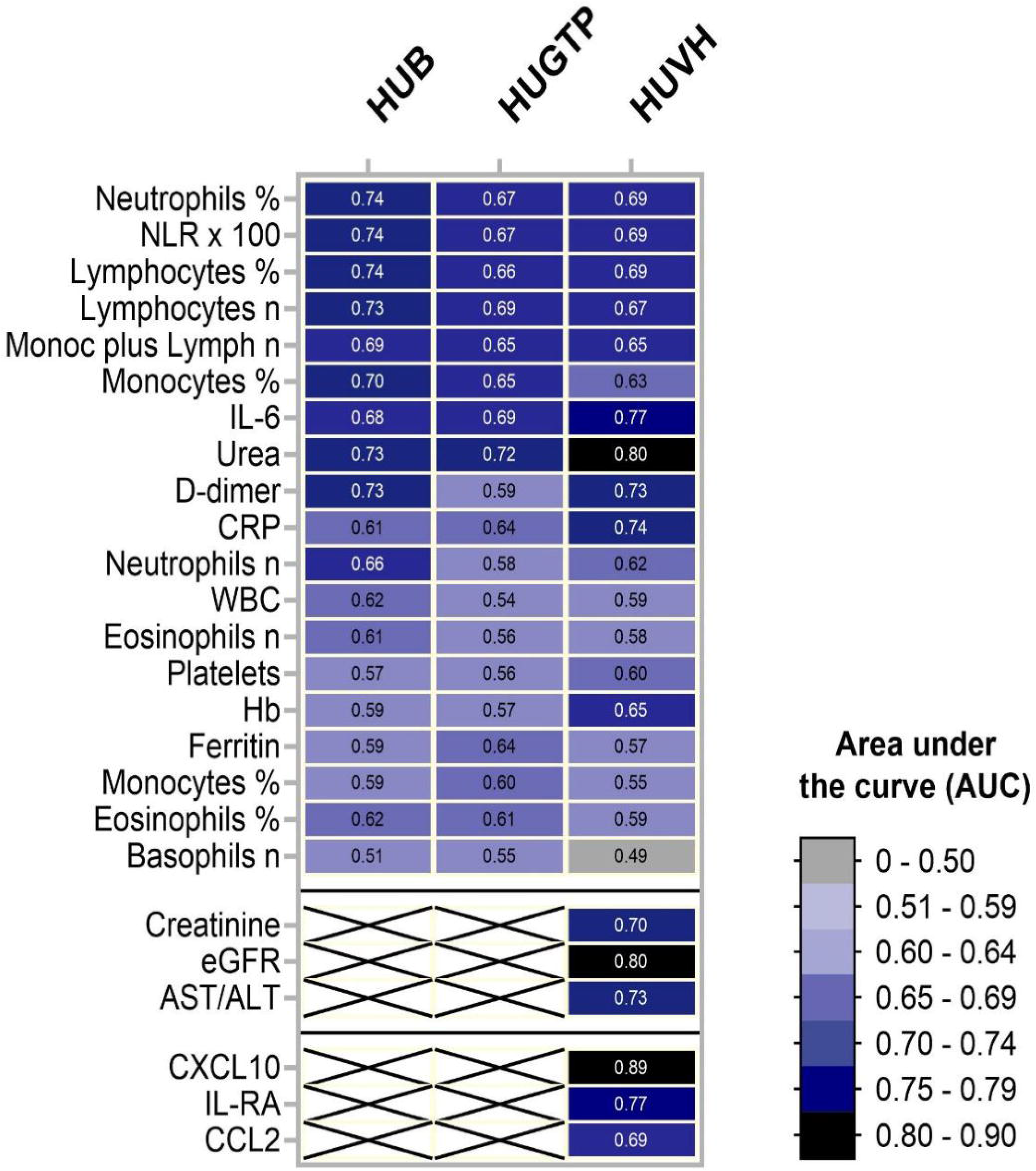
Heatmap of the area under the curve (AUC) of the receiver-operating characteristic curves corresponding to the variables available for the three cohorts. IL-6, CRP, urea, lymphocytes, and neutrophils occupy central positions. At the bottom, the AUC for some variables available only from the HUVH cohort and the AUC values for the three cytokines that perform better in the group of 74 patients who were analysed in the HUVH cohort. The numbers within the cells are the AUC values. APR, acute-phase reactants; Sa/Fi, oxygen saturation/fraction of inspired oxygen; DFSO, days from symptom onset; HUVH, Hospital Universitary Vall d?Hebron; LOS, length of stay; NLR, neutrophil-to-lymphocyte ratio; CRP, C-reactive protein; AST, aspartate aminotransferase; ALT, alanine aminotransferase; GFR, estimated glomerular filtration rate; IL, interleukin; LDH, lactate dehydrogenase; Hb, haemoglobin; ROC, receiver-operating characteristic; AUC, area under the curve. The variables only available in HUVH have been plotted for comparison; the dotted horizontal lines are to help in visual comparisons and highlight these missing variables.

**Table 6.**
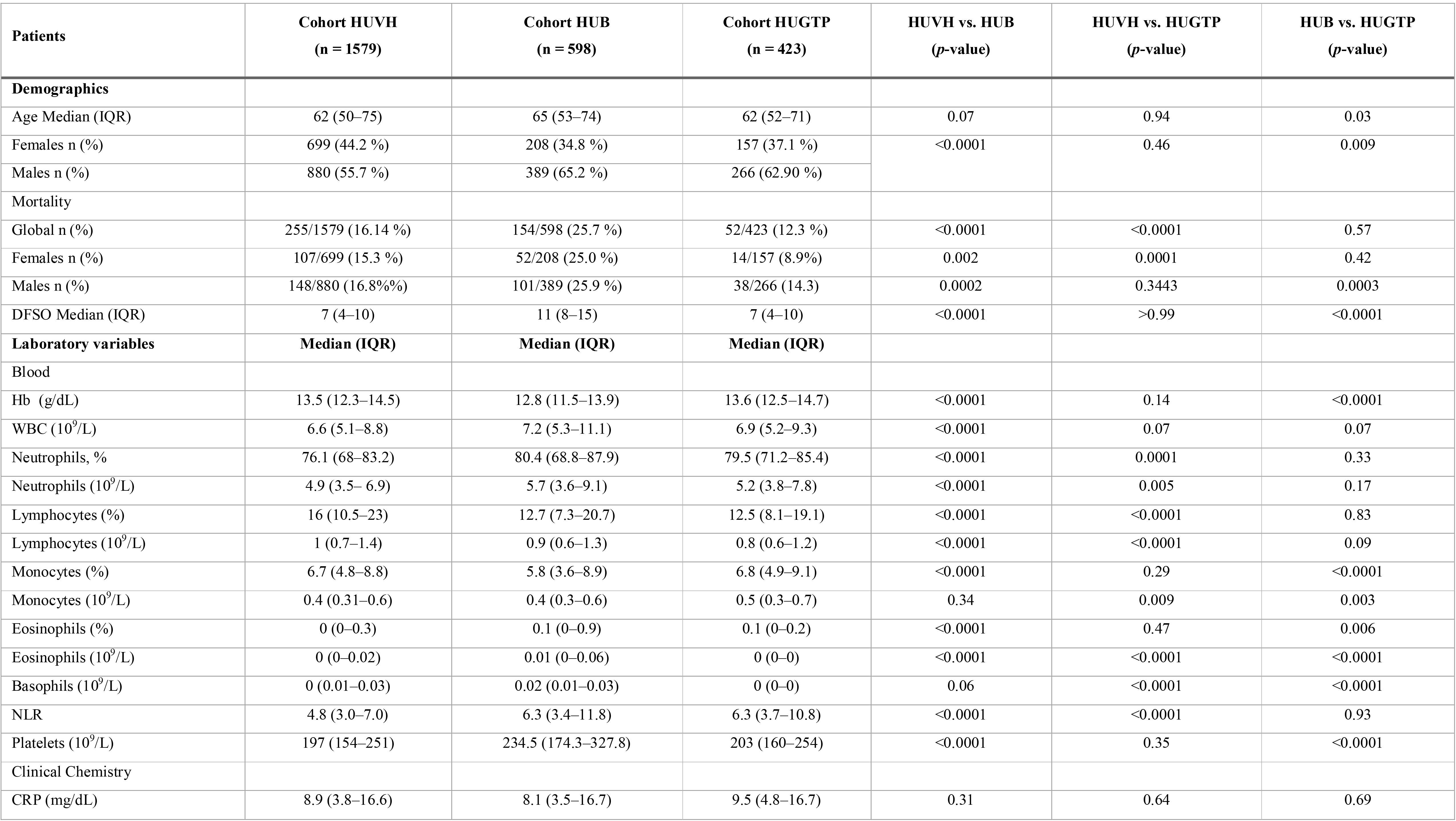
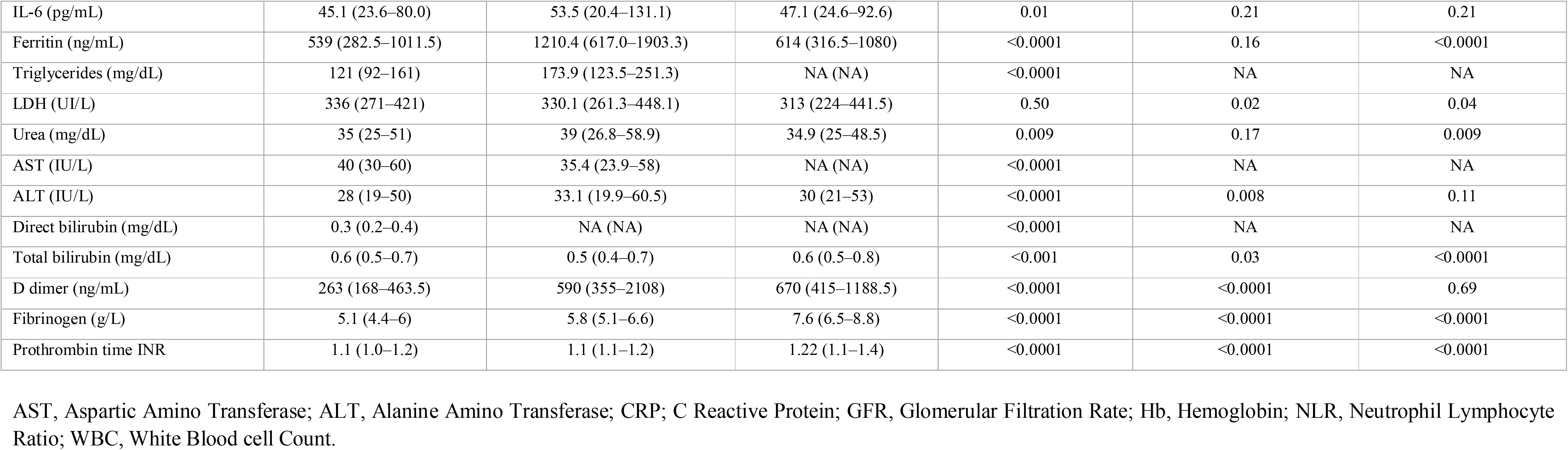
Pairwise comparison of demographic and clinical laboratory biomarkers in the exploratory (HUVH) and the two validation cohorts (HUGTP and HUB).

### Predictive ability of immunological variables

Despite the limited size of the group analysed in the pilot study (n=74, Table 10S), CXCL10 had the highest ROC curve (AUC=0.83) of all variables including age, IFRB and ODRB, and performed better than any of the other variables considered. IL1RA and CCL2 also showed promise as biomarkers (Table 7 and Fig 9). The immune phenotype was analysed in 41 patients (Table 10S). There was a steep reduction in the size of all T-cell subsets, which was more marked for CD8 effector and memory cells, and an increase in activation markers that was similar to the pattern observed in other time-series analyses [24, 37], revealing a deep disturbance of the immune response in severely ill patients (see Expanded phenotype analysis in supplementary) (Figs 10, 11S and 12S). No single subset of lymphocytes emerged as a biomarker from this analysis, but the pattern of the subsets within every major subpopulation was distinct and suggested that, in a larger study, it should be possible to identify patterns with prognostic value, as already proposed[26,29,38].

**Fig 9.**
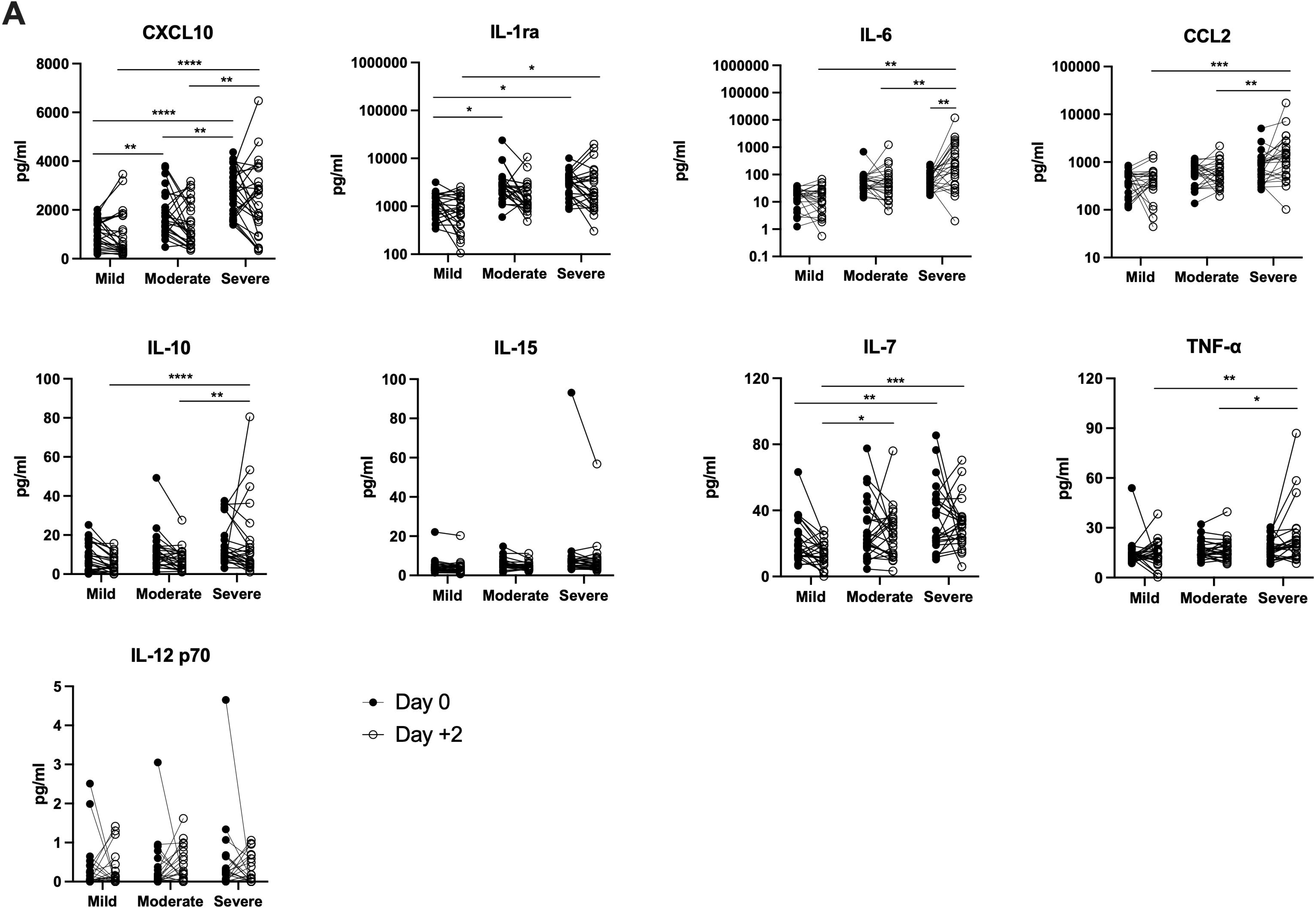
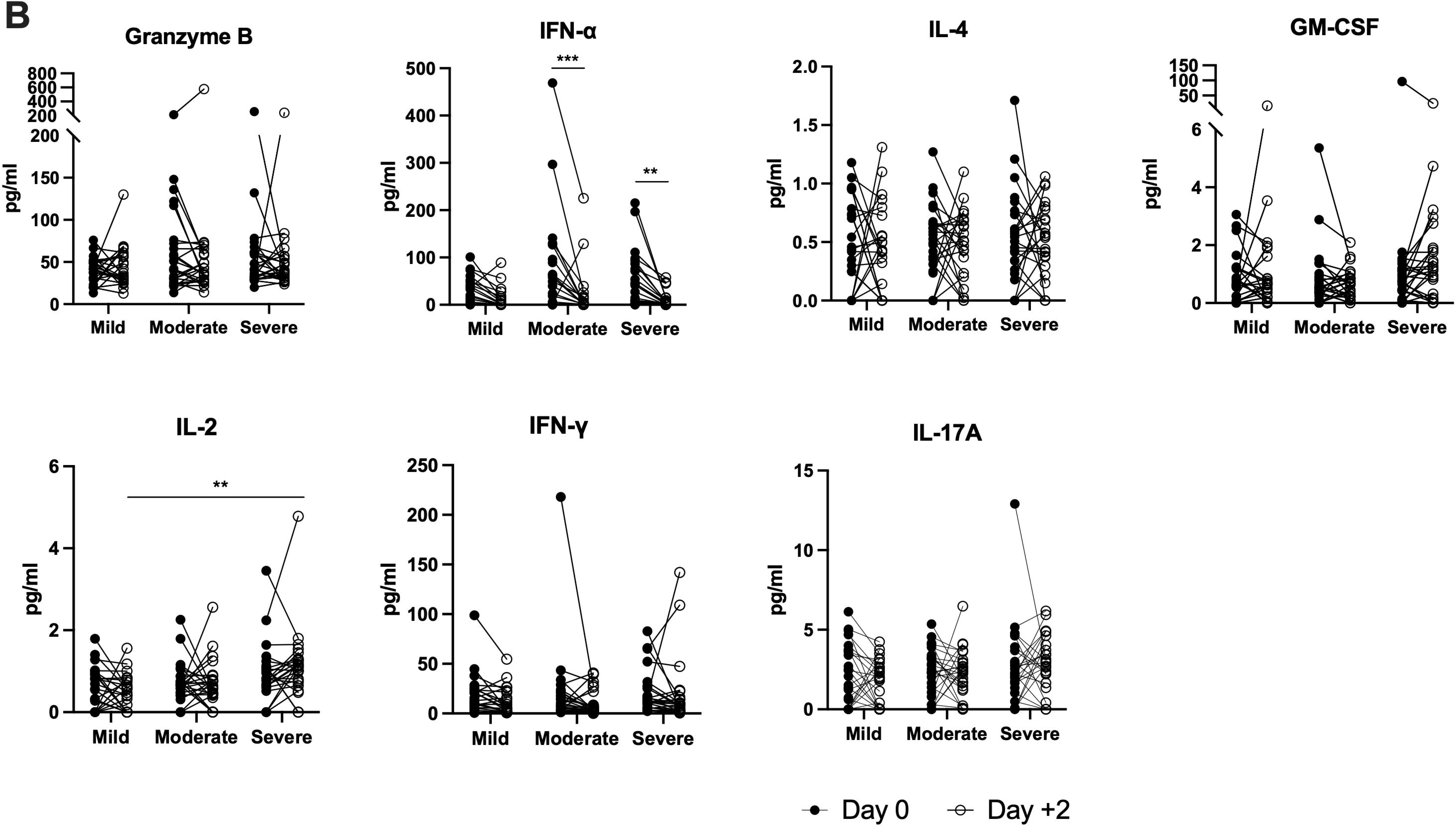
Levels of cytokines and related factors in the Hospital Universitary Vall d?Hebron cytokine studies sub-cohort. The levels of cytokines were measured in the ELLA® platform cytokines on days 0 and +2, and the changes in the levels are shown as before/after graphs. (A) cytokines mediating innate immunity and (B) IFN-alpha plus cytokines mediating adaptive immunity.

**Fig 10.**
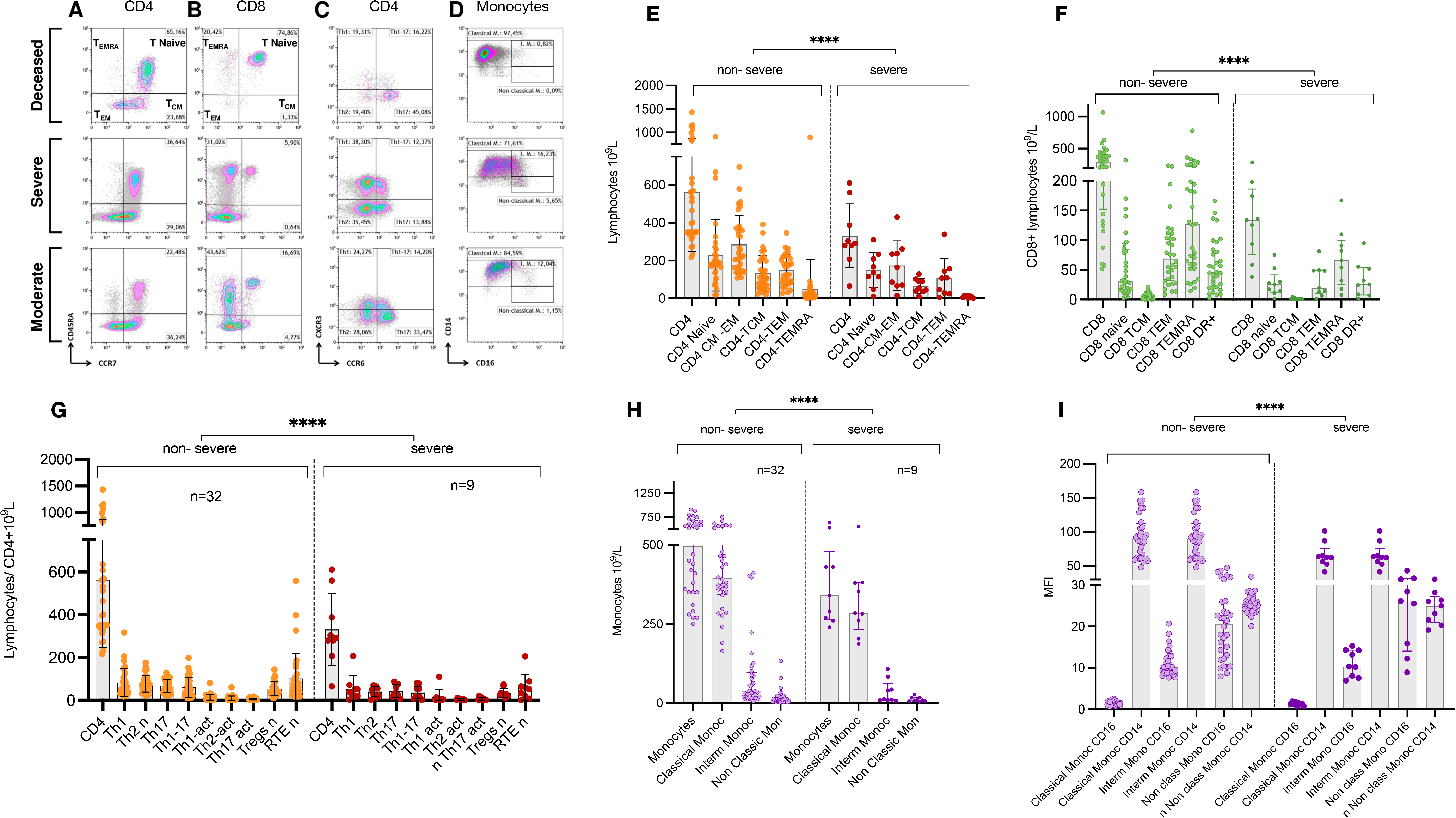
Representative flow cytometry plots from the Vall d’Hebron University Hospital sub-cohort. A, CD4 and B, CD8 T lymphocyte subpopulations distributed by phenotypes based on CD45RA and CCR7. C, CD4 T lymphocyte Th-polarisation by CXCR3 and CCR6 expression. D, Monocyte subpopulations (classical, intermediate monocytes [IM] and non-classical monocytes) in a comparison of patients belonging to the deceased, severe, and moderate patient categories. E, Distribution of CD4 naïve and memory subsets among non-severe and severe patients. F, Distribution of CD8 naïve and memory subsets among non-severe and severe patients; G, Distribution of CD4 Th polarized subsets among non-severe and severe patients; H Distribution of monocytes subsets among non-severe and severe patients; I, Mean Fluorescent Intensity (MFI) of CD14 and CD16 in the different monocyte subsets among non-severe and severe patients. Non severe patients n=32 and severe patients n=9, for all plots; **** p<0.0001 by non-parametric FDR corrected Kruskal-Wallis test.

**Table 7.**
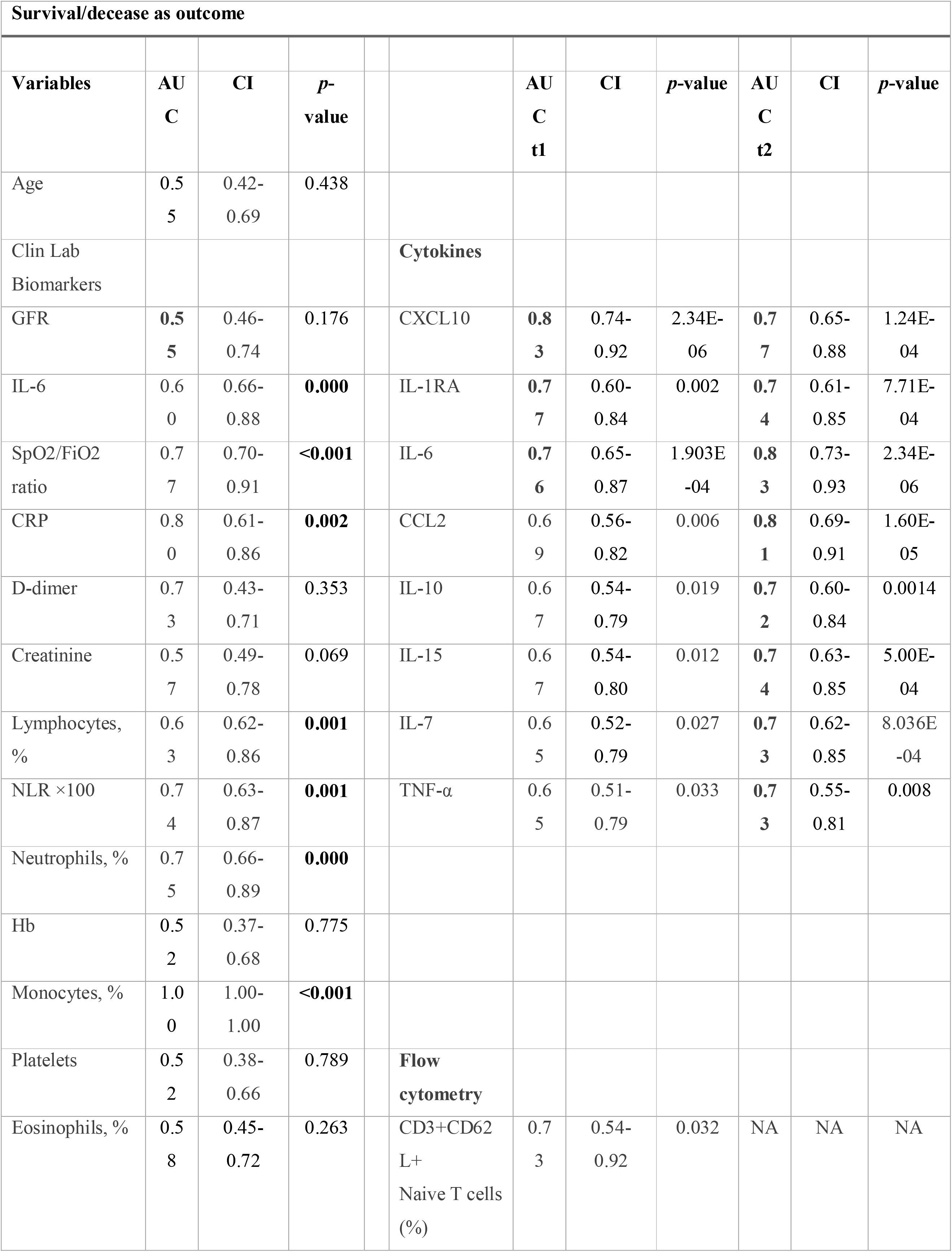
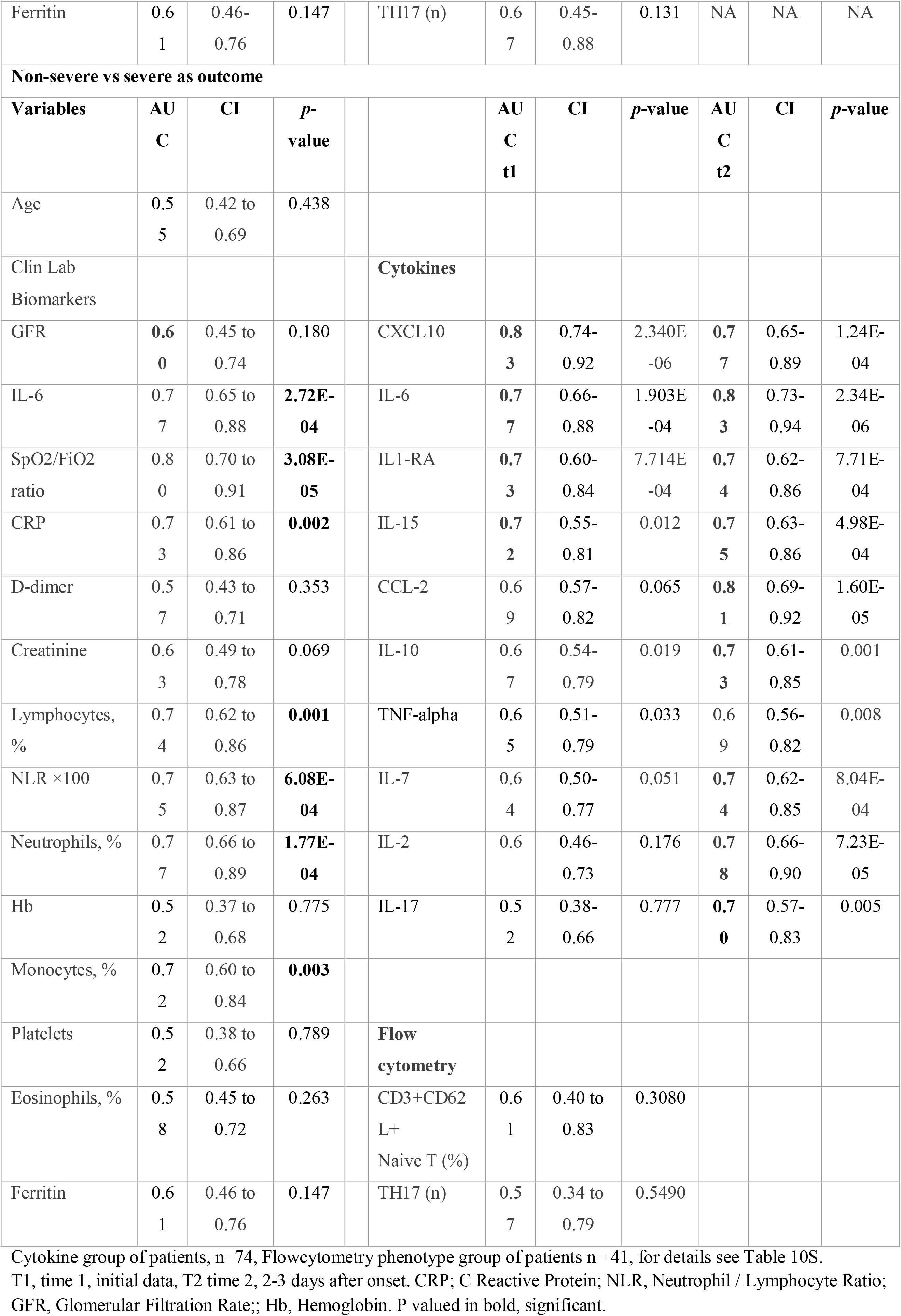
Performance of expanded immunological parameters in the special immunological studies group as assessed by ROC curve analysis and compared with other variables in the same group.

## DISCUSSION

The analyses revealed the limitations of currently used clinical laboratory tests used to assess the prognosis of patients with COVID-19 and tried to improve their interpretation by grouping them into categories that reflect the two main biological processes that are measured, i.e., inflammation and organ damage. As their limitations are due to redundancy, clinical management protocols could be simplified, but additional biomarkers with independent predictive power are urgently needed. This study highlights the lack of tests, for early prediction of the specific immune response to SARS-CoV-2. Such tests could provide critical non-redundant information required for prediction and clinical management. The results of the pilot study using a selection of robust immunological techniques derived from other areas of clinical immunology, suggest that better tests can be identified through systematic investigation.

As well as this central message, other notable findings are: 1) The three cohorts confirmed the strong association of: SpO_2_/FiO_2_, neutrophilia, lymphopenia, APRs, coagulation factors, kidney function and the AST/ALT ratio with survival and predicting disease severity. 2) There was a high level of collinearity (redundancy) among the different variables, which explains the limited predictive ability of current tests. 3) After reducing redundancy, the best combination of variables was age, comorbidity index, SpO_2_/FiO_2_, NLR, CRP, AST/ALT ratio, fibrinogen, and GFR. 4) The classification of biomarkers into inflammation and organ damage related helped with their interpretation and revealed that organ damage tests are better predictors of survival than severity, and that inflammation parameters are better predictors of severity than survival. 5) For the clinician at the bedside, some changes in GFR and liver test may be less conspicuous than CRP and IL-6 but they may deserve more attention.

There are several limitations to this study, including its retrospective nature. Another limitation is the absence of information regarding two key factors: the SARS-CoV-2 viral load and markers of the adaptive immune response. The SARS-CoV-2 detection techniques used during this period were not quantitative, and the variability of sampling efficiency reduces their value, even with current improved measurement methods. Serological markers need 7–21 days to become detectable and are not very helpful as a tool to predict the prognosis of the patients during the initial medical assessment [39]. Finally, the effect of treatment on mortality, which changed continuously during the first wave, was not analysed in this study. We did not strictly follow the transparent reporting of a multivariable prediction model for individual prognosis or diagnosis recommendations (TRIPOD), because generating a prediction mode was not an objective, but most requirements were fulfilled [38].

The analyses presented here are intended for improved interpretation of available biomarkers, but no algorithm is proposed. Most algorithms with good predictive power include parameters, such as oxygen requirements and imaging data, that reflect organ damage in patients that are already on the path to severe disease [14–22,40]. The ideal algorithm/biomarker should be able to identify patients at risk before organ damage occurs. Our results suggest that this is difficult with current tests because inflammation and organ damage biomarkers are strongly correlated at the time the patients reach the emergency department. If, as postulated, the main determinant of severity is a pre-existing latent pro-inflammatory state that leads to a late and inefficient adaptive immune response, biomarkers of this basal inflammation and inefficient response should be identifiable; if the generation of specific cytotoxic T lymphocytes is the main defence mechanism against an acute respiratory infection to a novel virus such as SARS-CoV-2, the early monitoring of these cells would help to predict the patient outcome [1,24,28,29,41–43]. These are the two obvious approaches to generate better biomarkers and the corresponding tests. Reliable early biomarkers would reduce the rate of hospitalization and as new treatments that are becoming available require early administration, generation of such biomarkers is urgent and should be feasible.

### Data sharing

The supplementary material contains detailed information on the statistical analysis but deidentified data tables will shared on request after approval of a proposal, with a signed data access agreement.

## Supporting information

SEQUENCE OF STATISTICAL BIOMARKERS ANALYSES

Correlation r values

Correlation p values

Repeated Multiple Logistic Regression non-severe vs severe

Repeated Multiple Logistic Regression survivors vs non survivors

Repeated Multiple Logistic Regression for follow-up

## Data Availability

All data produced in the present study are available upon reasonable request to the authors

## Acknowledgments

The following are members of the “Hospital Vall d’Hebron Group for the study of COVID-19 immune profile”:

Artur Llobell Uriel MD^1^, Romina Dieli MD^1^, PhD, Roger Colobran-Oriol PhD^1,2,3^, Gemma Codina MD, PhD^4^, Tomás Pumarola MD, PhD^4^, Roser Ferrer PhD^5^, Vicente Cortina BSc^5^, Magda Campins MD, PhD^6^, Isabel Ruiz MD^7^, Nuria Fernández MD^7^, Esteban Ribera MD^7^, Joan Roig MD^7^, Ricardo Ferrer MD^8.9^, Adolfo Ruiz-Sanmartín MD^8,9^, Albert Selva MD, PhD^10.11^, Moisés Labrador MD, PhD^10,11^, María José Soler Romeo MD, PhD^12^, Jaume Ferrer MD, PhD^13^, Eva Polverino MD, PhD^13^ Antonio Alvarez MD, PhD^13^, María Queralt Gorgas PhD^14^, Marta Miarons PhD^14^, Pere SoleŕPalacín, MD, PhD^15^, Andrea Martin, MD^15^, Anna Suy MD, Maria Jose Buzon PhD^17^ and Meritxell Genesca PhD^18^.

1. Immunology Department, Hospital Universitari Vall Hebron, Barcelona, Spain

2. Translational Immunology Research Group, Vall Hebron Research Institute (VHIR), Campus Vall Hebron, Barcelona, Spain

3. Department of Cell Biology, Physiology, and Immunology, Universitat Autònoma Barcelona, Campus Vall Hebron, Barcelona, Spain.

4. Microbiology Department, Hospital Universitari Vall Hebron, Barcelona, Spain.

5. Clinical Laboratory Department, Hospital Universitari Vall Hebron, Barcelona, Spain

6. Epidemiology and Public Health Department, Hospital Universitari Vall Hebron, Campus Vall Hebron, Barcelona, Spain.

7. Infectious Disease Department, Hospital Universitari Vall Hebron, Barcelona, Spain.

8. Intensive Medicine Department, Hospital Universitari Vall Hebron, Barcelona, Spain.

9. Organ Dysfunction and Resuscitation Research Group, Vall Hebron Research Institute (VHIR), Campus Vall Hebron, Barcelona, Spain

10. Internal Medicine Department, Hospital Universitari Vall Hebron, Barcelona, Spain

11. Department of Medicine, Universitat Autònoma Barcelona, Campus Vall Hebron, Barcelona, Spain

12. Nephrology Division, Hospital Universitari Vall Hebron, Barcelona, Spain.

13. Pneumology Division, Hospital Universitari Vall Hebron, Barcelona, Spain.

14. Clinical Pharmacy Division, Hospital Universitari Vall Hebron, Barcelona, Spain.

15. Department of Pediatrics, Hospital Universitari Vall Hebron, Barcelona, Spain.

16. Department pf Obstetrics and Gynecology, Hospital Universitari Vall Hebron, Barcelona, Spain.

17. International Health Programme Institut Català de la Salut, Vall Hebron Research Institute (VHIR), Campus Vall Hebron, Barcelona, Spain.

The Immunology Group at Vall d’Hebron Hospital are part of the Universitat Autònoma de Barcelona – Focis Center of Excellence (www.focisnet.org)

The authors thank all the patients and health staff of the Hospitals Vall d’Hebron, Bellvitge, and Germans Trias i Pujol and of the associated hospitals mentioned in the text who endured and did their best to overcome the first wave of COVID-19 in Barcelona, an experience that none of us would ever forget. The authors are grateful to Dr Isabel Novoa Garcia and Ms Sheyla Pascual Martin for their invaluable help in organizing and maintain the COVID-19 sera collection in the Bio-Bank; to Mr Àlex Pérez Rodríguez, Ms Jessica Muñoz, Ms Cinta Rabaza Martí, and Ms Aina Aguiló Cucurull, the technical staff of the immunology laboratory who collected and organized the COVID-19 patient samples during the most difficult days of COVID-19 first wave; and to Ms Adelaida Parada Ramos, the secretary of the Immunology Division, who helped to retrieve the electronic medical records and discharge notes in regard to this study for review by the medical personnel.

## Supplementary supporting information

Sequence of statistical Biomarkers Analyses, text

Table 1S, Patients excluded from HUVH cohort.

Table 2S. Monoclonal Antibodies used in in the flowcytometric phenotypic analysis

Table 3S. Distribution of comorbidities among the severity categories, demographics and hospitalization data from the HUVH cohort.

Table 4S. Demographics and hospitalization data by severity categories from the HUVH cohort.

Table 5S. Pairwise comparison of variables for maximal severity in four severity categories.

Table 6S. Median values of laboratory variables, the proportion out of the normal range and relation with mortality.

Table 7S. ROC curve analysis for all variables, age stratified.

Table 8S. Random Forest model applied to HUVH cohort.

Table 9S. Random Forest Model; comparison exploratory and combination of the three cohorts.

Table 10S. Patients included in immunological studies, cytokines and mononuclear cell phenotype by flowcytometry

## Tables in xlsx format

Correlation of variables r values.xlsx

Correlation of variables p values.xlsx

Repeated Multiple Logistic regression binary severity.xlsx

Repeated Multiple logistic regression decease.xlsx

Multiple logistic regression for follow up.xlsx

## Figures

Fig 1S. Sequence of statistical analyses and summary of conclusions from every step.

Fig 2S. Heatmap summarizing pairwise comparison; p values scale refers to non-parametric tests (Mann-Whitney and Kruskal-Wallis).

Fig 3S. Principal component analysis of the main variables, exitus is equivalent to decease

Fig 4S. Correlograms of the main variables for patients under (A) and over (B) 65 years of age.

Fig 5S. Representative Kaplan-Meyer survival curves clinicodemographic biomarkers (ODRB) and inflammation related biomarkers (IFRB).

Fig 6S. Heatmap summarizing pairwise comparison of the survivors vs deceased in the three hospital cohorts; p values scale refer non-parametric comparisons (Mann-Whitney and Kruskal-Wallis.

Fig 7S. PCA analysis representation of the three hospitals cohorts, indicating similar main vectors, with local differences.

Fig 8S. Heatmap summarizing the reduction of mean Gini index in the Random Forest model that reflects the importance of each variable in the predictive model.

Fig 10S. Performance of cytokines as clinical laboratory test in ROC curve analysis

Fig 11S. Summary of flowcytometry analyses of peripheral blood mononuclear cells.

## References

1 Abers MS, Delmonte OM, Ricotta EE, et al. An immune-based biomarker signature is associated with mortality in COVID-19 patients. JCI Insight 2021;6:m1985. doi:10.1172/jci.insight.144455

2 Zhou F, Yu T, Du R, et al. Clinical course and risk factors for mortality of adult inpatients with COVID-19 in Wuhan, China : a retrospective cohort study. The Lancet 2020;6736:1–9. doi:10.1016/S0140-6736(20)30566-3

3 Chams N, Chams S, Badran R, et al. COVID-19: A Multidisciplinary Review. Frontiers in Public Health 2020;8:1–20. doi:10.3389/fpubh.2020.00383

4 Ghayda RA, Lee J, Lee JY, et al. Correlations of clinical and laboratory characteristics of covid-19: A systematic review and meta-analysis. International Journal of Environmental Research and Public Health 2020;17:1–15. doi:10.3390/ijerph17145026

5 Coma Redon E, Mora N, Prats-Uribe A, et al. Excess cases of influenza and the coronavirus epidemic in Catalonia: a time-series analysis of primary-care electronic medical records covering over 6 million people. BMJ Open 2020;10:e039369. doi:10.1136/bmjopen-2020-039369

6 Guallar MP, Meiriño R, Donat-Vargas C, et al. Inoculum at the time of SARS-CoV-2 exposure and risk of disease severity. International journal of infectious diseases : IJID : official publication of the International Society for Infectious Diseases 2020;97:290–2. doi:10.1016/j.ijid.2020.06.035

7 Marks M, Millat-Martinez P, Ouchi D, et al. Transmission of COVID-19 in 282 clusters in Catalonia, Spain: a cohort study. The Lancet Infectious Diseases 2021;21:629–36. doi:10.1016/S1473-3099(20)30985-3

8 Korber B, Fischer WM, Gnanakaran S, et al. Tracking Changes in SARS-CoV-2 Spike: Evidence that D614G Increases Infectivity of the COVID-19 Virus. Cell 2020;182:812–827.e19. doi:10.1016/j.cell.2020.06.043

9 Andrés C, Piñana M, Borràs-Bermejo B, et al. A year living with SARS-CoV-2: an epidemiological overview of viral lineage circulation by whole-genome sequencing in Barcelona city (Catalonia, Spain). Emerging Microbes and Infections 2022;11:172–81. doi:10.1080/22221751.2021.2011617

10 Meyerholz DK, Perlman S. Does common cold coronavirus infection protect against severe SARS-CoV-2 disease? Journal of Clinical Investigation 2021;131. doi:10.1172/JCI144807

11 Zhang Q, Bastard P, Liu Z, et al. Inborn errors of type I IFN immunity in patients with life-threatening COVID-19. Science (New York, NY) 2020;370:eabd4570. doi:10.1126/science.abd4570

12 Ellinghaus D, Degenhardt F, Bujanda L, et al. Genomewide Association Study of Severe Covid-19 with Respiratory Failure. New England Journal of Medicine 2020;383:1522–34. doi:10.1101/2020.10.06.20205864

13 COVID-19 Host Genetics Initiative. Mapping the human genetic architecture of COVID-19. Nature 2021;600:472–7. doi:10.1038/s41586-021-03767-x

14 Ambale-Venkatesh B, Quinaglia T, Shabani M, et al. Prediction of Mortality in hospitalized COVID-19 patients in a statewide health network. medRxiv : the preprint server for health sciences Published Online First: 19 February 2021. doi:10.1101/2021.02.17.21251758

15 Galván-Román JM, Rodríguez-García SC, Roy-Vallejo E, et al. IL-6 serum levels predict severity and response to tocilizumab in COVID-19: An observational study. Journal of Allergy and Clinical Immunology 2021;147:72–80.e8. doi:10.1016/j.jaci.2020.09.018

16 Gupta RK, Marks M, Samuels THA, et al. Systematic evaluation and external validation of 22 prognostic models among hospitalised adults with COVID-19: an observational cohort study. The European respiratory journal 2020;56:2003498. doi:10.1183/13993003.03498-2020

17 Jehi L, Ji X, Milinovich A, et al. Development and validation of a model for individualized prediction of hospitalization risk in 4,536 patients with COVID-19. PLOS ONE 2020;15:e0237419. doi:10.1371/journal.pone.0237419

18 Knight SR, Ho A, Pius R, et al. Risk stratification of patients admitted to hospital with covid-19 using the ISARIC WHO Clinical Characterisation Protocol: development and validation of the 4C Mortality Score. BMJ 2020;2:m3339. doi:10.1136/bmj.m3339

19 Marcolino MS, Pires MC, Ramos LEF, et al. ABC2-SPH risk score for in-hospital mortality in COVID-19 patients: development, external validation and comparison with other available scores. International Journal of Infectious Diseases 2021;110:281–308. doi:10.1016/j.ijid.2021.07.049

20 Riveiro-Barciela M, Labrador-Horrillo M, Camps-Relats L, et al. Simple predictive models identify patients with COVID-19 pneumonia and poor prognosis. PLoS ONE 2020;15. doi:10.1371/journal.pone.0244627

21 Smith GB, Prytherch DR, Meredith P, et al. The ability of the National Early Warning Score (NEWS) to discriminate patients at risk of early cardiac arrest, unanticipated intensive care unit admission, and death. Resuscitation 2013;84:465–70. doi:10.1016/j.resuscitation.2012.12.016

22 Wynants L, Van Calster B, Collins GS, et al. Prediction models for diagnosis and prognosis of covid-19: systematic review and critical appraisal. BMJ 2020;369:m1328. doi:10.1136/bmj.m1328

23 Mathew D, Giles JR, Baxter AE, et al. Deep immune profiling of COVID-19 patients reveals distinct immunotypes with therapeutic implications. Science 2020;369. doi:10.1126/science.abc8511

24 Lucas C, Wong P, Klein J, et al. Longitudinal analyses reveal immunological misfiring in severe COVID-19. Nature 2020;584:463–9. doi:10.1038/s41586-020-2588-y

25 Wilk AJ, Rustagi A, Zhao NQ, et al. A single-cell atlas of the peripheral immune response in patients with severe COVID-19. Nature Medicine 2020;26:1070–6. doi:10.1038/s41591-020-0944-y

26 Kuri-Cervantes L, Pampena MB, Meng W, et al. Comprehensive mapping of immune perturbations associated with severe COVID-19. Science Immunology 2020;5. doi:10.1126/sciimmunol.abd7114

27 Mann ER, Menon M, Knight SB, et al. Longitudinal immune profiling reveals key myeloid signatures associated with COVID-19. Science Immunology 2020;5:eabd6197. doi:10.1126/sciimmunol.abd6197

28 Arunachalam PS, Wimmers F, Mok CKP, et al. Systems biological assessment of immunity to mild versus severe COVID-19 infection in humans. Science 2020;369:1210–20. doi:10.1126/SCIENCE.ABC6261

29 Mueller YM, Schrama TJ, Ruijten R, et al. Stratification of hospitalized COVID-19 patients into clinical severity progression groups by immuno-phenotyping and machine learning. Nature Communications 2022;13:915. doi:10.1038/s41467-022-28621-0

30 Marshall JC, Murthy S, Diaz J, et al. A minimal common outcome measure set for COVID-19 clinical research. The Lancet Infectious Diseases 2020;20:e192–7. doi:10.1016/S1473-3099(20)30483-7

31 Levey AS, Stevens LA, Schmid CH, et al. A new equation to estimate glomerular filtration rate. Annals of internal medicine 2009;150:604–12. doi:10.7326/0003-4819-150-9-200905050-00006

32 Maecker HT, McCoy JP, Nussenblatt R. Standardizing immunophenotyping for the Human Immunology Project. Nature Reviews Immunology. 2012;12:191–200. doi:10.1038/nri3158

33 Garcia-Prat M, Álvarez-Sierra D, Aguiló-Cucurull A, et al. Extended immunophenotyping reference values in a healthy pediatric population. *Cytometry Part B*, Clinical cytometry 2019;96:223–33. doi:10.1002/cyto.b.21728

34 Barcelona demography, June 2020. https://www.ine.es/jaxiT3/Datos.htm?t=2861

35 Sempere A, Salvador F, Monforte A, et al. Covid-19 clinical profile in latin american migrants living in spain: Does the geographical origin matter? Journal of Clinical Medicine 2021;10:1–9. doi:10.3390/jcm10225213

36 Ferrer R, Báguena M, Balcells J, et al. Planning for the assistance of critically ill patients in a Pandemic Situation: The experience of Vall d’Hebron University Hospital. Enfermedades infecciosas y microbiologia clinica (English ed) 2020;110:697–700. doi:10.1016/j.eimc.2020.08.007

37 Chen Z, John Wherry E. T cell responses in patients with COVID-19. Nature reviews Immunology 2020;20:529–36. doi:10.1038/s41577-020-0402-6

38 Cantenys-Molina S, Fernández-Cruz E, Francos P, et al. Lymphocyte subsets early predict mortality in a large series of hospitalized COVID-19 patients in Spain. Clinical and experimental immunology 2021;203:424–32. doi:10.1111/cei.13547

39 Röltgen K, Powell AE, Wirz OF, et al. Defining the features and duration of antibody responses to SARS-CoV-2 infection associated with disease severity and outcome. Science Immunology 2020;5:eabe0240. doi:10.1126/sciimmunol.abe0240

40 Collins GS, Reitsma JB, Altman DG, et al. Transparent reporting of a multivariable prediction model for individual prognosis or diagnosis (TRIPOD): The TRIPOD statement. Annals of Internal Medicine 2015;162:55–63. doi:10.7326/M14-0697

41 Hadjadj J, Yatim N, Barnabei L, et al. Impaired type I interferon activity and inflammatory responses in severe COVID-19 patients. Science 2020;369:718–24. doi:10.1126/science.abc6027

42 Lucas C, Klein J, Sundaram ME, et al. Delayed production of neutralizing antibodies correlates with fatal COVID-19. Nature medicine 2021;27:1178–86. doi:10.1038/s41591-021-01355-0

43 Sette A, Crotty S. Adaptive immunity to SARS-CoV-2 and COVID-19. Cell 2021;184:861– 80. doi:10.1016/j.cell.2021.01.007

