## Supplementary material for "Exposing Limitations of Clinical Laboratory Tests in COVID-19 and the Promise of Immunological Biomarkers": SEQUENCE OF STATISTICAL BIOMARKERS ANALYSES

#### SEQUENCE OF STATISTICAL BIOMARKERS ANALYSES

The statistical analysis sequence and main conclusions are summarised in figure 1S. The sequence is split into exploratory, in depth and complementary analyses.

**Exploratory Analysis**
22 of 29 available variables were found strongly associated to 28-day outcome by pairwise comparison (table 2, and figure 3, main text). Despite these strong associations, Multiple Logistic Regression, Random Forest Model and Principal Component (PCA) analysis showed limited prediction power as only around half of decease/severe cases were correctly classified in the multiple logistic regression and random forest classification tables and separation was poor in PCA (tables 3 and 8S and figure 3S). To explain this limitation, multiple mutual correlation analysis was carried out (figure 4) and conclusions were confirmed by repeated logistic regression, see sections below "Complexity reduction and the weight of age" and "Repeated multiple Logistic regression…".

**In Depth Analyses**
*Multiple correlations*
In the global correlogram (figure 4), lymphocytes, monocytes and eosinophils constitute a cluster of variables that correlated positively with each other (r=0.42 to r=0.3), while neutrophils, basophils and platelets form another cluster (r= 0.4 to r=0.38); each cluster kept a negative correlation with the other (lymphocytes with neutrophils % r= -0.95). These reciprocal changes in neutrophil and lymphocyte clusters are also seen across the severity categories; Acute Phase Reactants (APRs) correlated among themselves (r=0.71 and r=0.46 for IL-6 with CRP and with Ferritin respectively) with coagulation factors (r=0.67, r=0.42, r=0.35 for IL-6, with fibrinogen, D-dimer and prothrombin time (INR) respectively) and with neutrophils %, (IL-6 r=0.6, CRP r=0.6). This may reflect the central position of CRP in the network of interactions typically occurring in infection diseases and systemic inflammation in which IL-1, IL-6 and TNF-alpha all act synergistically on the liver increasing the production of acute phase proteins and coagulation factors.[1] Age correlated strongly with kidney function tests (r=0.6, r=0.32 and r=-0.6 for urea, creatinine and GFR respectively), moderately with AST/ALT ratio (r =0.40) and weakly with SpO2/FiO2 (r=0.32). All above correlations were significant, (tables in excel format r and p values of correlogram). The relative weight of age in different age intervals was investigated by comparing the correlograms of patients over and under 65 years which showed that they are maintained (figure 4S <65 years vs >65 years correlation heatmaps). These networks of correlations explain the compound effect of age on the clinical laboratory variables and indicated that the limited prediction of power of biomarker combinations is due to redundancy. This conclusion was supported by the VIF indexes of many variables in the multiple logistic regression analysis, see below the sections "Repeated multiple logistical regression to confirm redundancy and identification minimal set of variables".

*Complexity reduction and the weight of age*
The above analysis and the understanding of the biological interrelations of the variables led to reduce the complexity of the analysis by combining variables of physiopathologically related

families that were statistically correlated in: 1) Blood including Haemoglobin (Hb), Blood White Cell Count (WBC) and differential counts in % and number, Neutrophil to Lymphocyte Ratio (NLR) and platelets; 2) Acute Phase Reactants including C Reactive Protein (CRP), IL-6 and Ferritin; 3) Coagulation including D-dimer, fibrinogen, prothrombin time (INR); 4) Liver tests including Bilirubin direct and total, AST, ALT and AST/ALT ratio); and 5) Kidney function tests including urea, creatinine and glomerular filtration rate (GFR). As these groups of variables were found to behave similarly as predictors of survival and of severity and they are known to participate in common pathophysiological networks, we further combined them in Clinical Demographic (CD) variables, InFlammation Related Biomarkers (IFRB) that include the blood, APRs and coagulation and in Organ Damage Related biomarkers (ODRB) that include liver and kidney tests plus SpO2/FiO2, as a biomarker of lung damage.

In the pairwise comparisons for survival/decease the lowest p values among CDs were for age and comorbidities e.g., exact $p=7.26x10^{-81}$ and $2.3x10^{-38}$ respectively (table 2). Among ODRBs the lowest p values were GFR $2.37x10^{-101}$, AST/ALT ratio $6.06x10^{-31}$, while among IFRBs lowest p values were IL-6, $10^{-55}$; CRP 5.99x10-$^{43}$; and NLR $0.34x10^{-41.}$

The analyses of variables in patients split into four maximal severity categories (Kruskal-Wallis test) showed that in severe vs deceased, ODRB (GFR and AST/ALT ratio) kept a high differential association (p values $10^{-11}$-$10^{-112}$ ) while the association with IFRBs, APR and WBC differential counts was weaker i.e., only Hb, platelets and coagulation factors were significantly associated to outcome. Age and comorbidities are differentially associated to severity categories as outcome with p values of $2.8x10^{-53}$ and $10^{-10}$ respectively but in the comparison of moderate vs severe, age association was not significant and the significancy of the associations is low for ODRB while is high for IFRBs (table 4S and figure 2S).

The bivariate logistic regression analysis of the 19 main biomarkers adjusted by age confirmed that the association of 16 of 19 variables including ODRB and IFRB, with survival and severity respectively is only partially linked to age. The corresponding age corrected Z values followed the ranking IL-6> CRP> SpO2/FiO2> neutrophiles %> NLR > Monocytes % > Neutrophils n > GFR > lymphocytes % for survival/decease and IL-6> CRP> neutrophiles % SpO2/FiO2>> NLR > lymphocytes % > Monocytes % for severity where GFR is displaced to position 11th in the ranking (table 4).

Kaplan-Mayer survival curve analysis using Youden indexes from laboratory test performance ROC curves as cut-off (see next section Performance of variables by ROC curves…) was applied to assess the relation of each biomarker with survival within the 28d period (figures 5B and 5S). Age had the highest hazard ratio (32.8), followed by GFR (9.3), urea (6.3), IL-6 (5.9), D-dimer (4.7), comorbidities (4.7), AST/ALT (4.3), CRP (4.3), SpO2/FiO2 (2.8) and differential WBC % (2.8–2.6) while platelets, ferritin and sex gave low or no-significant hazard ratios.

*Performance of variables by ROC curve analysis as applied to clinical laboratory tests*
ROC curves were generated to assess biomarker predictive power in the clinical context. For survival/decease as outcome, GFR, IL-6, AST/ALT, and SpO2/FiO2 showed the best curves, AUCs (CI): 0,80 (0.77-0.83), 0.77 (0.73-0.81), 0.73 (0.69–0.77) and 0.73 (0.70-0.78) respectively, followed by the other APRs and blood variables. Age, treated as a variable for

comparison, gave an AUC of 0.87 (0.85-0.89) better than any of other variables; comorbidities gave an AUC 0.75 (0.72-0.78) (table 5 and figure 5A). For non-severe vs severe as outcome, the larger AUC corresponded to: IL-6, 0.78 (0.75-0.80) followed by SpO2/FiO2, 0.77 (0.74-0.81), CRP, 0.75 (0.71-0.77), NLR 0.71 (0.68-0.73), GFR 0.69 (0.65-0.71) and age 0.67 (0.64-0.70). The comparison of the ROC curves to predict severity with those to predict survival shows that ODRBs are better predictor of survival/decease and IFRBs of severity/non-severe except for SpO2/FiO2 that is a very good predictor of both outcomes (table 5). To reduce the effect of age better assess the effect of the other variables, patients were stratified by age intervals (40-55, 55-65, 66-75, 76-85 and >85 years old); the variable giving the larger AUC across all age group was SpO2/FiO2 (0.72 to 0.79) except in over 85 year-old patients in which IL-6 had the larger AUC (0.79); in this stratified analysis tests, IFRBs variables were found to be better than ODRBs probably because there is higher collinearity of ODRB with age, as suggested by the reduction of their prediction power when stratified by age (table 7S).

*Repeated multiple logistical regression to identify of the minimal set of variables, and redundancy confirmation*

Repeated multiple logistic regression analyses of variables for outcomes decease and severity were carried out to confirm their redundancy, calculating VIF scores of collinearity to identify the combinations giving with the best prediction score. The exploratory analysis had already indicated some redundancies, and the number of variables was reduced to 19 which included age, sex, comorbidity index, Hb, neutrophil, lymphocytes, monocyte, and eosinophils % and number (n), NLR, platelets, CRP, IL-6, D-dimer, ferritin, fibrinogen, prothrombin time INR, SpO2/FiO2, AST/ALT ratio and GFR. Notice that when SpO2/FiO2 was included, the number of observations was reduced to 411, as SpO2/FiO2 was available in only 52% of these patients. The scores generated for each variable were odd ratios (OR), Z scores, p values, VIF, area under the ROC curve (AUC, CI) at 50% cut-off, Positive Predictive Value (PPV, predicting either decease or severity) and Negative Predictive Value (NPV, predicting survival or non-severity) and the % of correctly classified patients for the outcome decease or severity (tables logistic regression in xlsx format).

For survival/decease prediction with 19 variables, scores were AUC 0.95, PPP 75.7, NPP 94.4 and 54.4% of deceased patients correctly classified (n=411); if age, comorbidities, and sex were excluded, values were AUC 0.84, PPP 76.6, NPP 80.6 and 58.2% correctly classified (n=411). When SpO2/FiO2, was excluded from the biomarkers the prediction scores were AUC 0.86, PPP 68.9, NPP 92.9 and correct classification of decease cases was reduced to 22.7 %, however these variables correctly classified 99% of the patients who survived as deduced by the PPP and NPP (n=994). Age, comorbidity, and sex by themselves would give an AUC of 0.89, PPP 56.7, NPP 88.5 and 36.5% deceased correctly classified (n=1,579) with sex not modifying the scores. Therefore, while, if additive, the prediction of demographics and biomarkers should give 90.7% of correctly classified decease cases, only 58.2% of cases were actually correctly classified. A reduced set of variables to predict survival/decease was generated by repeated analyses progressively excluding redundant variables thus reducing the VIF scores of collinearities; the reduced set including eight variables: age, comorbidity index, SpO2/FiO2, NLR, CRP, fibrinogen, AST/ALT ratio and GFR gave AUC 0.94, PPP 83.3, NPP 94.1, and 65.8 of deceased patients were correctly classified (n=502).

The same analysis with 19 variables applied to non-severity/severity outcome gave AUC 0.85, PPP 74.7%, NPP 79.2 and 54.6 of severe were correctly classified (n=411). If age and comorbidities were excluded, scores were AUC 0.84, PPP 76.6% NPP 80.6, and 58.2 % of severe cases correctly classified. When SpO2/FiO2, was excluded, the prediction scores were AUC 0.81, PPP 65.3, NPP 81.0 and correct classification of severe cases was reduced to 36.2% (n=994). The reduced set of eight variables gave AUC of 0.86, PPP 81,5, NPP 80.3 and correctly classified 64,1% of the severe cases (n=502).

**Time course and biomarkers**

The predictive value of laboratory variables was expected to evolve during the disease course but because of the retrospective nature of this study, data at regular intervals for every patient were not available. However, as we had data from 7,586 additional follow up samples corresponding 1,079 of the 1,579 patients, we plotted them to generate an approximation of the evolution of the variables along the 28-days follow-up period. The curves representing means ± CI values for each variable for survivors and deceased maintained a clear separation during the initial 10 days and only overlapped at the end of the 28d period. Of note, NLR curves were more clearly separated between days 5 and 20, while IL-6 values overlapped after day 4-5; CRP, lymphocyte, and neutrophil % curves remained separated over most of the period; the eosinophil curve shows a remarkable increase in survivors but only after day 10. GFR curves were clearly separated from the beginning, but the mean values differ little from the normal range. From this analysis it cannot be deduced whether IFRBs precede ODRB or viceversa. (figure 6)

**Additional Analyses: Comparison of the exploratory with validation cohorts**

The patient populations in the three hospitals were not matched for demographics nor mortality (table 6). The mortality rate was higher in HUB (25.7% vs. 16.1% and 12.3% in HUVH and HUGTP, respectively). This may be explained by their higher age, 65 (5374) as compared to HUVH 62 (50-75) and in HUGTP 62 (52-71) (table 6). There are also differences in the median laboratory variables that indicates that this cohort includes more critically ill patients. These differences however made these almost contemporary cohorts adequate for comparing the relative predictive power of clinical laboratory test with demographics and clinical variables in a real-life situation.

The pairwise univariate analysis of laboratory variables association with 28d outcome of the HUB and HUGTP data sets were conducted similarly as for the HUVH data set. The significant association was confirmed for seven blood variables, three APRs (CRP, IL-6, and ferritin), and the D-dimer (table 6 and figure 6S). The only index of renal function that was available in the three hospitals, urea, was clearly validated, and this supports the findings from the HUVH data of creatinine and GFR as having strong prognostic value after adjustment for age.
Multiple logistic regression analysis that was adjusted for age showed that the ranking of the odds ratios of clinical laboratory variables associated with mortality was similar in the three centres. The APRs, IL-6, CRP, ferritin, and D-dimer occupy the top positions of the ranking, followed by neutrophils and, in the other side, lymphocytes, monocytes, and eosinophils (figure 7).

The PCA of 17 clinical laboratory variables show an almost complete overlap that support identical basic physiopathology of the disease, regardless of the differences in the patient population (figure 7S).

The Random Forest model was applied to the common variables of the three cohorts and the reduction of the mean Gini index in the exploratory HUVH cohort and in the combination of the three cohorts were calculated. Table 9S and figure 8S show the similar rankings of the variables in the different analyses.

The clinical laboratory test performance as assessed by the AUC from ROC curves from the three hospitals, when subjected to unbiased clustering, showed the central role of APRs and kidney function; the AST/ALT ratio and GFR, available only from the HUVH cohort, showed the larger AUC, together but AUC of cytokines from the immunological studies, see below, section expanded immunological biomarkers (figure. 8 and table 7).

**Additional Analyses: Pilot study of Immunological tests**

*Cytokine profile*
Cytokines were measured in 74 patients, who were representative of the HUVH cohort's moderate and severe disease categories (age 53 [44–64] years) (see table 10S Immunological studies patient). The ELLA platform to measure cytokines has been in use in the HUVH immunology laboratory for six years to monitor cytokines in transplantation and sepsis projects and has proved very robust. Samples were collected on days 0 and +2. The levels of cytokines IL-6, TNF-α, IL-10, IL-2, CXCL10, and CCL2 as well as the receptor antagonist of IL-1 (IL-1RA) reached significantly higher values in the severe patients and tended to increase over time. The remaining cytokines and sCD163 did not show significant differences in relation to time and severity categories. On day 0, the IFN-α decreased, but increased in two mild cases; this may be interpreted as indicating the end of an initial peak in most patients. The IL 12p70 level seemed to follow a similar pattern, but the data did not show significant differences (figure 9A & B).
The comparison of correlograms of cytokines at days 0 and 2 did not show significant changes in the mutual correlations, probably because the interval was too short. However, greater differences were seen between the moderate and severe of the four categories of patients, with a loss of the correlation with IFN-α probably attributable to its quick drop in severely ill patients. We also observe a moderate increase in the correlations around IL-17 (figure 9S).
Predictive power of cytokines and sCD163 were compared for both survival and severity as outcome. For 7 of the 20 cytokines, the survival predictive power as measured by the AUC of the ROC curves at day 0 was significant and followed the order CXCL10 > ILRA > IL-6 > CCL2 > IL10 > IL-15 > IL-7 > TNF-α > IFN-α. On day +2, the predictive power order for IL-6, CCL2, IL-15, TNF-α, and IL-7 and significant for eight of them. The prediction power for non-severity vs severity categories followed the order CXCL10 > IL-6 > IL1-RA > IL-15 at day 0 and IL-6, CCL2, IL-2 at day 2 of the 10 largest AUCs were statistically significant. The ROC curves for the main clinical laboratory biomarkers were also calculated for this group and in the comparison showed smaller AUCs than these cytokines (table 5 and figure 10S).

*Blood extended phenotyping*

Of the 41 patients, 5, 27, 6, and 3 were in the mild, moderate, severe, and deceased severity categories, respectively. The median age was lower than that of the HUVU cohort, although the M/F proportion, DFSO, and LOS were similar (table 10S immunology cohort). As adapted for this project, the HIPC protocol generated 163 variables corresponding to 42 lymphocytes, monocytes, and neutrophil subsets.
In pairwise comparison for decease and severity outcomes, 12 lymphocyte subsets were significantly associated with the 28-day decease according to a ranking as follows: naïve T lymphocytes (n) > Th2 (n) > total T lymphocytes > CD8 T lymphocytes (n) > CD8 TCM (n) > CD8 TEM (n)> CD4 T lymphocytes (n) > CD4EM (n) > non-classical monocytes. Remarkably, for this subgroup, the significance was similar to that of CRP, IL-6, and D-dimer. Interestingly, the CD4 TEM and CD8 TEM populations showed a continuous decrease with the disease severity, whereas the naïve populations showed a surge in the most severe cases (probably, due to the mobilization of naïve cells which did not contribute to improved outcome (figure 10).
In the correlogram between laboratory biomarkers and cell phonotypes there is a negative correlation of APRs and T-cell populations except for Th1- and Th17-activated T cells. There is, therefore, an inverse correlation between inflammation and the T-cell subsets that drive the adaptive immune response; there is a strong negative correlation of age with CD8-naïve T cells, which is critical for the immune response to the virus; these changes reveal the deep dysregulation of lymphocyte biology in COVD-19 patients (figure 11S), The positive correlations between APR and B cells could be interpreted as resulting from an accelerated recruitment of naïve B cells to the switching stage; furthermore, there is a steeper reduction in non-classical monocytes and a reduction in their DR expression. This fits well with the high mortality rate seen among patients with <2% monocytes and with the lack of changes in the sCD163 (see the section titled Cytokines), which is a marker of M2 macrophages. The ROC curves of CD3+CD62L (n) naïve T cells had an AUC of 0.74, with a sensitivity and specificity of 66.7% and 72%, respectively, for severity (table 5).

References:

1 Jones BE, Maerz MD, Buckner JH. IL-6: a cytokine at the crossroads of autoimmunity. Curr. Opin. Immunol. 2018; **55**: 9–14.

**Table 1S. Patients excluded from HUVH cohort.**

| Groups | Number of patients | Sex (male/female ratio) | Age median (IQR) | 28-day mortality (%) |
|---|---|---|---|---|
| Nosocomial | 57 | 57.9% | 67 (60–75) | 36.8% |
| Severe oncological disease | 35 | 65.7% | 66 (58–74.25) | 34.3% |
| Pregnancy | 15 | NA | 76 (76–76) | 0.0% |
| Transplanted | 13 | 84.6% | 53 (51–55) | 46.2% |
| Severe haematological disease | 11 | 36.4% | 37 (30.75–44.25) | 36.4% |
| Paediatric | 11 | 36.4% | 75 (74.5–75.5) | 0.0% |
| Autoimmune disease | 7 | 28.6% | 65 (56.5–71) | 14.3% |
| Referred for ECMO | 7 | 71.4% | 61 (59.5–69) | 0.0% |
| Primary immunodeficiency | 1 | 0.0% | 44 (NA) | 0.0% |
| Other severe disease | 1 | 0.0% | 49 (NA) | 0.0% |
| Total | 158 | | | 27.8% |

Patients who were excluded from the HUVH cohort and their pathology.ECMO, ExtraCorporeal Membrane Oxygenation; NA, Not Applicable.

**Table 2S. Monoclonal Antibodies used in in the flowcytometric phenotypic analysis**

| | ISOTYPE | CLONE |
|---|---|---|
| General lymphocyte populations | | |
| CD45-FITC/CD8-PE/CD4-ECD/CD3-PC5 | IgG2b / IgG1 / IgG1 / IgG1 mouse | B3821F4A/SFCI12T4D11/SFCI21Thy2D3/UCHT1 |
| CD45-FITC/CD56-PE/CD19-ECD/CD3-PC5 | IgG2b / IgG1 / IgG1 / IgG1 mouse | B3821F4A/SFCI12T4D11/SFCI21Thy2D3/UCHT1 |
| T- Lymphocyte populations | | |
| CXCR3/CD183 AF488 | IgG1 mouse | G025H7 |
| CCR7/CD197 PE | IgG2a mouse | G043H7 |
| CD45RA ECD | IgG1 mouse | ALB11 |
| CCR6/CD196 PC7 | IgG2a mouse | B-R35 |
| CD4 APC | IgG1 mouse | 13B8.2 |
| CD8 APC700 | IgG1 mouse | SFCI21Thy2D3 (T8) |
| CD3 APC750 | IgG1 mouse | UCHT1 |
| HLADR PB | IgG1 mouse | Immu-357 |
| CD45 KRO | IgG1 mouse | J.33 |
| Recent thymic emigrants | | |
| CD31 FITC | IgG1 mouse | 5.6E |
| CD62L PE | IgG1 mouse | DREG56 |
| CD3 ECD | IgG1 mouse | UCHT1 |
| CD27 PC7 | IgG1 mouse | 1A4CD27 |
| CD4 APC | IgG1 mouse | 13B8.2 |
| CD45RA PB | IgG1 mouse | 2H4LDH11LDB9 (2H4) |
| CD45 KRO | IgG1 mouse | J.33 |
| T-regulatory cells | | |
| CD45RO FITC | IgG2a mouse | UCHL1 |
| CD25 PE | IgG2a mouse | B1.49.9 |
| CD3 ECD | IgG1 mouse | UCHT1 |
| CCR4/ CD194 PC7 | IgG1 mouse | 1G1 |
| CD4 APC | IgG1 mouse | 13B8.2 |
| CD127 APC750 | IgG1 mouse | R34.34 |
| HLADR PB | IgG1 mouse | Immu-357 |
| CD45 KRO | IgG1 mouse | J.33 |
| DC/Monocytes/NK | | |
| CD16 FITC | IgG1 mouse | 3G8 |
| CD11c PE | IgG1 mouse | BU15 |
| CD3 ECD | IgG1 mouse | UCHT1 |
| CD19 ECD | IgG1 mouse | J3-119 |
| CD20 ECD | IgG2a mouse | B9E9(HRC20) |

| | | |
|---|---|---|
| CD56 PC7 | IgG1 mouse | N901 (NKH-1) |
| CD123 APC | IgG1 mouse | SSDCLY107D2 |
| CD14 APC750 | IgG1 mouse | RMO52 |
| HLADR PB | IgG1 mouse | Immu-357 |
| B Lymphocytes | | |
| IgD FITC | IgG2a mouse | IA6-2 |
| CD21 PE | IgG1 mouse | BL13 |
| CD19 ECD | IgG1 mouse | J3.119 |
| CD27 PC7 | IgG1 mouse | 1A4CD27 |
| CD24 APC | IgG1 mouse | ALB9 |
| CD38 APC750 | IgG1 mouse | LS198-4-3 |
| IgM PB | IgG1 mouse | SA-DA4 |
| CD45 RO | IgG1 mouse | J.33 |

DC, dendritic cells; NK, Natural Killer.

**Table 3S. Distribution of comorbidities among the severity categories and demographics and hospitalization data in the HUVH cohort**

| Comorbidities | n (% ) | Age (mean) | Male | Female | M/F % | DFSO (days) [mean ± SD] | LOS (days) [mean ± SD] | Mortality (%) | Mild (n) | Moderate (n) | Severe (n) | Deceased (n) |
|---|---|---|---|---|---|---|---|---|---|---|---|---|
| **0** | 546 (34.6%) | 51.6 | 305 | 241 | 0.56 | 7.8 ± 4.4 | 14.9 ± 22.4 | 2.9% | 40 | 400 | 89 | 16 |
| **1** | 431 (27.3%) | 61.6 | 240 | 191 | 0.56 | 8.1 ± 5.4 | 17.1 ± 24.0 | 12.8% | 15 | 273 | 88 | 55 |
| **2** | 303 (19.2%) | 71.1 | 165 | 138 | 0.54 | 6.7 ± 4.7 | 17.6 ± 23.5 | 28.4% | 9 | 162 | 46 | 86 |
| **3** | 186 (11.8%) | 71.2 | 101 | 85 | 0.54 | 6.3 ± 4.6 | 24.2 ± 30.5 | 28.5% | 5 | 93 | 35 | 53 |
| **4** | 88 (5.6%) | 72.1 | 52 | 36 | 0.59 | 5.2 ± 3.9 | 20.2 ± 26.4 | 39.8% | 2 | 31 | 20 | 35 |
| **5–7** | 25 (1.6%) | 73.7 | 17 | 8 | 0.59 | 5.20 ± 3.8 | 33.9 ± 32.9 | 40.0% | 0 | 6 | 4 | 10 |

DFSO, days from symptom onset; LOS, length of stay; n, number. For comorbidities considered, see text

**Table 4S. Demographics and hospitalization data by severity categories of the HUVH cohort**.

| Severity among patients (n = 1579) | Mild | Moderate | Severe | Deceased | Mild vs. Moderate (Adj. p-values) | Mild vs. Severe (Adj. p-values) | Mild vs. Deceased (Adj. p-values) | Moderate vs. Severe (Adj. p-values) | Moderate vs. Deceased (Adj. p-values) | Severe vs. Deceased (Adj. p-values) | Multiple comparison (Adj. p-values) |
|---|---|---|---|---|---|---|---|---|---|---|---|
| Patients | 71 (4.5%) | 969 (61.4%) | 284 (18.0%) | 255 (16.1%) | | | | | | | |
| Age median (IQR) | 47 (36–69) | 58 (48–72) | 55 (51–68) | 82 (74–87) | 7.55E-04 | 7.50E-03 | 4.94E-36 | 3.80E-01 | 1.65E-72 | 2.28E-53 | <0.001 |
| Female n (%) | 44 (62.0%) | 446 (46.1%) | 101 (35.6%) | 107 (39.6%) | 1.32E-02 | 7.60E-05 | 3.00E-03 | 1.73E-03 | 2.6E-01 | 1.3E-01 | <0.001 |
| DFSO median (IQR) | 8 (5–10) | 7 (5–10) | 7 (4–9) | 5 (2–7) | 5.10E-01 | 4.69E-02 | 7.42E-08 | 6.83E-03 | 9.50E-20 | 1.09E-07 | <0.001 |
| LOS median (IQR) | 2.0 (2–2) | 5 (2–11) | 35 (17–59) | 7 (4–11) | 1.53E-06 | 8.74E-41 | 1.57E-08 | 7.93E-69 | 1.73E-02 | 5.35E-32 | <0.001 |
| Disease duration median (IQR) | 10 (8–16) | 13 (9–21) | 41 (25–66) | 12 (9–18) | 1.01E-02 | 1.92E-24 | 5.96E-01 | 2.44E-53 | 4.95E-04 | 5.30E-50 | <0.001 |
| Composite Morbidity (mean ± SD) | 0 (0–1) | 1 (0–2) | 1 (0–2) | 2 (1 ) | 2.00E-02 | 2.00E-04 | <1.0E-10 | 9.9E-04 | 1.00E-10 | 1.00E-10 | <0.001 |

Statistical comparisons include univariate comparisons, using Kruskal–Wallis with the two-stage step-up method of Benjamini and Hochsberg for the adjustment of the false-discovery rate. IQR, 25–75 interquartile range; DFSO, days from symptom onset; LOS, length of stay.

#### Table 5S. Pairwise comparison of variables for maximal severity in four categories

| | Severity categories | | | | Multiple comparisons | | | | | | |
|---|---|---|---|---|---|---|---|---|---|---|---|
| **Variables** | **Mild (n = 71) Median (IQR)** | **Moderate (n = 969) Median (IQR)** | **Severe (n = 275) Median (IQR)** | **Deceased (n = 255) Median (IQR)** | **Mild vs. Moderate (exact p-value)** | **Mild vs. Severe (exact p-value)** | **Mild vs. Deceased (exact p-value)** | **Moderate vs. Severe (exact. p-value)** | **Moderate vs. Deceased (exact. p-value)** | **Severe vs. Deceased (exact p-value)** | **Global comparison (Adj. *p*-value)** |
| **Age** | 47 (36–69) | 58 (48–72) | 55 (51–68) | 82 (74–87) | **7.55E-04** | **7.50E-03** | **4.943E-36** | 0.380 | **1.65E-72** | **2.28E-53** | <0.001 |
| **Morbidity Index (mean ± SD)** | 0 (0–1) | 1 (0–2) | 1 (0–2) | 2 (1) | **2.00E-02** | **2.00E-04** | **<1.0E-10** | **9.9E-04** | **1.00E-10** | **1.00E-10** | **<0.001** |
| **INFLAMMATION RELATED BIOMARKERS (IRFBS)** | | | | | | | | | | | |
| **Blood** | | | | | | | | | | | |
| Hb (12–15 g/dL) | 13.8 (12.9–14.5) | 13.7 (12.5–14.6) | 13.5 (12.4–14.6) | 12.7 (11.4–13.8) | 2.80E-01 | 0.100 | **7.63E-07** | 0.230 | **4.70E-14** | **3.71E-07** | <0.001 |
| WBC (4–11 $10^9$/L) | 6.3 (5.1–8.0) | 6.4 (4.7–8.8) | 7.2 (5.3–10.1) | 7.6 (5.3–10.7) | 9.77E-01 | 0.026 | **6.20E-03** | **1.38E-05** | **1.33E-07** | 0.435 | <0.001 |
| Neutrophils (40–80%) | 65 (57.0–72.4) | 74 (66.7–80.2) | 81.3 (73.7–86.7) | 83 (75.4–88.0) | **4.99E-09** | **1.17E-24** | **4.68E-27** | **3.29E-21** | **2.51E-25** | 0.389 | <0.001 |
| Neutrophils (2–7 $10^9$/L) | 4.02 (3.11–5.45) | 4.56 (3.35–6.16) | 5.59 (3.94–8.15) | 6.12 (3.99–8.65) | **3.98E-02** | **2.93E-07** | **2.56E-08** | **6.42E-10** | **2.89E-12** | 0.491 | <0.001 |
| Lymphocytes (20–50%) | 24.9 (19.1–34.0) | 17.4 (12.5–24.1) | 12.3 (7.9–18.5) | 10.3 (7.1–16.7) | **2.64E-09** | **2.27E-23** | **3.46E-29** | **4.98E-18** | **5.02E-28** | 0.045 | <0.001 |
| Lymphocytes (1.2–3.5 $10^9$/L) | 1.65 (1.22–2.26) | 1.08 (0.81–1.44) | 0.87 (0.62–1.14) | 0.78 (0.55–1.04) | **1.24E-11** | **2.02E-23** | **9.68E-31** | **7.62E-13** | **1.01E-23** | 0.014 | <0.001 |
| Monocytes (0.1–1.0%) | 8.1 (6.7–9.4) | 7.1 (5.4–9.0) | 5.2 (3.8–7.3) | 5.4 (3.5–7.8) | **5.90E-03** | **5.67E-13** | **1.08E-10** | **4.14E-19** | **8.00E-14** | 0.262 | <0.001 |
| Monocytes (0.1–1 $0^9$/L) | 0.51 (0.38–0.71) | 0.45 (0.32–0.60) | 0.37 (0.26–0.54) | 0.4 (0.28–0.56) | **7.40E-03** | **1.00E-06** | **6.85E-05** | **3.74E-06** | **0.004** | 0.178 | 0.19 |
| Eosinophils (0.0–5.0%) | 0.4 (0.0–1.0) | 0 (0.0–0.2) | 0 (0–0.1) | 0 (0–0) | **1.38E-14** | **2.01E-21** | **4.36E-23** | **5.32E-06** | **1.24E-07** | 0.480 | <0.001 |
| Eosinophils (0.0–0.5 $10^9$/L) | 0.02 (0–0.06) | 0 (0–0.01) | 0 (0–0.01) | 0 (0–0) | **8.14E-16** | **7.26E-17** | **5.33E-19** | 0.096 | **5.00E-03** | 0.328 | <0.001 |
| Basophils (0.0–0.2 $10^9$/L) | 0.02 (0.01–0.03) | 0.02 (0.01–0.02) | 0.02 (0.01–0.03) | 0.02 (0.01–0.03) | **6.68E-04** | 0.018 | 0.021 | 0.138 | 0.121 | 0.945 | 0.003 |
| NLR | 2.64 (1.60–3.61) | 4.27 (2.76–6.41) | 6.53 (4.02–10.85) | 8.11 (4.3–12.51) | **1.44E-09** | **4.26E-24** | **1.49E-28** | **9.14E-19** | **7.17E-26** | 0.098 | <0.001 |

| | | | | | | | | | | | |
|---|---|---|---|---|---|---|---|---|---|---|---|
| Platelets (140–400 $10^9$/L) | 223 (167–285) | 200 (158–252) | 200 (154–256) | 175 (133–229) | **2.60E-02** | 0.017 | **5.44E-06** | 0.517 | **1.76E-06** | **0.001** | <0.001 |
| **Clinical Chemistry & Immunology** | | | | | | | | | | | |
| **APR and related parameters** | | | | | | | | | | | |
| CRP (0.03–0.5 mg/dL) | 1.49 (0.50–4.62) | 7.46 (3.18–13.16) | 15.53 (7.91–22.43) | 17.21 (9.76–25.08) | **2.05E-12** | **1.73E-30** | **2.64E-33** | **9.42E-21** | **7.09E-23** | 0.116 | **<0.001** |
| IL-6 (0.0–4.3 pg/mL) | 10 (4–25) | 37 (21–63) | 80 (47–127) | 90 (55–163) | **8.92E-13** | **5.41E-35** | **7.07E-38** | **2.09E-27** | **1.21E-28** | 0.092 | **<0.001** |
| Ferritin (25–400 ng/mL) | 247 (104.5–441) | 501 (269–935) | 795 (474–1478) | 671 (334–1217) | **3.67E-08** | **2.12E-17** | **3.10E-10** | **2.23E-10** | **0.007** | 0.045 | **<0.001** |
| LDH (0–248 UI/L) | 243 (209–284) | 318 (267–391) | 434 (330–564) | 429 (342–523) | **3.11E-11** | **3.43E-29** | **2.32E-24** | **6.30E-21** | **4.48E-13** | 0.902 | **<0.001** |
| Triglycerides (43–200 mg/dL) | 121 (89.7–159.5) | 120 (91–158.5) | 122.5 (93.5–172.8) | 126.5 (93.5–158.8) | 0.953 | 0.723 | 0.681 | 0.533 | 0.558 | 0.897 | 0.882 |
| Coagulation factors | | | | | | | | | | | |
| Fibrinogen (2.39–6.1 g/L) | 4.37 (4.00–4.83) | 5.12 (4.48–5.92) | 5.45 (4.73–6.28) | 4.94 (4.23–5.90) | **2.96E-09** | **1.26E-13** | **8.47E-06** | **2.04E-04** | 7.95E-02 | **1.48E-05** | **<0.001** |
| D dimer (0–243 ng/mL) | 168 (115–284) | 241 (160–401) | 303 (192–549) | 477 (292.5–861) | **9.90E-04** | **3.24E-08** | **3.58E-17** | **3.78E-06** | **4.93E-20** | **1.38E-05** | **<0.001** |
| Prothrombin time INR (0.7–1.3) | 1.03 (0.99–1.11) | 1.1 (1.03–1.17) | 1.11 (1.03–1.20) | 1.14 (1.05–1.30) | **4.17E-05** | **2.89E-05** | **2.33E-10** | 0.423 | 1.38E-06 | **0.001** | **<0.001** |
| **ORGAN DAMAGE RELATED BIOMARKERS (ODBR)** | | | | | | | | | | | |
| **Liver function test** | | | | | | | | | | | |
| AST (12–50 IU/L) | 27 (22–38) | 38 (29–56) | 47 (35–71) | 45 (31–68) | **2.20E-07** | **3.27E-14** | **6.19E-11** | **5.45E-08** | **6.48E-04** | 0.117 | **<0.001** |
| ALT (19–50 IU/L) | 22 (13–42) | 29 (19–51) | 34 (22–54) | 22.5 (15–35) | **1.80E-03** | **6.13E-05** | 0.993 | 0.030 | **3.37E-08** | **7.53E-10** | **<0.001** |
| AST/ALT (0.5–1) | 1.30 (0.87–1.69) | 1.30 (1.00–1.71) | 1.47 (1.14–1.86) | 1.92 (1.43–2.60) | 0.427 | 0.008 | 2.14E-12 | **2.00E-04** | **8.10E-33** | **1.37E-11** | **<0.001** |
| Direct bilirubin (0.1–0.57 mg/dL) | 0.25 (0.22–0.32) | 0.3 (0.24–0.37) | 0.31 (0.25–0.42) | 0.35 (0.27–0.46) | **4.76E-04** | **1.39E-05** | **7.14E-08** | 0.033 | **1.27E-04** | 0.067 | **<0.001** |
| Total bilirubin (0.3–1.2 mg/dL) | 0.49 (0.4–0.61) | 0.57 (0.45–0.74) | 0.57 (0.42–0.775) | 0.62 (0.47–0.85) | **4.50E-03** | 0.017 | **2.28E-04** | 0.726 | **4.16E-02** | 0.048 | 0.002 |
| **Kidney function test** | | | | | | | | | | | |
| Urea (17–43 mg/dL) | 29.5 (23.0–35.7) | 32 (24.0–45.0) | 36 (26.0–50.0) | 58 (42.0–87.0) | 0.077 | **0.001** | **1.30E-20** | **2.50E-03** | **6.95E-47** | **1.92E-19** | <0.001 |

| | | | | | | | | | | | |
|---|---|---|---|---|---|---|---|---|---|---|---|
| Creatinine (0.67–1.17 mg/dL) | 0.72 (0.59–0.86) | 0.78 (0.64–0.93) | 0.85 (0.70–1.02) | 1.01 (0.78–1.36) | 0.030 | **1.82E-05** | **6.43E-14** | **1.02E-05** | **1.12E-24** | **6.32E-07** | <0.001 |
| GFR (>75 mL/1.73 $m^2$) | 90 (90–90) | 90 (88.–90) | 90 (79–90) | 57 (36–79) | 0.206 | **5.75E-04** | **3.06E-65** | **2.36E-06** | **1.42E-196** | **1.5E-112** | <0.001 |

AST, Aspartic Amino Transferase; ALT, Alanine Amino Transferase; CRP; C Reactive Protein; GFR, Glomerular Filtration Rate; Hb, Hemoglobin; NLR, Neutrophil Lymphocyte Ratio; WBC, White Blood cell Count.

**Table 6S. Median values of laboratory variables and the proportion out of the normal range and relation with mortality**

| | n | Median (IQR) | ONR<br>n (%) | Mortality<br>n (%) | *p*-value | OR |
|---|---|---|---|---|---|---|
| **Blood** | | | | | | |
| Hb | 1550 | 13.5 (12.3–14.5) | 590 (38.06%) | 107 (18.1%) | n.s. | |
| <12 | | | 272 | 83 (30.5%) | 3.00E-10 | 2.77 |
| >15 | | | 246 | 21 (8.5%) | 1.16E-04 | 2.38 |
| Leukocytes | 1550 | 6.6 (5.08–8.76) | 344 (22.19%) | 86 (25.0%) | 5.63E-06 | 2.00 |
| <4.0 e3 | | | 166 | 28 (16.9%) | n.s. | |
| >11.0 e3 | | | 178 | 58 (32.6%) | 2.30E-08 | 2.83 |
| Neutrophils, % | 1550 | 76.1 (68–83.2) | 550 (35.48%) | 153 (27.8%) | 1.00E-10 | |
| <40% | | | 9 | 1 (11.1%) | n.s. | |
| >80% | | | 541 | 152 (28.1%) | 1.00E-10 | 3.33 |
| Top 10% cutoff | | | 156 | 58 (37.2%) | 1.00E-10 | 3.53 |
| Bottom 10% cutoff | | | 157 | 15 (9.6%) | 1.24E-02 | 2.00 |
| Neutrophils, n | 1550 | 4.9 (3.5–6.8 ) | 415 (26.77%) | 114 (27.5%) | 1.00E-10 | 2.61 |
| <2.0 e3 | | | 43 | 8 (18.6%) | n.s. | |
| >7.0 e3 | | | 372 | 106 (28.5%) | 1.0E-10 | 2.69 |
| Top 10% cutoff, 88.4% | | | 156 | 58 (37.2%) | 1.0E-10 | 3.53 |
| Top 5% cutoff, 90.6 | | | 77 | 36 (46.8%) | 2.0E-10 | 4.95 |
| Lymphocytes, % | 1550 | 16.0 (10.5–23.0) | 1031 (66.52%) | 211 (20.5%) | 3.7E-09 | 2.58 |
| <20% | | | 1022 | 210 (20.5%) | 2.8E-09 | 2.59 |
| >50% | | | 9 | 1 (11.1%) | n.s. | |
| Bottom 10% | | | 157 | 54 (34.4%) | n.s. | |
| Bottom 5% | | | 77 | 34 (44.2%) | 2.58E-02 | 1.61 |
| Lymphocytes, n | 1550 | 1.0 (0.7–1.4) | 993 (64.06%) | 207 (20.8%) | 1.1E-09 | 2.61 |
| <1.2 e3 | | | 985 | 206 (20.9%) | 8.00E-10 | 2.61 |
| >3.5 e3 | | | 8 | 1 (12.5%) | n.s. | |
| Bottom 10% | | | 163 | 60 (36.8%) | 1.00E-10 | 3.50 |
| Bottom 5% | | | 87 | 32 (36.8%) | 3.95E-06 | 3.18 |
| Monocytes, % | 1551 | 6.7 (4.8–8. 8) | 184 (11.86%) | 37 (20.1%) | n.s. | |
| <2% | | | 25 | 12 (48.0%) | 2.39E-04 | 4.80 |
| >11% | | | 159 | 25 (15.7%) | n.s. | |
| <0.1 e3 | | | 1 | 1 (100.0%) | n.s. | |
| >1.0 e3 | | | 67 | 16 (23.9%) | n.s. | |
| Eosinophils, % and n | 1551 | 0.0 (0–0.3) | 16 (1.03%) | 2 (12.5%) | n.s. | |
| >5% | | | 11 | 1 (9.1%) | n.s. | |
| >0.05 e3 | | | 5 | 1 (20.0%) | n.s. | |
| Basophils, n | 1551 | 0.0 (0–0) | 1 | 1 (100.0%) | n.s. | |

| | | | | | | |
|---|---|---|---|---|---|---|
| >0.02 e3 | | 0.02 (0.01 -0.03) | 1 | 1 (100.0%) | n.s. | |
| Platelets | 1550 | | 311 | 83 (26.7%) | 3.96E-04 | 2.21 |
| <140 e3 | | | 251 | 72 (28.7%) | 1.46E-04 | 2.41 |
| >400 e3 | | | 60 | 11 (18.3%) | n.s. | |
| **APR and related parameters** | | | | | | |
| CRP (N 0.03-0.5 mg/dL) | 1226 | 8.9 (3.8–16.6 ) | 1219 (99.43%) | 146 (11.98%) | n.s. | |
| Top decile cutoff, 20 | | | 122 | 37 (30.33%) | 6.40E-09 | 3.93 |
| Top 5% cutoff, 30 | | | 62 | 24 (38.71%) | 2.44E-08 | 5.35 |
| IL-6 (0.0–4.3 pg/mL) | 1259 | 45.0 (24–80) | 1029 (81.73%) | 146 (14.19%) | 1.07E-02 | Infinity |
| Top decile cutoff, 140 | | | 125 | 47 (37.60%) | 1.00E-10 | 6.30 |
| Top 5% cutoff, 30 219.5 | | | 62 | 24 (38.71%) | 1.24E-08 | 5.57 |
| Ferritin (25-400 ng/mL) | 1128 | 539.0 (283.5–1011) | 732 (64.89%) | 77 (10.52%) | n.s. | |
| Top decile cutoff, 1667 | | | 108 | 12 (11.11%) | n.s. | |
| Top 5% cutoff, 2279 | | | 57 | 10 (17.54%) | n.s. | |
| Triglycerides (43-200 mg/dL) | 620 | 121.0 (92 -161) | 78 (12.58%) | 6 (7.69%) | n.s. | |
| Top 10% cutoff, 224 mg/dL | | | 63 | 4 (6.35%) | n.s. | |
| Top 5% cutoff, 269 mg/dL | | | 50 | 1 (2.00%) | n.s. | |
| LDH (0-248 UI/L) | 1128 | 336.0 (271–421) | 935 (82.89%) | 98 (10.48%) | 0.04 | 1.94 |
| Top 5% cutoff, 661 UI/L | | | 56 | 14 (25.00%) | 4.9E-04 | 3.43 |
| **Coagulation parameters** | | | | | | |
| Fibrinogen (2.39–6.1 g/L) | 1440 | 5.1 (4.43–5.95) | 311 (21.60%) | 48 (15.43%) | n.s. | |
| >6.1 | | | 310 | 48 (15.48%) | n.s. | |
| <2.4 | | | 2 | 0 (0.00%) | n.s. | |
| Top decile cutoff, 6.7 | | | 146 | 28 (19.18%) | n.s. | |
| Top 5% cutoff, 7.22 | | | 72 | 13 (18.06%) | n.s. | |
| Bottom 5% cutoff, 1.68 | | | 73 | 18 (24.66%) | 0.0495 | |
| D-dimer (0-243 ng/mL) | 1248 | 253.0 (168–463) | 683 (54.73%) | 121 (17.72%) | 1.0E-10 | 4.65 |
| Top decile cutoff, 1074 ng/mL | | | 124 | 28 (22.58%) | 3.1E-04 | 2.49 |
| Top 5% cutoff, 2352 ng/mL | | | 62 | 17 (27.42%) | 4.0E-04 | 3.10 |
| Prothrombin time, INR (0.7-1.3) | 1445 | 1.1 (1.03 -1.19) | 148 (10.24%) | 57 (38.51%) | 1.0E-09 | 3.99 |
| Top decile cutoff, 1.31 | | | 148 | 57 (38.51%) | 1.0E-10 | 3.99 |
| Top 5% cutoff, 1.51 | | | 72 | 30 (41.67%) | 9.3E-08 | 4.12 |
| Bottom 5% | | | 84 | 12 (14.29%) | n.s. | |
| **Liver function test** | | | | | | |
| AST (12-50 IU/L) | 1560 | 40.0 (30–60) | 532 (34.10%) | 109 (20.49%) | 4.1E-03 | 1.50 |
| Top decile cutoff, 92 IU/L | | | 156 | 39 (25.00%) | 4.6E-03 | 1.78 |
| Top 5% cutoff, 116 | | | 79 | 17 (21.52%) | n.s. | |
| ALT (19–50 IU/L) | 1563 | 28.0 (1911 -50 ) | 370 (23.67%) | 41 (11.08%) | 1.1E-03 | 0.56 |

| | | | | | | |
|---|---|---|---|---|---|---|
| Top decile cutoff, 75 UI/dL | | | 162 | 18 (11.11%) | 3.3E-02 | 0.56 |
| Top 5% cutoff, 106 IU/L | | | 80 | 5 (6.25%) | 8.2E-03 | 0.32 |
| AST/ALT | 1560 | 1.4 (1.05- 1.8) | 1213 (77.76%) | | | |
| Top decile cutoff, 2.4 | | | 155 | 74 (47.74%) | | |
| Bilirubin, T (0.3-1.2 mg/dL) | 1022 | 0.6 (0.45 -0.74) | 46 (4.50%) | 12 (26.09%) | n.s. | |
| Top decile cutoff, 0.95 mg/dL | | | 103 | 22 (21.36%) | 5.7E-04 | 2.63 |
| Top 5% cutoff | | | 51 | 11 (21.57%) | 1.7E-02 | 2.48 |
| Bilirubin, D (0.1-0.57 mg/dL) | 982 | 0.3 (0.24–0.38) | 49 (4.99%) | 10 (20.41%) | 2.0E-02 | 2.63 |
| Top decile cutoff, 0.64 mg/dL | | | 98 | 20 (20.41%) | 4.0E-04 | 2.85 |
| Top 5% cutoff | | | 49 | 10 (20.41%) | 2.0E-02 | 2.63 |
| **Kidney function test** | | | | | | |
| Urea (17-42 mg/dL) | 1291 | 35.0 (35 51) | 627 (48.57%) | 169 (26.95%) | 1.0E-10 | 4.01 |
| Top decyl cutoff, 78 mg/dL | | | 132 | 78 (59.09%) | 1.0E-10 | 9.94 |
| Top 5% cutoff, 103 mg/dL | | | 66 | 41 (62.12%) | 1.0E-10 | 9.28 |
| Bottom 5% cutoff, 5 mg/dL | | | 72 | 3 (4.17%) | 1.1E-03 | 5.12 |
| Creatinine >1.17 | 1576 | | 270 (17.13%) | 107 (39.63%) | 1.6E-26 | 5.34 |
| Creatinine, top decile cutoff, 1.42 | | | 157 | 74 (47.13%) | 4.2E-23 | 6.35 |
| GFR <75 | 1573 | | 499 (31.7%) | 326 (65.33%) | 4.4E-39 | 6.42 |
| Bottom 10%, <42.7 mL | | | 154 | 83 (53.90%) | 6.5E-31 | 8.53 |

Clinical laboratory variables and the 28-day decease outcome. Mortality among patients who had abnormal and extreme deviations in the laboratory values. The WBC count was at the above upper limit of normal (ULN) in 22.2% of the patients, and the mortality rate of these patients was significantly higher (25.0%) than that of the entire study cohort (16.1%). In most patients, the platelet count was within the normal range; however, when values fell below the lower limit of normal (LLN), the mortality rate was 26.7%, which was significantly higher than that of the HUVH cohort's. The CRP values were outside the normal range in all but one patient; the mortality rate was 30.3% among patients with values in the top 10% (>20 mg/dL), which was significantly higher than the HUVH cohort's. The IL-6 values were above the ULN in 96.8% of cases; the mortality rate was 37.7% in the participants with the top 10% values (>140 ng/dL). LDH and Triglycerides were excluded from other analysis because the excessive and unbalance number or missing data. Comparisons by Fisher exact test. APR, acute-phase reactants; AST, aspartate aminotransferase; ALT, alanine aminotransferase; CRP, C-reactive protein; DFSO, days from symptom onset; GFR, glomerular filtration rate; IL-6, Interleukin-6; Hb, hemoglobin; LDH, lactate dehydrogenase; LOS, length of stay; NLR, neutrophil-to-lymphocyte ratio; OR, Odd Ratio; ONR, out of the normal range.

#### Table 7S. ROC curve analysis, age stratified

| Variable | AUC 40-55 years | 95% CI | *p* | n | AUC 56-65 years | 95% CI | *p* | n | AUC 66-75 years | 95% CI | *p* | n | AUC 76-85 years | 95% CI | *p* | n | AUC >85 years | 95% CI | *p* | n |
|---|---|---|---|---|---|---|---|---|---|---|---|---|---|---|---|---|---|---|---|---|
| Age | 0.63 | 0.56-0.68 | 5.68E-05 | 417 | 0.58 | 0.50-0.66 | 3.72E-02 | 281 | 0.53 | 0.45-0.59 | 4.38E-01 | 306 | 0.58 | 0.50-0.64 | 3.89E-02 | 247 | 0.59 | 0.56-0.68 | 8.50E-03 | 320 |
| Comorbidity | 0.66 | 0.60-0.71 | 2.96E-07 | 417 | 0.57 | 0.50-0.66 | 8.56E-02 | 281 | 0.63 | 0.56-0.68 | 2.33E-04 | 306 | 0.59 | 0.51-0.65 | 1.91E-02 | 247 | 0.64 | 0.60-0.71 | 3.63E-05 | 320 |
| SpO2/FiO2 | **0.76** | 0.69-0.82 | 6.85E-11 | 223 | **0.77** | 0.68-0.85 | 3.70E-02 | 132 | **0.79** | 0.71-0.86 | 3.82E-10 | 159 | 0.72 | 0.63-0.80 | 4.37E-06 | 146 | 0.75 | 0.69-0.82 | 1.20E-08 | 184 |
| Hb | 0.55 | 0.48-0.61 | 1.27E-01 | 411 | 0.58 | 0.49-0.65 | 3.05E-07 | 286 | 0.54 | 0.47-0.60 | 2.73E-01 | 304 | 0.58 | 0.50-0.64 | 3.98E-02 | 247 | 0.56 | 0.48-0.61 | 1.14E-01 | 315 |
| NLR | **0.72** | 0.66-0.77 | 3.02E-12 | 411 | 0.66 | 0.58-0.73 | 5.53E-05 | 277 | 0.69 | 0.62-0.75 | 3.63E-08 | 304 | 0.65 | 0.61-0.75 | 2.72E-05 | 247 | 0.71 | 0.66-0.77 | 9.76E-10 | 315 |
| Monocytes % | 0.64 | 0.57-0.69 | 1.81E-05 | 411 | 0.65 | 0.57-0.72 | 1.85E-04 | 277 | 0.69 | 0.62-0.75 | 6.28E-08 | 304 | 0.69 | 0.53-0.67 | 3.33E-07 | 247 | 0.64 | 0.57-0.69 | 3.82E-05 | 315 |
| Monocytes n | 0.58 | 0.51-0.64 | 1.22E-02 | 411 | 0.56 | 0.48-0.63 | 1.32E-01 | 277 | 0.54 | 0.47-0.61 | 2.00E-01 | 304 | 0.61 | 0.51-0.65 | 3.60E-03 | 247 | 0.59 | 0.51-0.64 | 7.60E-03 | 315 |
| Eosinophils % | 0.64 | 0.58-0.69 | 1.08E-05 | 411 | 0.60 | 0.52-0.67 | 1.56E-02 | 277 | 0.60 | 0.53-0.65 | 5.80E-03 | 304 | 0.59 | 0.51-0.65 | 1.61E-02 | 247 | 0.63 | 0.58-0.69 | 1.65E-04 | 315 |
| CRP | **0.72** | 0.66-0.77 | 1.85E-10 | 359 | 0.73 | 0.65-0.80 | 2.83E-07 | 235 | 0.76 | 0.69-0.82 | 2.75E-11 | 240 | 0.73 | 0.64-0.80 | 6.35E-07 | 165 | 0.73 | 0.66-0.77 | 1.40E-09 | 274 |
| IL6 | **0.78** | 0.72-0.83 | 1.16E-16 | 374 | **0.76** | 0.68-0.82 | 3.32E-09 | 245 | **0.78** | 0.72-0.84 | 5.27E-14 | 253 | 0.72 | 0.64-0.80 | 1.24E-06 | 159 | 0.79 | 0.72-0.83 | 6.24E-15 | 287 |
| Ferritin | 0.65 | 0.58-0.71 | 3.41E-05 | 343 | 0.67 | 0.58-0.76 | 2.31E-04 | 225 | 0.61 | 0.53-0.68 | 9.30E-03 | 214 | 0.61 | 0.51-0.70 | 2.43E-02 | 145 | 0.65 | 0.58-0.71 | 1.91E-04 | 260 |
| D-dimer | 0.65 | 0.59-0.71 | 1.03E-05 | 343 | 0.64 | 0.55-0.71 | 1.40E-03 | 245 | 0.62 | 0.55-0.69 | 1.70E-03 | 245 | 0.64 | 0.55-0.72 | 1.90E-03 | 164 | 0.64 | 0.59-0.71 | 2.54E-04 | 279 |
| AST/ALT | 0.61 | 0.55-0.67 | 3.90E-04 | 412 | 0.59 | 0.51-0.66 | 2.12E-02 | 280 | 0.64 | 0.56-0.70 | 9.90E-05 | 298 | 0.61 | 0.54-0.68 | 2.70E-03 | 244 | 0.59 | 0.55-0.67 | 1.05E-02 | 315 |
| GFR | 0.63 | 0.56-0.68 | 5.12E-05 | 416 | 0.57 | 0.49-0.64 | 8.09E-02 | 281 | 0.65 | 0.58-0.71 | 1.03E-05 | 305 | 0.61 | 0.53-0.67 | 4.00E-03 | 245 | 0.62 | 0.56-0.68 | 8.87E-04 | 319 |

AST, Aspartic Amino Transferase; ALT, Alanine Amino Transferase; CRP; C Reactive Protein; GFR, Glomerular Filtration Rate; Hb, Hemoglobin; NLR, Neutrophil Lymphocyte Ratio; NC, not calculated, SpO2/FiO2, Oxygen saturation to fraction of inspired oxygen ratio; WBC, White Blood cell Count; CI, Confidence interval 5 to 95%.

Table 8S. Random Forest model applied to HUVH cohort

| Random forest models | | | | | |
|---|---|---|---|---|---|
| **Laboratory data model** | | **Clinical data model** | | **Laboratory and clinical data combined model** | |
| **Variables** | **Mean Decrease Gini** | **Variables** | **Mean Decrease Gini** | **Variables** | **Mean Decrease Gini** |
| IL-6 | 12.3 | Age, years | 40.5 | Age | 13.8 |
| CRP | 10.4 | Comorbidity Index | 11.1 | IL-6 | 9.7 |
| GFR | 8.5 | Sex | 5.6 | CRP | 7.1 |
| Neutrophils, % | 7.6 | Cardiovascular conditions | 5.2 | GFR | 6.4 |
| Urea | 7.3 | Obesity | 4.1 | Lymphocytes, n | 6.3 |
| AST/ALT ratio | 7.0 | Chronic lung disease | 3.5 | Neutrophils, % | 5.5 |
| NLR | 6.5 | Diabetes | 3.5 | Urea | 5.3 |
| Lymphocytes, n | 6.5 | Chronic neurological disease | 3.2 | Lymphocytes, % | 5.2 |
| Fibrinogen | 6.4 | Chronic kidney disease | 2.4 | AST/ALT ratio | 5.2 |
| Creatinine | 6.2 | Active non-terminal malignancy | 2.3 | NLR | 5.2 |
| Lymphocytes, % | 5.9 | Chronic liver disease | 1.2 | Creatinine | 4.7 |
| Creatinine | 5.7 | | | D-dimer | 4.6 |
| Platelets | 5.3 | | | Monocytes, % | 4.4 |
| Ferritin | 5.2 | | | Fibrinogen | 4.4 |
| Monocytes, % | 5.2 | | | Ferritin | 4.3 |
| AST | 5.2 | | | Platelets | 4.0 |
| Hb | 5.1 | | | AST | 4.0 |
| ALT | 5.1 | | | Hb | 3.8 |
| Prothrombin time, INR | 4.7 | | | Monocytes, n | 3.8 |
| Monocytes, n | 4.5 | | | Prothrombin time, INR | 3.7 |
| WBC | 4.3 | | | ALT | 3.6 |
| Neutrophils, n | 4.2 | | | Neutrophils, n | 3.6 |
| Eosinophils, % | 3.2 | | | WBC | 3.3 |
| Eosinophils, n | 2.4 | | | Comorbidity Index | 2.9 |
| | | | | Eosinophils, % | 2.8 |
| | | | | Eosinophils, n | 2.6 |
| | | | | Cardiovascular conditions | 1.6 |
| | | | | Obesity | 0.9 |
| | | | | Chronic lung disease | 0.7 |
| | | | | Chronic neurological disease | 0.6 |
| | | | | Sex | 0.8 |
| | | | | Active non-terminal malignancy | 0.4 |
| | | | | Chronic kidney disease | 0.2 |
| | | | | Chronic liver disease | 0.125 |

AST, Aspartic Amino Transferase; ALT, Alanine Amino Transferase; CRP; C Reactive Protein; GFR, Glomerular Filtration Rate; Hb, Hemoglobin; NLR, Neutrophil Lymphocyte Ratio; NC, not calculated, SpO2/FiO2, Oxygen saturation to fraction of inspired oxygen ratio; WBC, White Blood Cell count.
For the HUVH simulation the training dataset has 1264 samples and 51 features, and the test dataset had 315 samples and 51 features. Variables with more than 20% missing values were removed, missing value imputation by median. The Mean Decrease Gini value indicates the importance of the variable in the outcome predicted in the model.

**Table 9S. Random Forest Model; comparison exploratory and combination of the three cohorts.**

| | Mean Decrease Gini | | | |
|---|---|---|---|---|
| **Variables** | HUVH cohort | Rank | Combined cohorts | Rank |
| Urea | 16.7 | 1 | 31.3 | 2 |
| IL-6 | 12.8 | 2 | 40.3 | 1 |
| D.dimer | 11.4 | 3 | 27.6 | 3 |
| CRP | 8.4 | 4 | 20.6 | 9 |
| Hb | 7.0 | 5 | 22.5 | 6 |
| ALT | 6.3 | 6 | 25.2 | 4 |
| Platelets | 6.2 | 7 | 21.2 | 8 |
| Neutrophils % | 6.0 | 8 | 23.5 | 5 |
| NLR | 5.8 | 9 | 22.1 | 7 |
| Lymphocytes n | 5.8 | 10 | 20.3 | 10 |
| Lymphocytes % | 5.6 | 11 | 19.3 | 11 |
| Ferritin | 5.4 | 12 | 17.8 | 13 |
| Monocytes n | 4.7 | 13 | 14.3 | 16 |
| Protrombin (INR) | 4.6 | 14 | 15.7 | 14 |
| Monocytes % | 4.6 | 15 | 18.1 | 12 |
| Neutrophils n | 4.4 | 16 | 15.6 | 15 |
| Eosinophils % | 2.7 | 17 | 6.5 | 18 |
| Basophils n | 2.5 | 18 | 7.3 | 17 |
| Eosinophils n | 2.3 | 19 | 5.3 | 19 |

CRP, C Reactive Protein; Hb, Hemoglobin; NLR, Neutrophil Lymphocyte Ratio; WBC, White Blood cell Count; n, number. For the three cohorts' simulation the training dataset had 2600 samples, and the test dataset 519 samples and 19 features. Variables with more than 20% missing values were removed, missing values imputation by median. The Mean Decrease Gini value indicates the importance of the variable in the outcome predicted in model.

**Table 10S. Patients included in immunological studies, cytokines and mononuclear cell phenotype by flowcytometry**

| Patients | HUVH cohort (n = 1579) | Cytokines sub-cohort (n = 74) | *p*-value | Phenotype sub-cohort (n = 41) | *p*-value |
|---|---|---|---|---|---|
| Age median (IQR) | 62 (50–75) | 53 (44–63) | <0.0001 | 50 (41–65) | 0.012 |
| Female n (%) | 698 (44.2%) | 36 (48.70%) | 0.45 | 15 (30.0%) | 0.34 |
| Male n (%) | 880 (55.7%) | 38 (51.40%) | | 26 (52.0%) | |
| **Mortality** | | | | | |
| Overall n (%) | 255 (16.1%) | 1 (1.40%) | <0.001 | 3 (7.3%) | 0.18 |
| Female n (%) | 107 (15.3%) | 0 (0) | <0.01 | 1 (6.7%) | 0.48 |
| Male n (%) | 148 (16.8%) | 1 (2.6%) | <0.05 | 2 (7.7%) | 0.29 |
| DFSO median (IQR) | 7 (4–10) | 7 (5–9) | 0.73 | 7 (5–10) | 0.09 |
| LOS median (IQR) | 6 (2–19) | 11 (6–37) | <0.001 | 6 (2–16) | 0.97 |
| **Severity** | | | | | |
| Mild n (%) | 71 (4.40%) | 0 (0.0%) | <0.0001 | 5 (12.2%) | 0.06 |
| Moderate n (%) | 969 (61.50%) | 48 (64.9%) | | 27 (65.9%) | |
| Severe n (%) | 284 (17.40%) | 25 (33.8%) | | 6 (14.6%) | |
| Deceased n (%) | 255 (16.80%) | 1 (1.4%) | | 3 (7.3%) | |
| **Clinical presentation** | | | | | |
| Fever n (%) | 1325 (83.9%) | 71 (96.0%) | <0.01 | 36 (87.8%) | 0.13 |
| Respiratory symptoms | | | | | |
| Upper airways symptoms n (%) | 94 (5.9 %) | 4 (5.4%) | 0.99 | 3 (7.3%) | 0.73 |
| Lower airway symptoms n (%) | 1351 (85.5 %) | 64 (86.5%) | 0.99 | 34 (82.9%) | 0.65 |
| Pneumonia n (%) | 1525 (96.5 %) | 74 (1) | 0.17 | 30 (73.2%) | <0.0001 |
| Digestive n (%) | 492 (31.1 %) | 28 (37.8%) | 0.24 | 15 (36.6%) | 0.45 |
| **Comorbidities** | | | | | |
| Cardiovascular & hypertension n (%) | 713 (45.1 %) | 23 (31.1%) | 0.02 | 15 (36.6%) | 0.34 |
| Chronic lung disease n (%) | 278 (17.6 %) | 10 (13.5%) | 0.43. | 3 (7.3%) | 0.09 |
| Diabetes n (%) | 293 (18.5 %) | 8 (10.8%) | 0.12 | 11 (26.8%) | 0.03 |
| Neurological disease n (%) | 227 (14.4 %) | 5 (6.8%) | 0.08 | 3 (7.3%) | 0.25 |
| Chronic renal disease n (%) | 134 (8.5 %) | 1 (1.4%) | 0.02 | 1 (2.4%) | 0.25 |
| Active non-terminal malignancy n (%) | 113 (7.2 %) | 5 (6.8%) | 0.99 | 0 (0.0%) | 0.11 |
| Obesity n (%) | 261 (16.5 %) | 24 (32.4%) | 1 | 2 (4.9%) | 0.05 |
| Chronic liver disease n (%) | 61 (3.9 %) | 1 (1.4%) | 0.52 | 3 (7.3%) | 0.21 |
| Comorbidity index (mean ± SD) | 1.32 (±1.3) | 1 (±1.3) | 0.03 | 0.92 (1.14) | 0.04 |
| **Blood tests** | | | | | |
| Neutrophils, % median (IQR) | 76.1 (68–83.2) | 73.9 ( 68.4–82.3) | 0.69 | 79.6 (70.6–84.5) | 0.25 |
| Neutrophils, n median (IQR) | 4.9 (3.5– 6.9) | 4.7 (3.3–6.3) | 0.37 | 5.8 (4.1–7.7) | 0.17 |
| Lymphocytes, % median (IQR) | 16 (10.5–23) | 17.5 (12.8–17.5) | 0.14 | 13 (10.5–20.6) | 0.32 |

| | | | | | |
|---|---|---|---|---|---|
| Lymphocytes, n median (IQR) | 1.0 (0.7–1.4) | 1.1 (0.83–1.48) | 0.23 | 1.02 (0.77–1.02) | 0.63 |
| Monocytes, n median (IQR) | 0.4 (0.31–0.6) | 0.39 (0.29–0.58) | 0.34 | 0.44 (0.33–0.63) | 0.50 |
| Eosinophils, % median (IQR) | 0 (0–0.3) | 0.0 (0.0–0.2) | 0.45 | 0.10 (0.0–0.45) | 0.13 |
| Platelets median (IQR) | 197 (154–251) | 199 (167–199) | 0.46 | 229 (176–306) | 0.007 |
| **Clinical Chemistry & Immunology** | | | | | |
| A**PR and related parameters** | | | | | |
| CRP median (IQR) | 8.9 (3.8–16.6) | 7.8 (4.5 -18) | 0.93 | 9.9 (6.2–15) | 0.97 |
| IL-6 median (IQR) | 45.1 (23.80.0) | 37 (21–78) | 0.24 | 46 (25–79) | 0.86 |
| Ferritin median (IQR) | 539 (282.5–1011.5) | 581 (284–1,106) | 0.44 | 438 (237–915) | 0.63 |
| **Coagulation** | | | | | |
| D-dimer median (IQR) | 263 (168–463.5) | 246 (141–420) | 0.23 | 239 (152–375) | 0.25 |

HUVH Cohort: clinical and demographic features of the sub-cohorts from cytokines and blood cell-phenotypic analysis. Univariate comparison using Mann–Whitney U test with adjusted p-values. DFSO, days from symptom onset; LOS, length of stay; NLR, neutrophil-to-lymphocyte ratio; CRP, C-reactive protein; AST, aspartate aminotransferase; ALT, alanine aminotransferase; eGFR, estimated glomerular filtration rate; IL-6, Interleukin-6; LDH, lactate dehydrogenase; Hb, hemoglobin.

#### SUPPLEMENTARY FIGURES AND LEGENDS

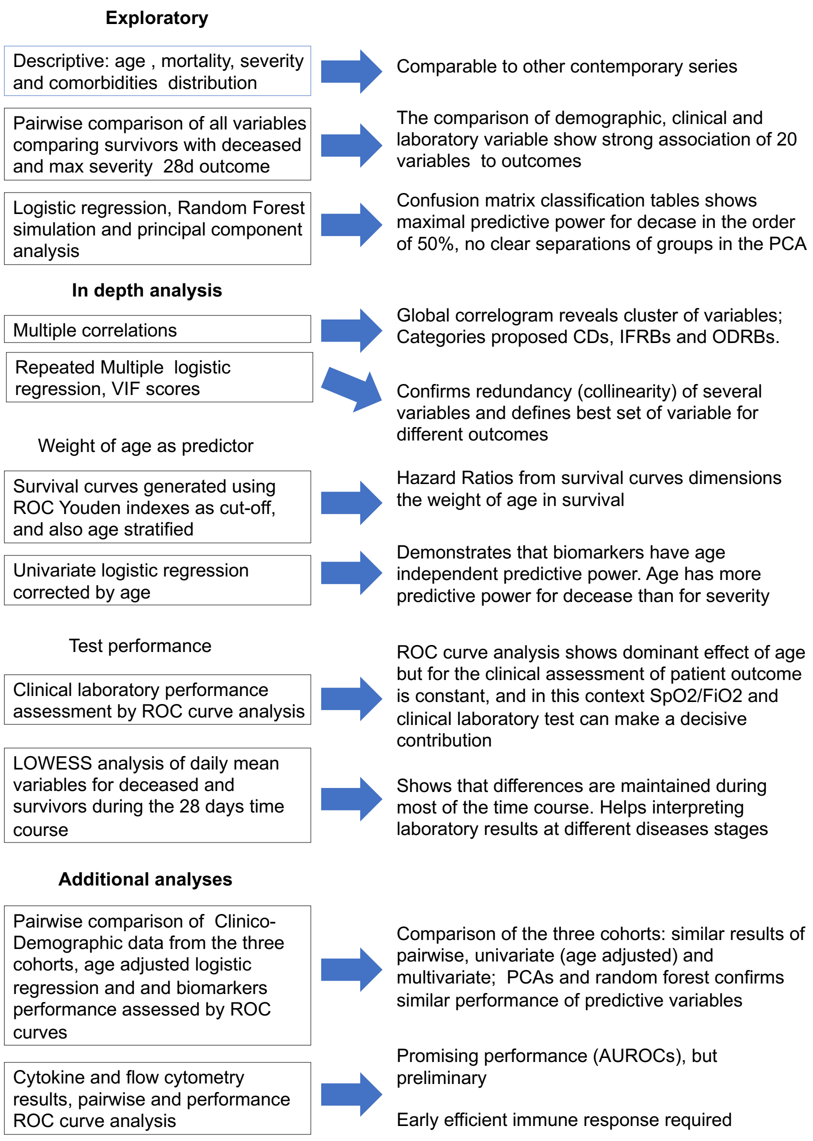

Figure 1 S

Figure 1S. Sequence of statistical analyses and summary of conclusions from every step, left column boxes type of analysis, right column boxes main conclusion. CD, Clinicodemographic; IFRB, Inflammatory Related Biomarkers; ODRB, Organ Damage Related Biomarkers.

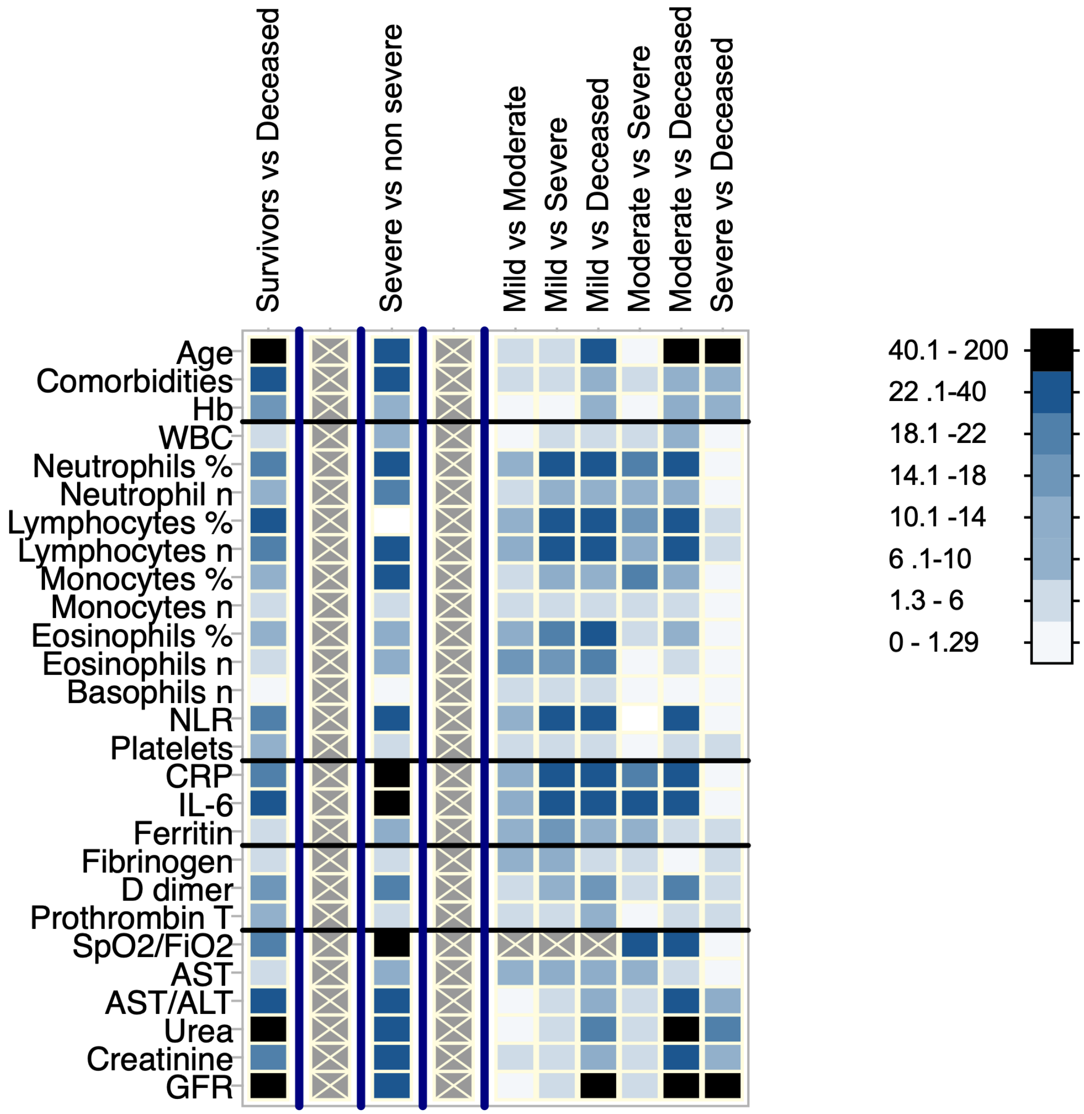

Figure 2S. Heatmap summarizing pairwise comparison for HUVH cohort; *p* values scale refers to non-parametric tests (Mann-Whitney and Kruskal-Wallis). The colour scale refers to the log of exact p x -1 to generate positive values

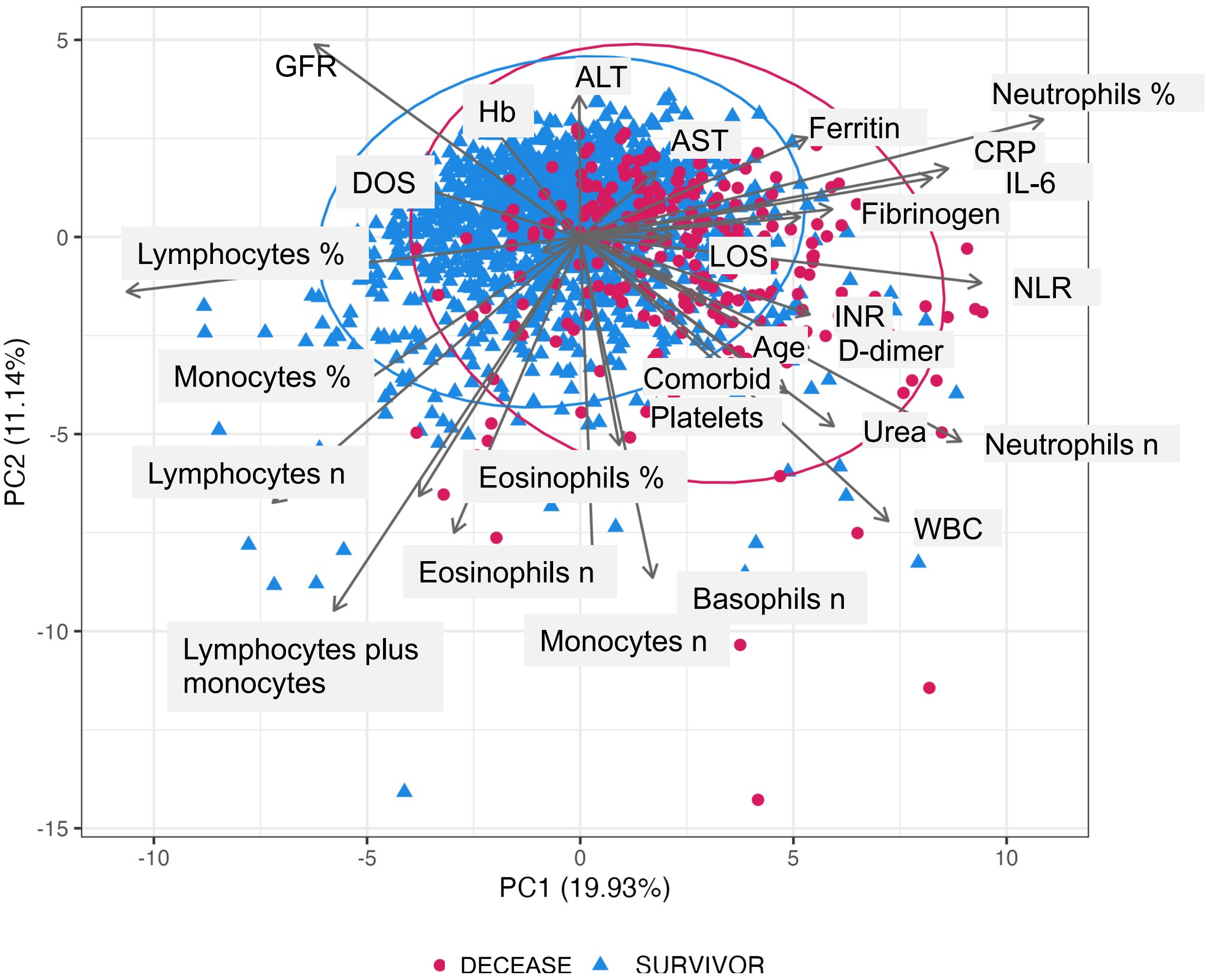

Figure 3S. HUVH cohort. Principal component analysis of the main variables, exitus is equivalent to decease. CRP, C-Reactive Protein; GFR, glomerular filtration rate; Hb, haemoglobin; LOS, length of stay at hospital; NLR, neutrophils to lymphocyte ratio; WBC, Whole Blood Count. The variables to the right of the plot contribute to decease, the variables to the left contribute to survival, the length of the vector and the angle is and indicator of the relative contribution to the outcome. Notice how lymphocyte % and n and neutrophils % and n pull to the opposite outcomes.

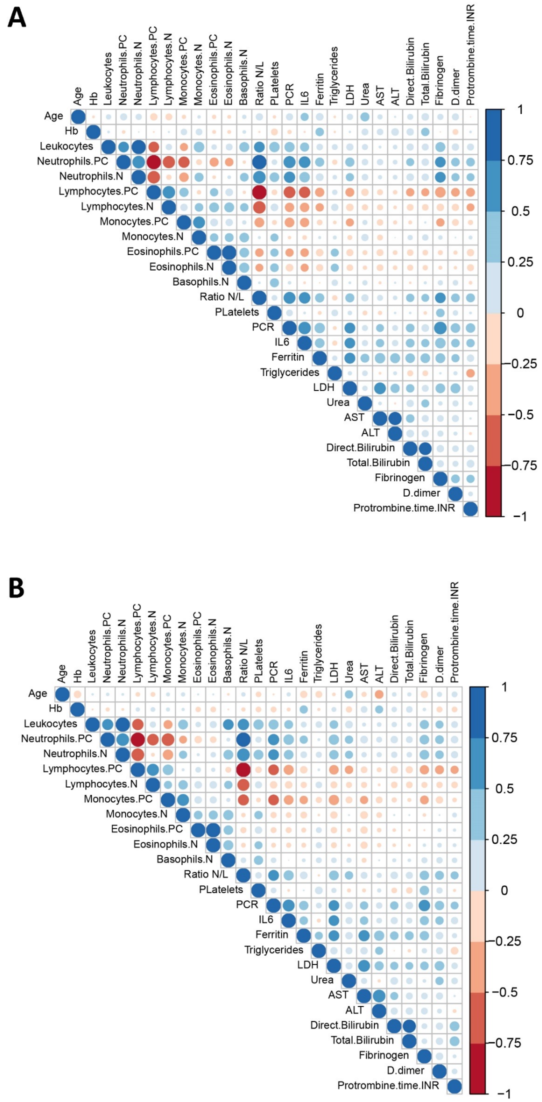

### Figure 4S

Figure 4S. HUVH cohort. Correlograms of the main variables for patients under (A) and over (B) 65 years of age. The main correlations are maintained (see text, "Detailed sequence of statistical analysis…".). The message from this figure is that the network of interactions is not age dependent even if outcome is, this supporting the point that the variables included here miss some crucial pathogenic factors.

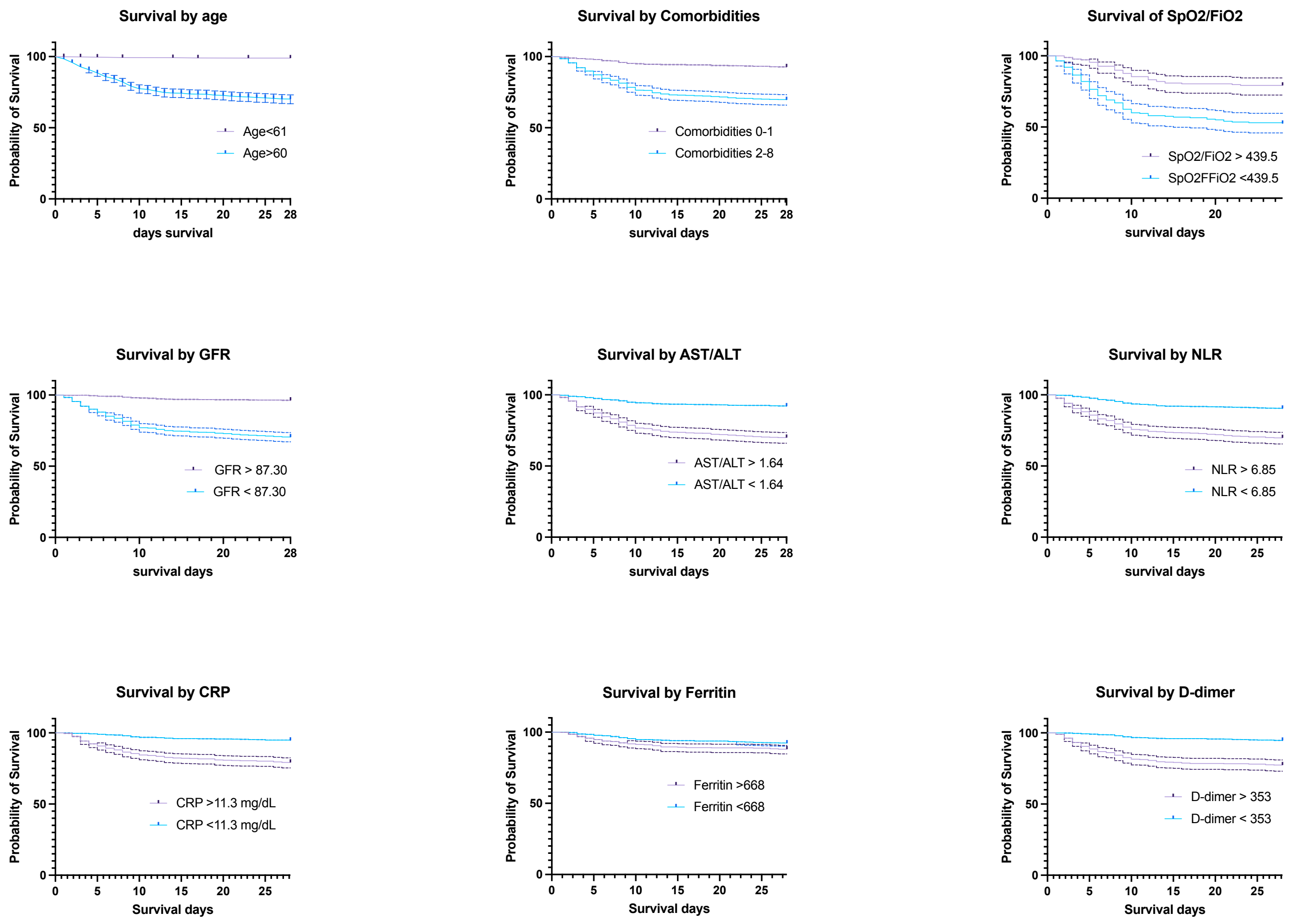

Figure 5S. HUVH cohort. Representative Kaplan-Meyer survival curves clinicodemographic biomarkers (ODRB) and inflammation related biomarkers (IFRB). Cut off points determined by the Youden index in performance clinical laboratory type of ROC curves. The message from this figure is that ODRB are more closely associated to mortality, even if their variations were not wide and they may be easily missed during clinical management.

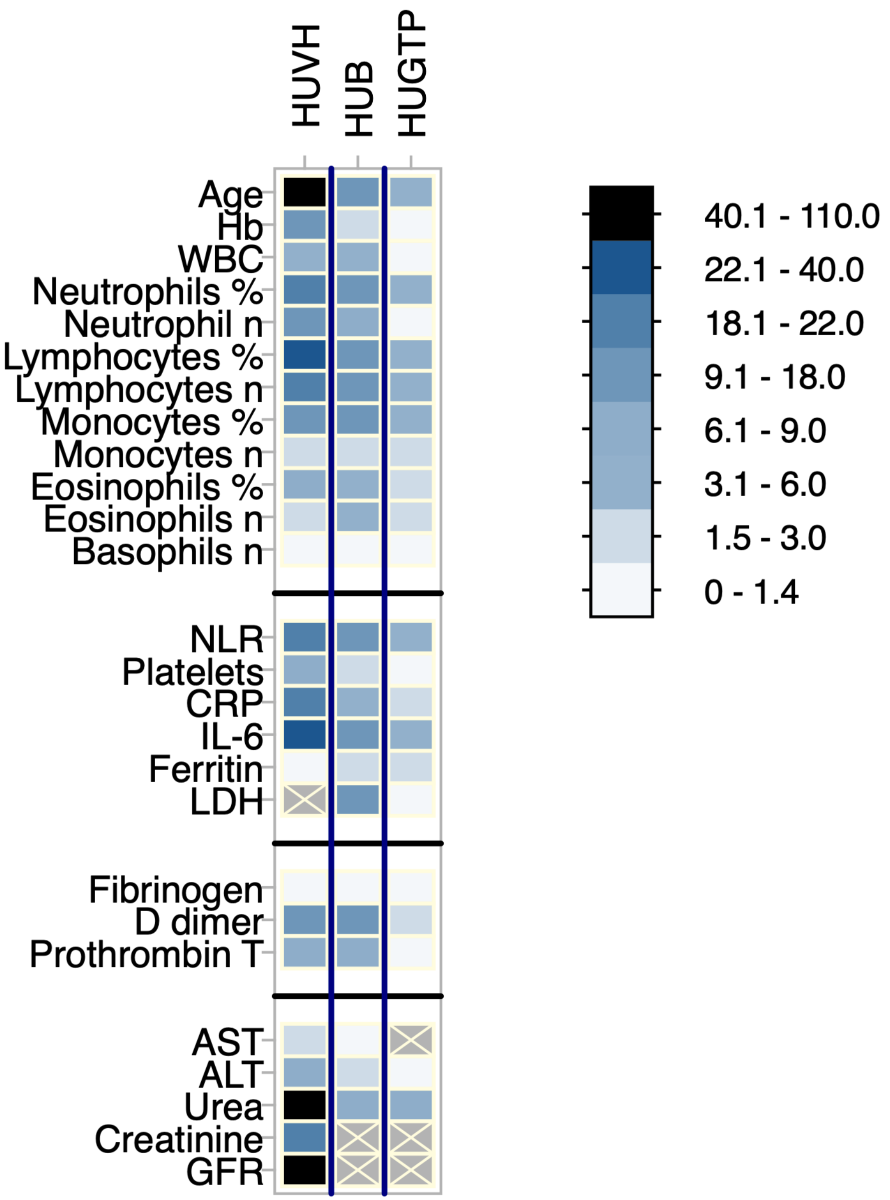

Figure 6S. Three hospital cohorts. Heatmap summarizing pairwise comparison of the survivors vs deceased in the three hospital cohorts; p values scale refer non-parametric comparisons (Mann-Whitney and Kruskal-Wallis). In general p values are not very informative of the weight of each independent variable in the outcome (dependent variable) but when differences are so large, that they are indicative of which variables are more closely associated to outcome, to be corroborated by the subsequent analyses. The heatmap makes visual those differences.

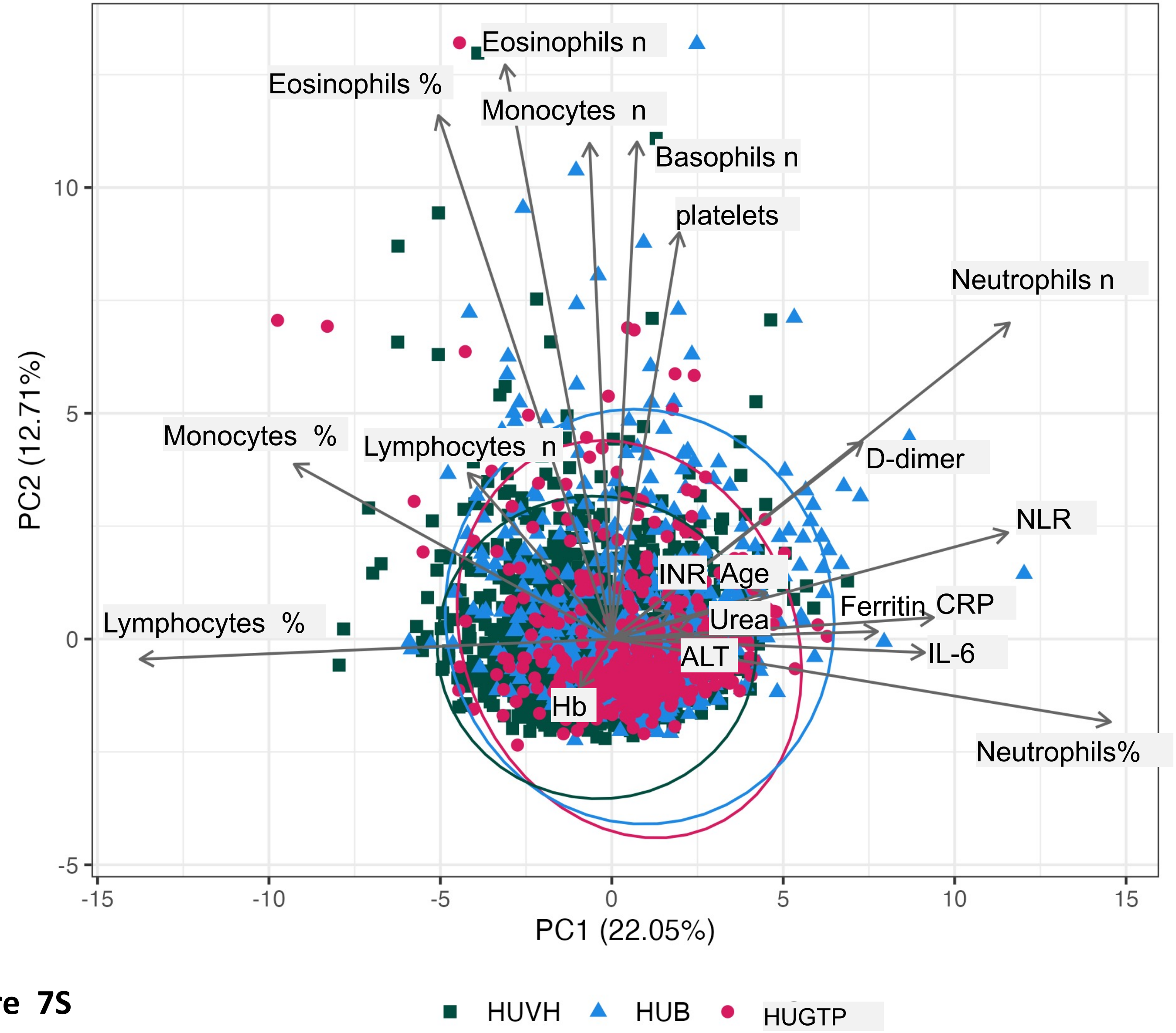

**Figure 7S**

Figure 7S. PCA analysis representation of the three hospitals cohorts, indicating similar main vectors, with local differences. The plot should be compared with plot figure 3S. The variables to the right of the plot contribute to decease, the variables to the left contribute to survival, the length of the vector and the angle is and indicator of the relative contribution to the outcome. Notice how lymphocyte % and n and neutrophils % and n pull to the opposite outcomes

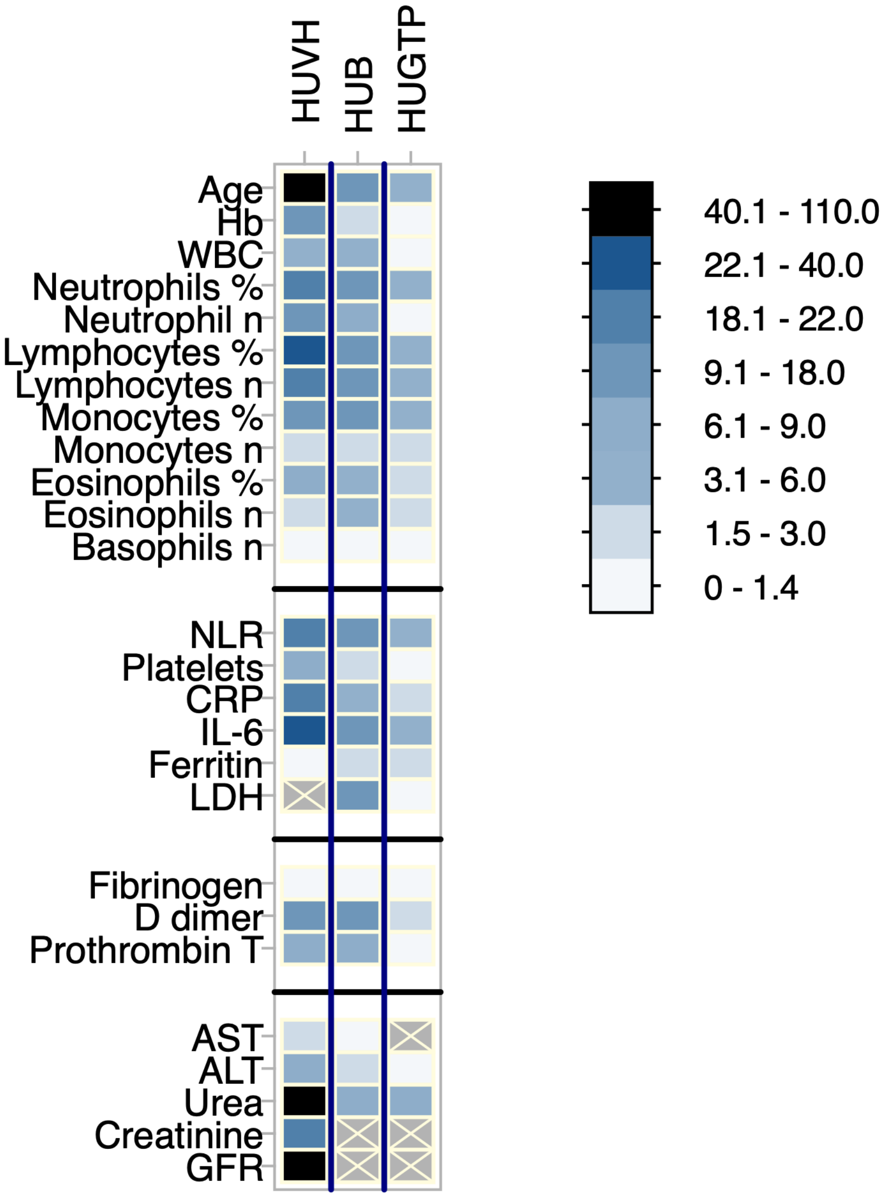

Figure 8S. Heatmap summarizing the reduction of mean Gini index in the Random Forest model that reflects the importance of each variable in the predictive model. The left column represents the values when laboratory and age plus comorbidities models were run separately and the right column, when run combined; notice that in both, age is the dominant variable.

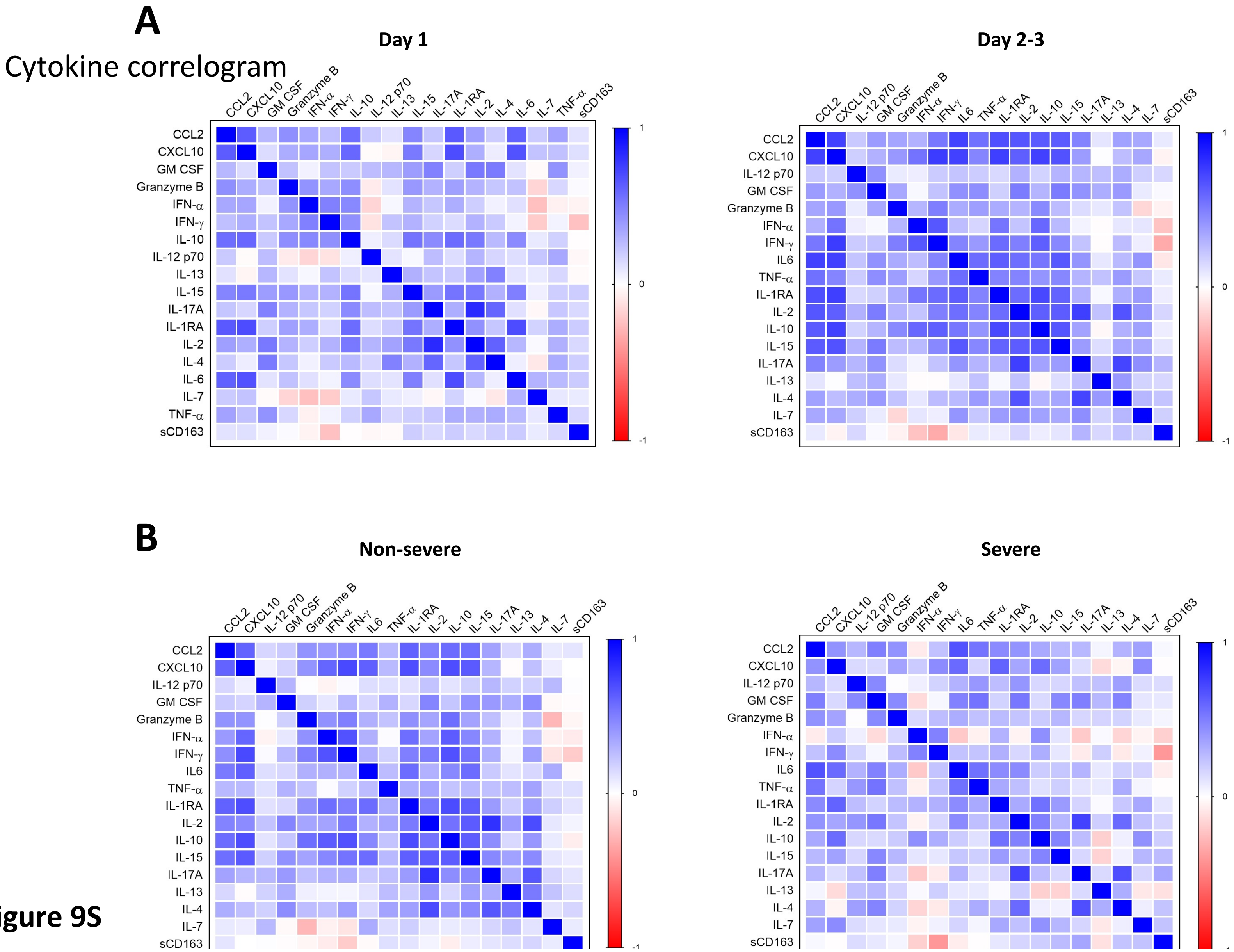

**Figure 9S**

Figure 9S. A, Comparison of correlograms among cytokines initial sample and two/three days late; no major changes are observed. B, Comparison of Non-severe vs severe; some more differences are observed (see supplementary text, subsection cytokines) but not striking, thus indicating that the interval was too short, an indication for future studies.

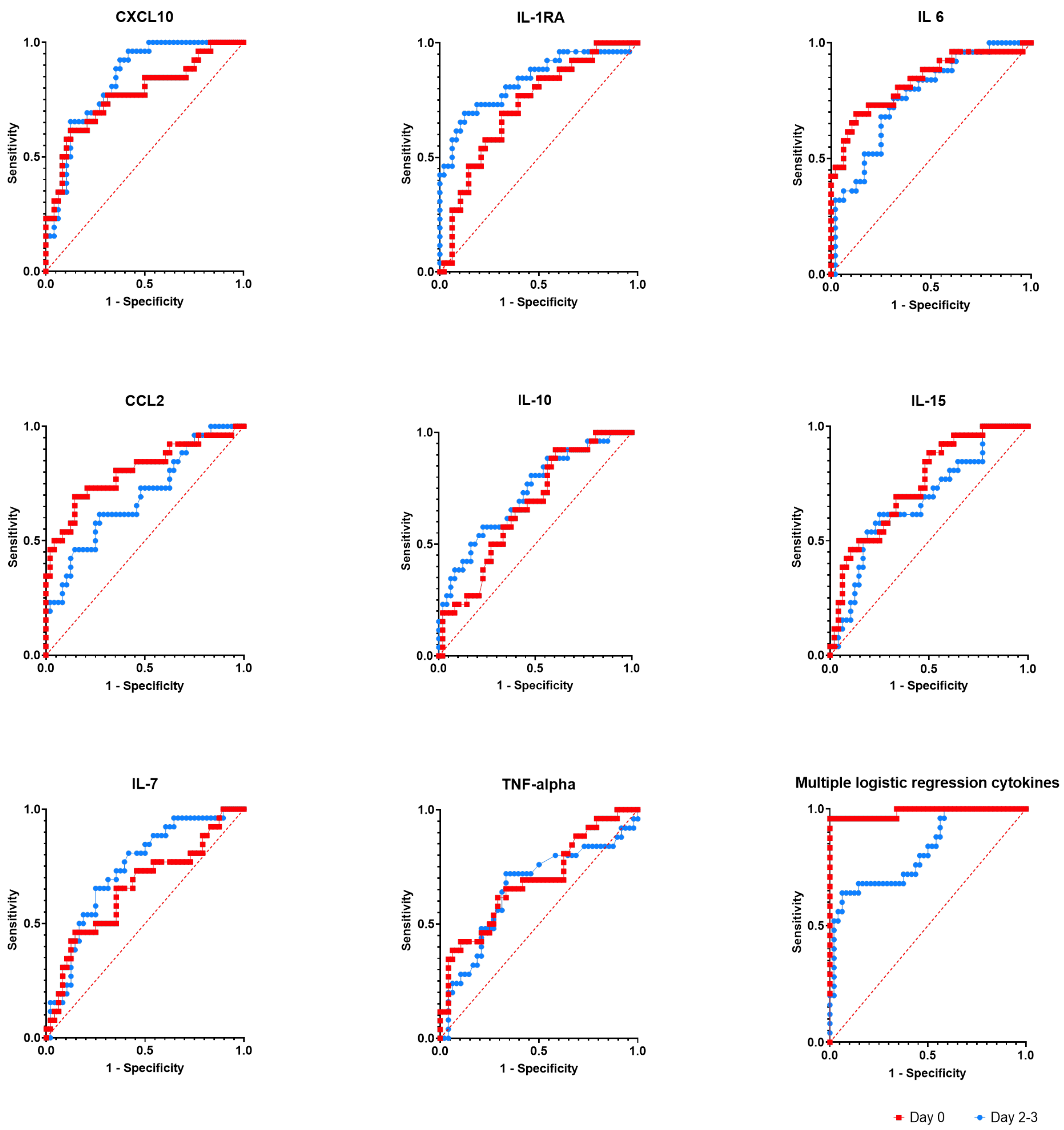

**Figure 10S**

Figure 10S. Performance of cytokines as clinical laboratory test in ROC curve analysis. For values see Table 6, but direct examination of the curves indicates that these variables may be useful for further studies.

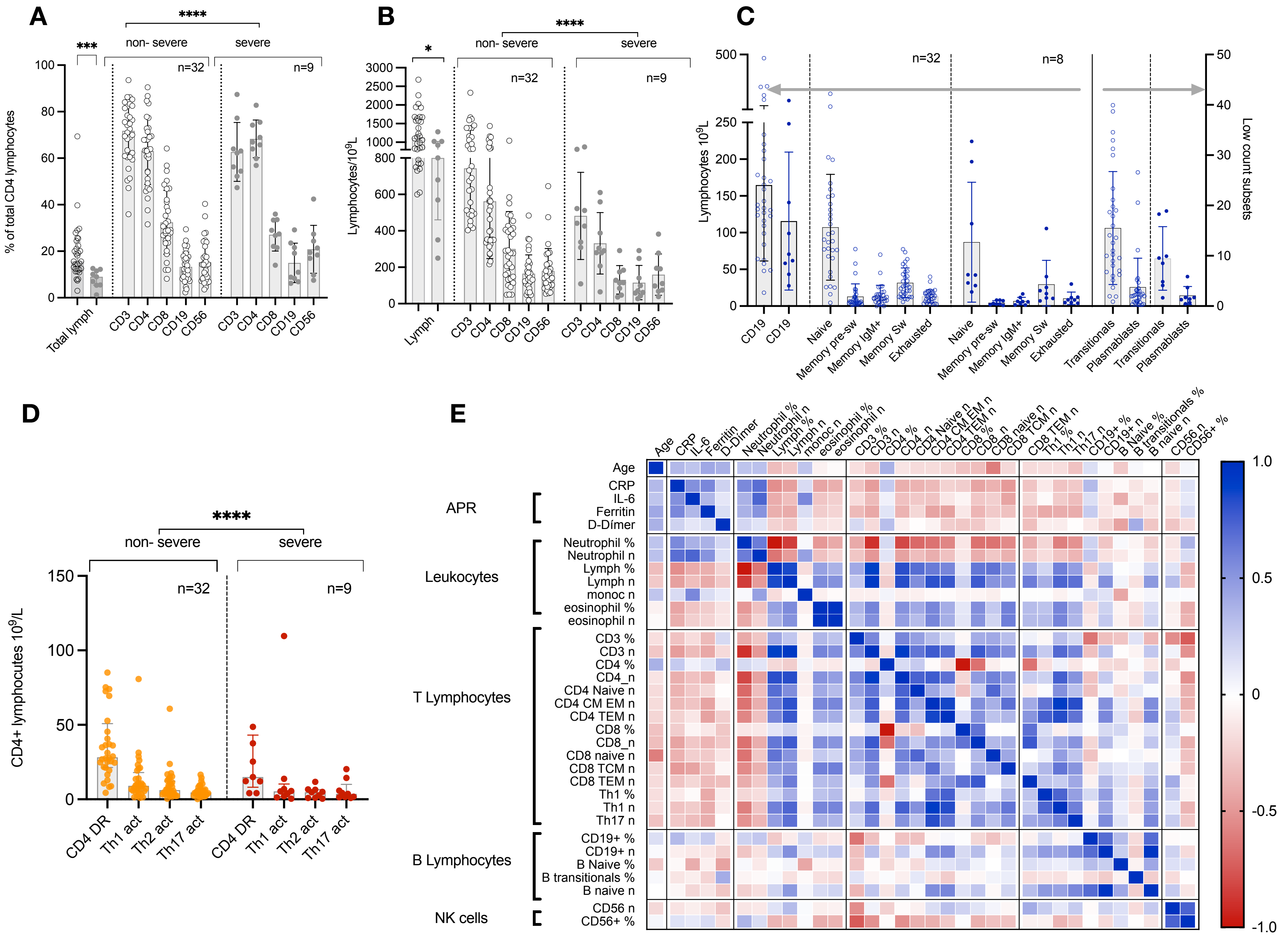
A
B
C
D
E
***
****
*
non- severe
severe
n=32
n=9
n=8
% of total CD4 lymphocytes
Lymphocytes/10$^9$L
Lymphocytes 10$^9$L
Low count subsets
CD4+ lymphocutes 10$^9$/L
Total lymph
Lymph
CD3
CD4
CD8
CD19
CD56
Naive
Memory pre-sw
Memory IgM+
Memory Sw
Exhausted
Transitionals
Plasmablasts
CD4 DR
Th1 act
Th2 act
Th17 act
Age
CRP
IL-6
Ferritin
D-Dímer
Neutrophil %
Neutrophil n
Lymph %
Lymph n
monoc n
eosinophil %
eosinophil n
CD3 %
CD3 n
CD4 %
CD4_n
CD4 Naive n
CD4 CM EM n
CD4 TEM n
CD8 %
CD8_n
CD8 naive n
CD8 TCM n
CD8 TEM n
Th1 %
Th1 n
Th17 n
CD19+ %
CD19+ n
B Naive %
B transitionals %
B naive n
CD56 n
CD56+ %
APR
Leukocytes
T Lymphocytes
B Lymphocytes
NK cells
1.0
0.5
0
-0.5
-1.0

Figure 4

11S. Summary of flowcytometry analyses of peripheral blood mononuclear cells (n=41) following HIPC protocol generated 163 variables corresponding to 42 subsets. A & B, distribution of the main subsets in severe and non-severe patients, by percentual distribution and n; C, distribution of main lymphocyte subsets among severe and non-severe patients; pairwise comparison does not show statistical differences, but the non-parametric multiple comparison by Kruskal-Wallis show p <0.0001. N=32 non sever and 8 severe patients. The right Y axis has been added to better visualize the small populations like transitional B cells and plasmablasts. D. Distribution of subsets of activated T cells among severe and non-severe patients, p<0-0001. E, Correlogram of the different subsets with age, acute phase reactants (APR), and the other leukocytes populations; notice mutual negative correlation between most lymphocyte subsets and both neutrophils and APR. CRP, C-Reactive Protein.
